# Cost-effectiveness of a telemonitoring program in patients with cardiovascular diseases compared to standard of care

**DOI:** 10.1101/2023.05.12.23289904

**Authors:** Andreas Ziegler, Alper Öner, Gisela Quadflieg, Raphael O Betschart, Alexandre Thiéry, Hugo Babel, Henry G Mwambi, Henriette Neumeyer, Steffen Mackschin, Sissy Hintz, Miriam Mann, Hermann Dittrich, Christian Schmidt

**Author notes:** Corresponding author Prof. Dr. Andreas Ziegler, Cardio-CARE, Medizincampus Davos, Herman-Burchard-Str. 1, CH-7265 Davos Wolfgang, Switzerland. **Funding:** The CardioCare MV trial was financed by the Gemeinsamer Bundesausschuss (Federal Joint Committee) within the project HerzEffekt MV (funding code: 01NVF16003). Diagnostic procedures and treatments for NICC and SoC patients were partly covered by the health insurance companies AOK Nordost and TK. The funder/sponsor had no role in the design and conduct of the study; collection, management, analysis, and interpretation of the data; preparation, review, or approval of the manuscript; and decision to submit the manuscript for publication. **Author contributions:** GQ and AZ designed and managed the health economic study with input from all other authors. HD, SH, and MM contributed to the data management and data transfer of the study. HB, HGM, SM, HN, GQ, AT, and AZ developed the statistical analysis plan. HB, ROB, HGM, AT, and AZ performed the statistical analyses. GQ, ROB, and AZ performed the health-economic analyses. HB performed the simulation study. All authors participated in data interpretation. AZ drafted the first and subsequent versions of this paper with input and revisions by all authors, who reviewed and approved the final submitted paper. **Availability of data and materials:** Individual participant data may not be made available to third parties because of the data protection contract for the trial. Study protocol, statistical analysis plan, and the primary analysis have been published in Trials (2018) 19:120, Trials (2020) 21:131, and Eur J Med Res (2023) 28:22, respectively. **Competing interests:** Prof. Ziegler is scientific director of the non-profit research organization Cardio-CARE. He is listed as co-inventor of an international patent on the use of a computing device to estimate the probability of myocardial infarction (International Publication Number WO2022043229A1), and he is shareholder of the ART-EMIS Hamburg GmbH. Dr. Thiéry was and Dr. Babel and Mr. Betschart are bioinformaticians at Cardio-CARE. Prof. Mwambi was visiting professor at Cardio-CARE. Prof. Neumeyer was and Drs. Mackschin and Quadflieg are employees of Philips GmbH Market DACH, Hamburg, Germany. Dr. Quadflieg holds stocks of Royal Philips N.V. All other authors declare no competing interests. **Licence agreement:** The Corresponding Author has the right to grant on behalf of all authors and does grant on behalf of all authors, a worldwide licence to the Publishers and its licensees in perpetuity, in all forms, formats and media (whether known now or created in the future), to i) publish, reproduce, distribute, display and store the Contribution, ii) translate the Contribution into other languages, create adaptations, reprints, include within collections and create summaries, extracts and/or, abstracts of the Contribution, iii) create any other derivative work(s) based on the Contribution, iv) to exploit all subsidiary rights in the Contribution, v) the inclusion of electronic links from the Contribution to third party material where-ever it may be located; and, vi) licence any third party to do any or all of the above.

## Abstract

**Objectives:** The main aim of this work was to analyze the cost-effectiveness of an integrated care concept (NICC) that combines telemonitoring with the support of a care center in addition to guideline therapy for patients. Secondary aims were to compare health utility and health-related quality of life (QoL) between NICC and standard of care (SoC).

**Methods:** The randomized controlled CardioCare MV trial compared NICC and SoC in patients from Mecklenburg-West Pomerania (Germany) with atrial fibrillation, heart failure, or treatment-resistant hypertension. QoL was measured using the EQ-5D-5L at baseline, 6 months, and 1 year follow-up. Quality-adjusted life-years (QALY), EQ5D utility scores, visual analogue scale (VAS) scores, and VAS adjusted life-years (VAS-AL) were calculated. Cost data were obtained from health insurance companies, and the payer perspective was taken in health economic analyses. Quantile regression was used with adjustments for stratification variables.

**Results:** The net benefit of NICC (QALY) was 0.031 (95%CI: 0.012–0.050; p=0.001) in this trial involving 957 patients. EQ5D index values, VAS-ALs and VAS were larger for NICC compared to SoC at 1 year follow-up (all p≤0.004). Direct cost per patient and year were €323 (CI: €157–489) lower in the NICC group. When 2000 patients are served by the care center, NICC is cost-effective if one is willing to pay 10,652€ per QALY per year.

**Conclusion:** NICC was associated with higher QoL and health utility. The program is cost-effective if one is willing to pay approximately 11,000€ per QALY per year.

**WHAT IS ALREADY KNOWN ON THIS TOPIC:**

- Integrated care concepts, i.e., the combination of telemedicine and the reinforcement of patient self-care in a multidisciplinary team together with telephone support, reduce mortality, morbidity, and levels of depression in patients with cardiovascular diseases.

**WHAT THIS STUDY ADDS:**

- The integrated care concept investigated in the CardioCare MV trial was also associated with higher quality of life, higher health utility, and it is cost-effective if one is willing to pay approximately 11,000 € per quality-adjusted life-year (QALY).

**HOW THIS STUDY MIGHT AFFECT RESEARCH, PRACTICE OR POLICY:**

- With the demonstrated cost effectiveness of the integrated care concept, policy makers and health insurance companies are enabled to decide on the introduction of these concepts as standard of care.

## Introduction

Cardiovascular diseases (CVDs) are a major economic burden on health care systems. According to the German Federal Statistical Office, the cost caused by CVD amounted to 44.4 billion Euro in 2015 for Germany. Early detection and management using counselling and medicines may reduce CVD morbidity and mortality (1). However, patient management may be challenging in rural areas, such as Mecklenburg-West Pomerania, the least densely populated and least industrialized state in Germany. In such areas, the combination of telemedicine and reinforcement of patient self-care in a multidisciplinary team together with telephone support is expected to be beneficial in CVDs. We thus implemented an integrated care concept (NICC), which combines telemonitoring, an integrated care network, and guideline therapy. We applied NICC in three CVDs, atrial fibrillation (AF), heart failure (HF) and treatment-resistant hypertension (TRH).

The main purpose of this work was to analyze the cost-effectiveness of NICC from the payer perspective using the EQ-5D-5L quality of life (QoL) questionnaire (2) in combination with individual costs data provided by health insurance companies. Secondary aims were to compare the health-related QoL and visual analogue scale (VAS) adjusted life-years (VAS-AL) between NICC and standard of care (SoC).

## Methods

### CardioCare MV trial

Data for this study are primarily based on the CardioCare MV trial, which was an open-label, two parallel groups trial conducted in Mecklenburg-Western Pomerania (Germany) in patients with AF, HF and/or TRH. Details of the trial design have been published elsewhere (3, 4). Inclusion and exclusion criteria of the trial are listed in Table 1, trial details and the CHEERS checklist are provided in Supplementary Material 1.

**Table 1.** Inclusion and exclusion criteria for the CardioCare MV trial.

|  |
| --- |
| **Inclusion criteria**<br>1. Heart failure (I50, NYHA II-IV) or atrial fibrillation (I48, European Heart Rhythm Association (EHRA) II-IV) or resistant hypertension (I10-15, $\geq 3$ antihypertensive medicines from different drug classes with systolic blood pressure $> 140/90$ mmHg or $\geq 4$ antihypertensive medicines from different drug classes).<br>2. Member of health insurance company AOK Nordost or Techniker Krankenkasse.<br>3. Inscription to integrated care contract with the health insurance company.<br>4. Residence in Mecklenburg-Vorpommern.<br>5. Age $\geq 18$ years.<br>6. Written informed consent. |
| **Exclusion criteria**<br>1. Pregnancy, suspected pregnancy or breast-feeding period.<br>2. Participation in other clinical trials up to 30 days before inclusion in this trial.<br>3. Cognitive deficits.<br>4. Chronic kidney disease. |

The NICC has been described in detail in (3). In brief, the care center was available 24/7 and utilized an electronic patient management platform. Patients provided information from home about their health status using a tablet. They received feedback about their therapy, their measurements, education, reminders, and motivation to follow care plans. The patient situation was evaluated at least on a daily basis, and necessary adjustments were coordinated with care providers.

Patients in the SoC group were treated according to guidelines of the European Society of Cardiology (ESC).

### Health-related quality of life

QoL was measured using the EuroQoL EQ-5D-5L (2); for details, see Supplementary Material 1. EQ-5D-5L health states were converted into a single index value, ranging from 1 (best health) to 0 (death), as was the single negative index value. Scores of the best and worst health states on the visual analog scale (VAS) were 100 and 0, respectively. Quality-adjusted life years (QALYs) and VAS adjusted life years (VAS-AL) after transformation to the 0 to 1 scale were estimated as the area under the curve (AUC).

### Cost data

The participating health insurance companies provided individual costs data for the 1-year follow-up period and for the 1-year period prior to randomization. Direct cost was the sum of hospitalization costs, ambulant costs, and prescription costs. Program costs were considered separately (see below). The cost difference between treatment groups was investigated with adjustment for the 1-year period prior to randomization. Details on cost data and the original analysis plan of the health insurance data are provided in Supplementary Material 1.

### Analysis of health utility and quality of life data

Details of the statistical analysis including the imputation model is provided in Supplementary Material 1. Mean QALY differences and EQ5D at 1-year follow-up between treatment groups were estimated using linear regression after multiple imputation using MICE (5). Sensitivity analyses included the complete case analysis as well as two reduced multiple imputation models.

### Analysis of costs data from health insurance companies

Details of the statistical analysis is provided in Supplementary Material 1. Costs were log-transformed after a +1 shift to achieve symmetric distributions in the comparison of treatment groups. Transformed costs were analyzed by median regression and by linear regression with adjustments for stratification variables and the costs in the year prior to randomization.

### Cost-effectiveness analysis

Details on the cost-effectiveness analysis is provided in Supplementary Material 1. In brief, life expectancy was obtained from data provided by the German Statistical Office for the new federal states (6). The discount rate was assumed to be 3%; sensitivity analyses were performed with 0% and 5% discount rates. Incremental cost per QALY were calculated. Costs for medical devices were taken from actual costs. Costs for nurses and trained cardiologists were based on actual costs (€50,000, and €160,000 annually). We assumed annual infrastructure costs of €100,000 in case of 1000 patients and of €150,000 in case of 2000 patients served by the care center. Software costs are based on actual costs and were €600 per patient.

We employed two different models for estimating cost-effectiveness from the payer perspective. Both models are described in detail in Supplementary Material 1. In brief, model 1 was a cohort-based model, where both survival and costs for the population were simulated. Model 2 was a projection model with fixed survival probabilities and QALYs. Two costs were of primary interest, cost in the first year and cost until median survival time (MST).

Calculations were done using R version 4 and R Markdown. The fixed cost-effectiveness model was estimated using MS Excel (program code: Supplementary Material 2).

## Results

### Patients

Between December 2017 and August 2019, 957 patients were randomly assigned either to NICC or SoC, stratified by primary diagnosis and inpatient/outpatient treatment. Detailed assessments were at 6 months and 1 year after randomization. The CONSORT flowchart has been provided in (7). The primary diagnosis was AF, HF, and TRH in 265 (27.7%), 406 (42.4%), and 286 (29.9%) patients, respectively. Mean age plus/minus standard deviation of the population was 71.3±10.5 years, and the proportion of males was 61.1%. Detailed descriptive statistics are provided in Table 2. Data transfer from health insurance companies was permitted for 923 patients (463 NICC, 460 SoC).

**Table 2.**
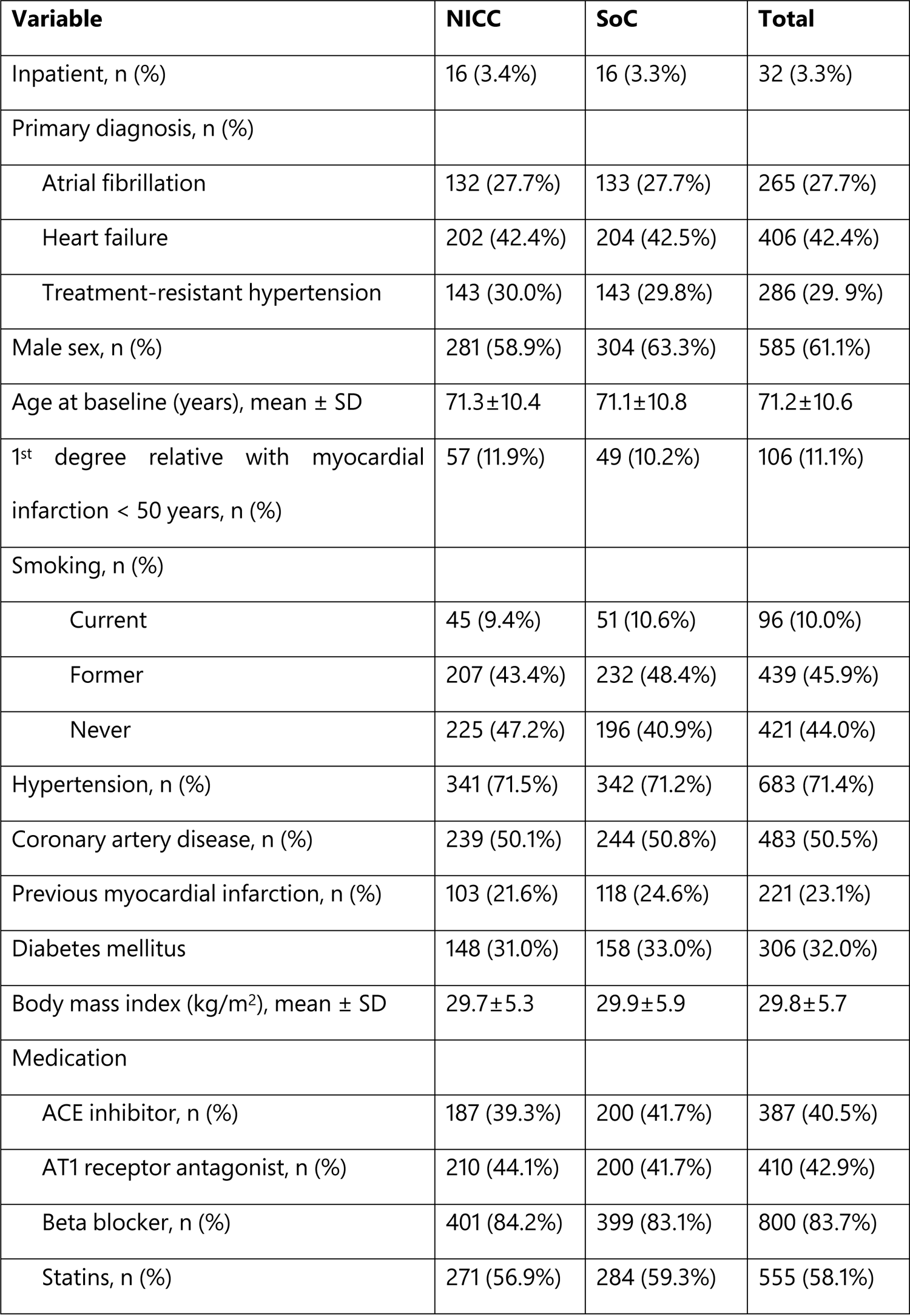
Baseline characteristics of patients in the CardioCare MV trial.

| Variable | NICC | SoC | Total |
| --- | --- | --- | --- |
| Inpatient, n (%) | 16 (3.4%) | 16 (3.3%) | 32 (3.3%) |
| Primary diagnosis, n (%) |  |  |  |
| Atrial fibrillation | 132 (27.7%) | 133 (27.7%) | 265 (27.7%) |
| Heart failure | 202 (42.4%) | 204 (42.5%) | 406 (42.4%) |
| Treatment-resistant hypertension | 143 (30.0%) | 143 (29.8%) | 286 (29.9%) |
| Male sex, n (%) | 281 (58.9%) | 304 (63.3%) | 585 (61.1%) |
| Age at baseline (years), mean ± SD | 71.3±10.4 | 71.1±10.8 | 71.2±10.6 |
| 1<sup>st</sup> degree relative with myocardial infarction < 50 years, n (%) | 57 (11.9%) | 49 (10.2%) | 106 (11.1%) |
| Smoking, n (%) |  |  |  |
| Current | 45 (9.4%) | 51 (10.6%) | 96 (10.0%) |
| Former | 207 (43.4%) | 232 (48.4%) | 439 (45.9%) |
| Never | 225 (47.2%) | 196 (40.9%) | 421 (44.0%) |
| Hypertension, n (%) | 341 (71.5%) | 342 (71.2%) | 683 (71.4%) |
| Coronary artery disease, n (%) | 239 (50.1%) | 244 (50.8%) | 483 (50.5%) |
| Previous myocardial infarction, n (%) | 103 (21.6%) | 118 (24.6%) | 221 (23.1%) |
| Diabetes mellitus | 148 (31.0%) | 158 (33.0%) | 306 (32.0%) |
| Body mass index (kg/m<sup>2</sup>), mean ± SD | 29.7±5.3 | 29.9±5.9 | 29.8±5.7 |
| Medication |  |  |  |
| ACE inhibitor, n (%) | 187 (39.3%) | 200 (41.7%) | 387 (40.5%) |
| AT1 receptor antagonist, n (%) | 210 (44.1%) | 200 (41.7%) | 410 (42.9%) |
| Beta blocker, n (%) | 401 (84.2%) | 399 (83.1%) | 800 (83.7%) |
| Statins, n (%) | 271 (56.9%) | 284 (59.3%) | 555 (58.1%) |

### Quality of life and quality-adjusted life-years

Figure 2 shows the relative change in problems from baseline to 1-year follow-up for NICC and SoC separately for all items of the EQ5D. For all items, fewer patients reported problems in the NICC compared to the SoC group, and all dimensions of the EQ5D questionnaire at 1-year follow-up but pain (p=0.057) showed differences between NICC and SoC at p≤0.002 (Supplementary Figure 1 for further details).

**Figure 1.**
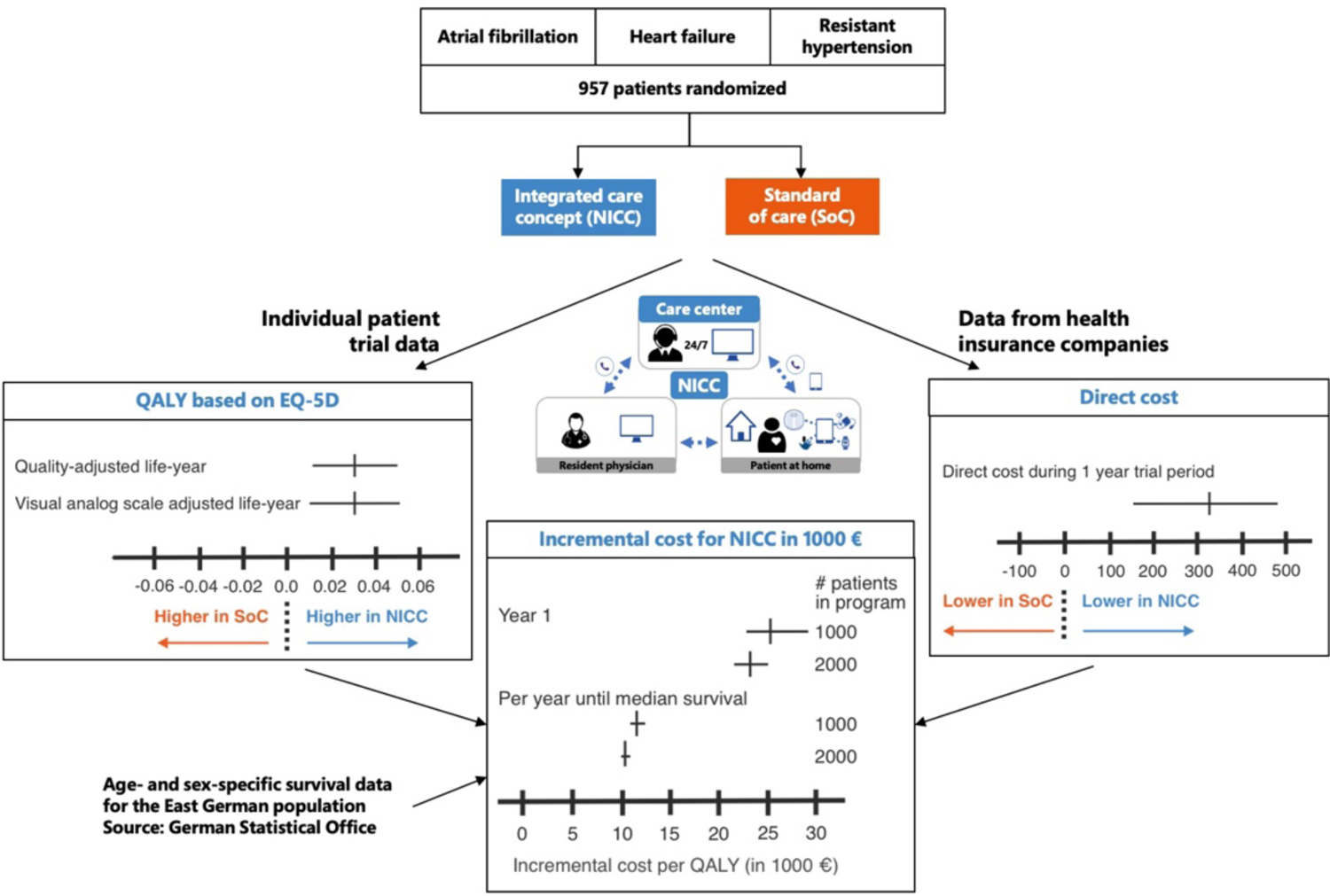
Summary of the main findings of the study. Vertical and horizontal bars correspond to mean and 95% confidence interval estimates, respectively.

**Figure 2.**
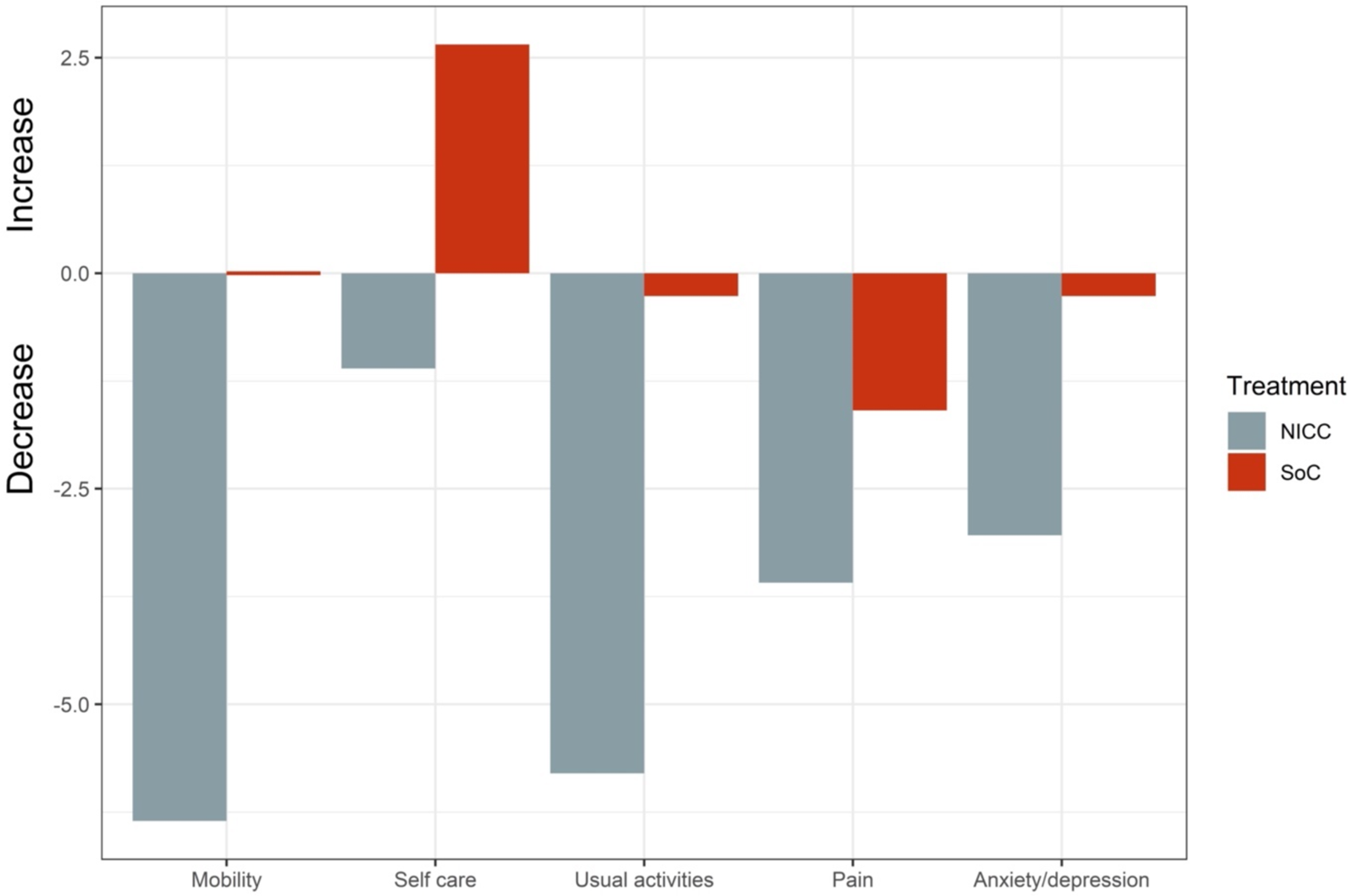
Difference (percentage) in any problems between 1-year follow-up for the EQ5D questionnaire. Displayed are all items separately for the integrated care concept (NICC (grey)) and standard of care (SoC (red)) using complete cases. More patients in the SoC group had problems with self care compared to baseline. The percentage of patients with problems decreased in the NICC group for all items and for all items more than in the SoC group.

Supplementary Figures 2 and 3 display the EQ5D index and the EQ5D VAS distribution for both treatment groups at baseline, 6 months, and 1 year follow-up, respectively. The EQ5D index distributions were similar for both groups at baseline, and differences between treatment groups were present at both follow-ups (Supplementary Figure 2). Results were similar for the EQ5D VAS (Supplementary Figure 3).

**Figure 3.**
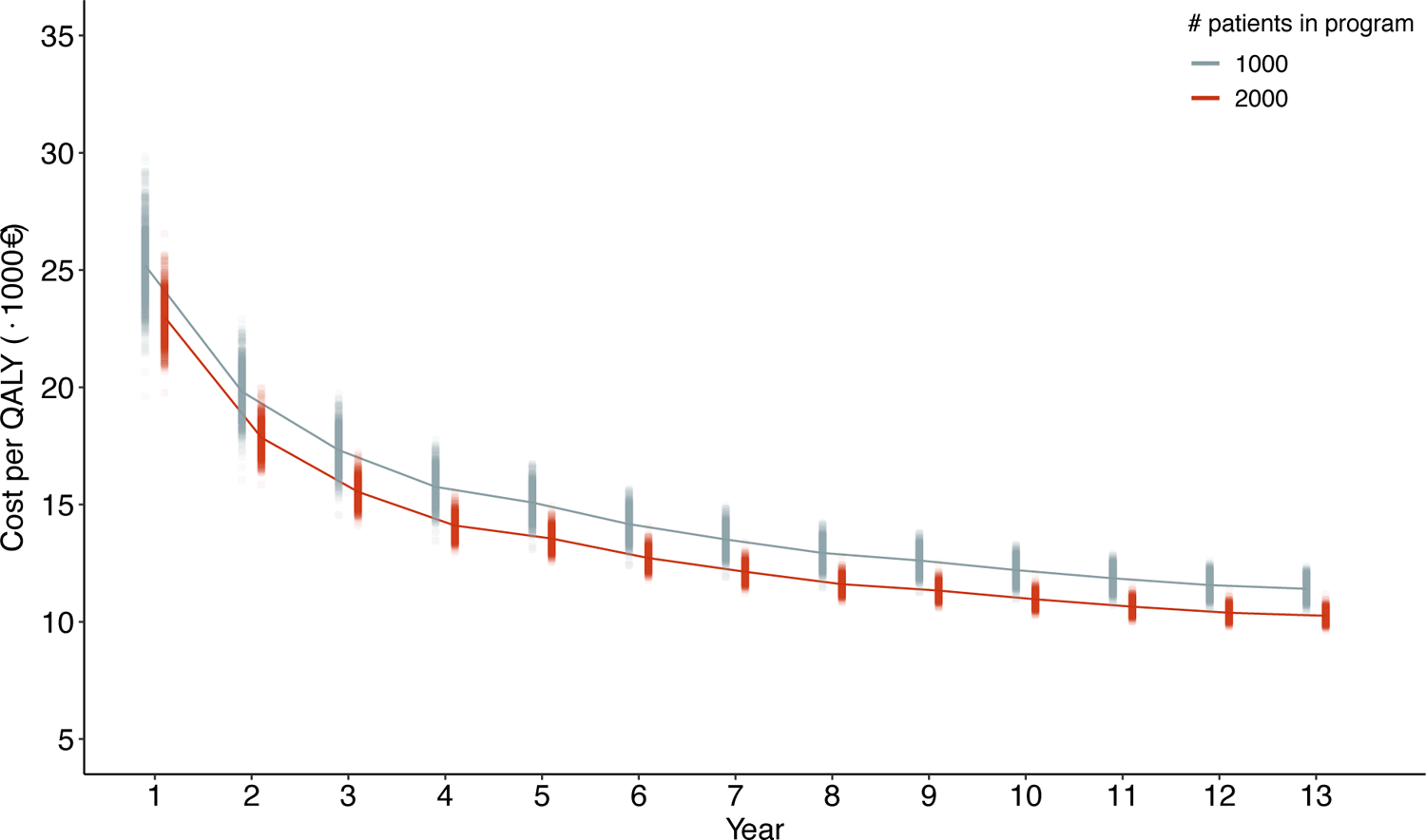
Cost-effectiveness analysis from the simulated cohort data. Incremental cost per QALY are displayed per year for direct cost and the case of 1000 patients and 2000 patients, respectively, served by the care center. Incremental cost was averaged up to the year displayed. The observed cost difference between NICC and SoC for the direct cost were considered in the simulation model. The discount rate was 3%. Details on the simulation model are provided in Supplementary Material 1.

The minimally important difference (MID) of the EQ5D index was assumed to be 0.03 for chronic HF (8, 9). Disease-specific MIDs have neither been reported for AF nor for TRH. The EQ5D index at follow-up was 0.034 higher for NICC compared to SoC (95% confidence interval (CI) 0.011–0.056; p=0.003; Supplementary Table 1). NICC thus exceeded the MID.

Health utility outcomes for the difference between NICC and SoC are displayed in Figure 1. Patients in the NICC group had higher QALY than patients in the SoC group (0.031; CI 0.012–0.050; p=0.001; for details, see Supplementary Table 2). The effect of NICC compared to SoC was similar for the VAS-AL (0.031; CI 0.010–0.053; p<0.001). The net benefit of NICC over SoC can therefore be assumed to be 0.031 QALY per patient and year. The sensitivity analysis using complete cases showed an even higher net benefit for NICC over SoC (Supplementary Table 1). Simple group comparisons are displayed in Supplementary Table 2 for health utility and QoL outcomes by primary diagnosis. When many baseline variables, including sex and age, and other patient-reported outcomes were added to the regression model, there was a positive treatment effect for NICC compared to SoC (Supplementary Material 3).

### Cost-effectiveness analysis: direct cost

Table 3 provides descriptive statistics and results from regression analysis for direct cost and components of direct cost. Median direct cost was €3045 (quartiles €1108 and €8200) in the NICC group and €3298 (€1185–€9474) in the SoC group. Hospitalization costs were asymmetric: median and lower quartile were €0 in both treatment groups (Table 3), and several patients had hospitalization costs exceeding €100,000 in the 1-year observation period. Because medians of hospitalization costs were €0, median regression could not converge for hospitalization costs. However, hospitalization costs were the main driver of total direct costs. Specifically, the costs per hospitalization day were approximately €707, obtained as ratio of average costs per case (€5088 per stay) (10) and average stay (7.2 days per stay) (11). The number of inpatient days in hospital were approximately 1.5 lower in the NICC group (CI 0.96–1.81) (7). Therefore, direct cost was expected to be lower in the NICC group compared to SoC. Indeed, direct cost was approximately €322.55 lower in the NICC group compared to the SoC group during the 1-year treatment period (CI €156.56–€488.53; Table 3).

**Table 3.**
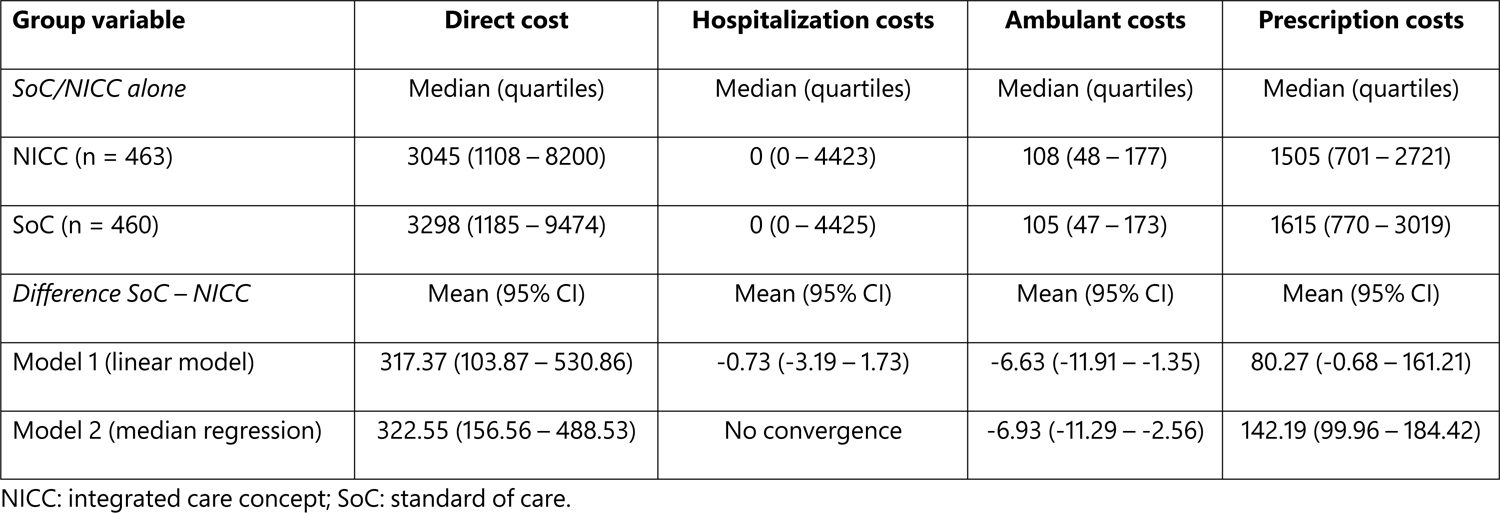
Cost analysis in the CardioCare MV trial using data obtained from health insurance companies. Data transfer from health insurance companies was permitted for 923 of 957 randomized patients.

### Cost-effectiveness analysis: Incremental cost per quality-adjusted life-years

Figure 3 displays the incremental cost per QALY averaged over the duration of the program for 1000 patients served by the care center, when the cohort was simulated (dynamic model) and all cost, i.e., direct cost and cost for operating the care center of the NICC were considered at a discount rate of 3%. Detailed results are shown in Table 4; Supplementary Figures 4 and 5 display the incremental cost at 0% and 5% discount rates, respectively. The incremental cost per QALY in the first year were on average €25,217 and €22,987 for 1000 patients and 2000 patients served by the care center, respectively (Figure 3). Incremental cost per QALY were thus €2230 lower per QALY in the care center serving 2000 patients compared to the care center serving 1000 patients (Table 4). Figure 4 displays the cost-effectiveness acceptability curve for varying scenarios for direct costs using the dynamic cost-effectiveness model at a discount rate of 3% with 1000 simulated cohorts per scenario until the MST of approximately 11 years. If one considers a care center serving respectively 1000 and 2000 patients at a threshold of 97.5% (low in Table 4), the willingness to pay (WTP) is €12,571 and €11,597 per QALY. If one conservatively assumes no difference in direct cost between NICC and SoC, the WTP per QALY was €14,513 and €13,289 in case of 1000 and 2000 patients served by the care center, respectively.

**Figure 4.**
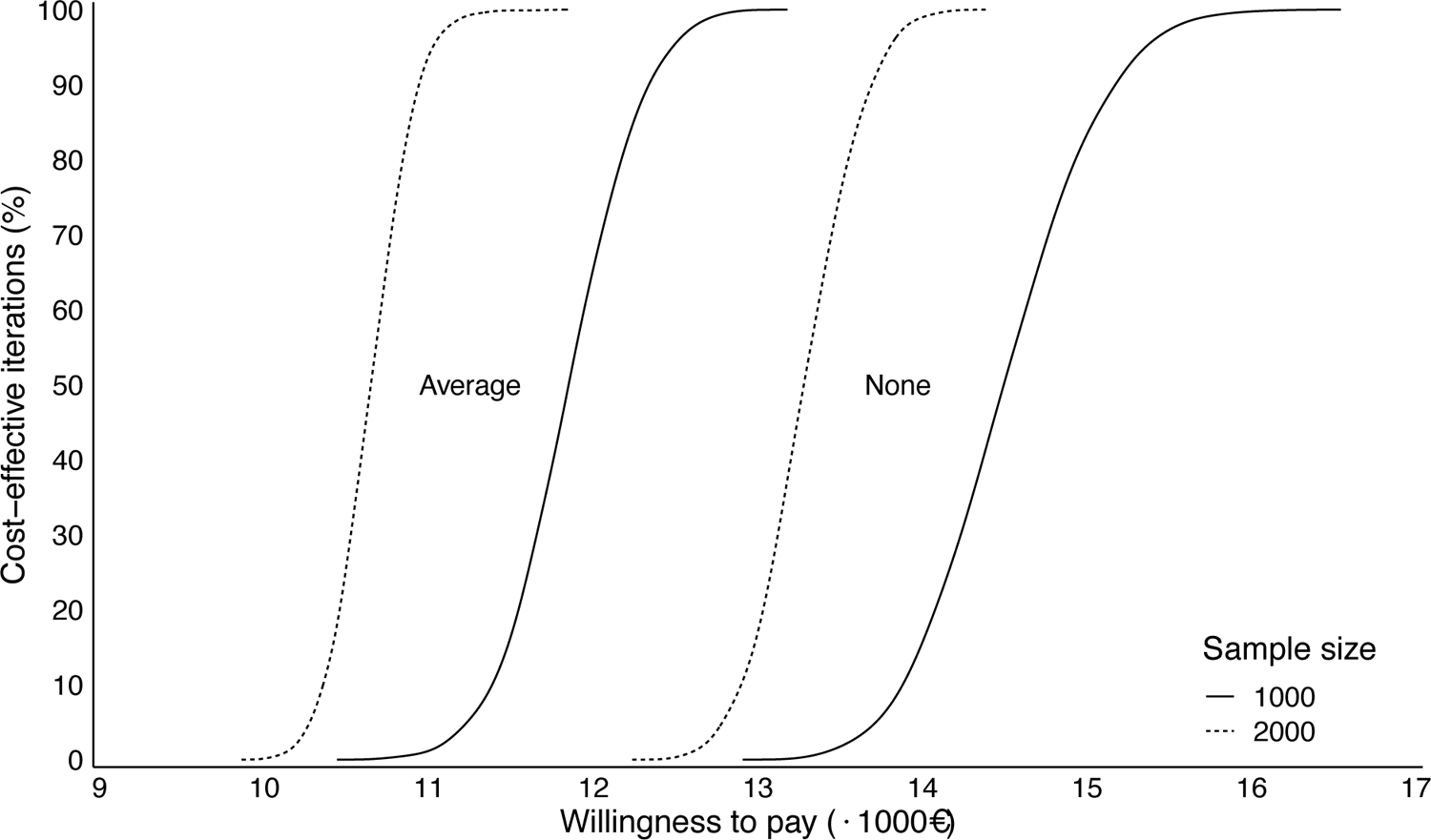
Cost-effectiveness acceptability curve for the integrated care concept (NICC) from the dynamic cost-effectiveness model using a discount rate of 3%. Cost per year are averaged until median survival time of the cohort. The two left curves display the cost-effective iterations (in %) by willingness to pay (in €1000) from 1000 simulations, when the average direct cost difference between NICC and SoC was considered. The two right curves display the cost-effective iterations (in %) by willingness to pay (in €1000), when the no difference in direct cost between NICC and SoC was assumed. Solid and dashed lines correspond to a care center, which serves 1000 and 2000 patients, respectively.

**Table 4.** Cost-effectiveness analysis in the CardioCare MV trial. Displayed are the incremental cost per QALY for year 1 and the average incremental cost until median survival time (MST) in the NICC group. MST was 11 years for the dynamic cohort-based simulation model, and it was 14 years and 10 years for the static life table and Weibull models, respectively. Models are described in detail in Supplementary material 1, and Supplementary Material 3 provides the calculations for the static models. Average saving: average of direct cost saving for NICC compared to SoC during the 1-year observation period. Low and high correspond to the 95% CI, and none corresponds to no difference in direct cost between NICC and SoC.

| Survival model | # patients in care center | Savings in direct cost (€) | Cost per QALY year 1 (€) | Cost per QALY year averaged until MST (€) |  |  |
| --- | --- | --- | --- | --- | --- | --- |
| Dynamic model |  |  |  | Discount 0% | Discount 3% | Discount 5% |
| Cohort-based | 1000 | Average | 25,216 | 13,433 | 11,856 | 10,986 |
| simulation model |  | None | 29,146 | 16,440 | 14,513 | 13,449 |
|  |  | Low | 28,157 | 14,235 | 12,571 | 11,669 |
|  |  | High | 22,604 | 12,644 | 11,135 | 10,296 |
|  | 2000 | Average | 22,987 | 12,070 | 10,652 | 9,870 |
|  |  | None | 26,908 | 15,055 | 13,289 | 12,314 |
|  |  | Low | 24,836 | 12,560 | 11,089 | 10,283 |
|  |  | High | 21,180 | 11,597 | 10,220 | 9,463 |
| Static model |  |  |  |  |  |  |
| Life table | 1000 | Average (322.55) | 27,300 | 15,237 | 14,590 | 14,226 |
|  |  | None (0) | 32,379 | 18,149 | 17,502 | 17,137 |
|  |  | Low (156.56) | 29,914 | 16,735 | 16,088 | 15,724 |
|  |  | High (488.53) | 24,684 | 13,738 | 13,091 | 12,727 |
|  | 2000 | Average (322.55) | 24,890 | 13,856 | 13,229 | 12,875 |
|  |  | None (0) | 29,970 | 16,768 | 16,140 | 15,787 |
|  |  | Low (156.56) | 27,504 | 15,355 | 14,727 | 14,374 |
|  |  | High (488.53) | 22,275 | 12,357 | 11,730 | 11,377 |
| Weibull model | 1000 | Average (322.55) | 27,290 | 12,866 | 11,419 | 12,286 |
|  |  | None (0) | 32,368 | 15,169 | 14,804 | 14,589 |
|  |  | Low (156.56) | 29,903 | 14,051 | 13,686 | 13,471 |
|  |  | High (488.53) | 24,675 | 11,680 | 11,315 | 11,101 |
|  | 2000 | Average (322.55) | 24,881 | 11,774 | 13,229 | 11,211 |
|  |  | None (0) | 29,959 | 14,077 | 13,722 | 13,514 |
|  |  | Low (156.56) | 27,494 | 12,959 | 12,604 | 12,396 |
|  |  | High (488.53) | 22,267 | 10,588 | 10,234 | 10,026 |

Table 4 provides the incremental cost per QALY using the static projection model for two different survival models, a life table model and a Weibull survival model, for patients served by the care center. In analogy to the dynamic model, two scenarios were considered for the care center, with either 1000 or 2000 patients being served by the care center. Incremental cost per QALY for the first year of the program varied from €22,267 to €32,379. Incremental cost per QALY were approximately €2400 lower in the care center serving 2000 patients compared to the care center serving 1000 patients. Incremental cost per QALY varied from €10,234 to €14,804, when the average of the cost over the period until MST was considered. MST was approximately 14 years for the life table model and approximately 10 years for the Weibull model (Table 4).

## Discussion

In this study, we observed different QALY, VAS-ALs, VAS values and QoL values overall and for all dimensions but pain between the NICC and the SoC treatment groups. The mean EQ5D index value at 1 year follow-up was 0.797 in the NICC group for patients with HF and 0.034 lower in the same patients in the SoC group (p=0.003). The QALY was 0.031 (CI: 0.012–0.050) points lower in the SoC group compared to the NICC group (p<0.001).

While EQ5D index values did not show a relevant change in the NICC group, they decreased in the SoC group. In contrast, the difference on the VAS scale between the two groups was due to an increase of the VAS from baseline to 1 year follow-up in the NICC group. Here, VAS values were similar in the SoC group at all time points. The results of this study are in line with the literature. For example, Mizukawa and colleagues (12) showed that collaborative self-management through interactive communication via telemonitoring led to improved QoL in patients with HF. Other trials in patients with HF also demonstrated the advantage of telemonitoring (13), including QoL. A review of systematic reviews (14) showed that telemonitoring and home telehealth were effective in reducing HF rehospitalization and mortality. Other interventions, such as the use of mobile phone–based monitoring require further investigation.

### How the work fits with the literature

Most patients with HF in this trial had New York Heart Association (NYHA) class II (n=282, 66.7% of all patients with HF), and 29.1% (n=123) of the HF patients had NYHA class III. Similarly, 56.5% (n=126) of the patients with AF had American Heart Association (AHA) class B, and 35.5% (n=150) AHA class C. Our findings are in line with the literature. Specifically, the EQ5D index was between 0.7 and 0.8 on average if between 0% and 33% of the patients with HF had NYHA classes III or IV (15), and it was approximately 0.8 in patients with NYHA II stadium, while it was approximately 0.65 in patients with NYHA III – IV failure (16). In patients with AF, a systematic review reported mean EQ5D index values between 0.65 and 0.95, with the majority being around 0.8 (17). The same work provided an overview on VAS estimates from the EQ5D, and VAS means varied from 65 to just below 90 points; most studies had a mean VAS between 70 and 75 (17). In contrast, comparable work on the EQ5D in TRH is scarce. Glybochko and colleagues (18) investigated the EQ5D in 14 patients with AF who underwent renal denervation. Carris and Smith (19) reviewed QoL in patients with TRH, but this study did not include the EQ5D. Overall, EQ5D index values and VAS values in this trial were similar to those reported in the literature for patients with similar disease severity.

### Strengths

Findings were consistent irrespective of the way data were analyzed for all EQ5D endpoints considered in this work. For example, adjustments for baseline variables did not alter the presence of significant differences between the treatment groups for the EQ5D variables.

In the cost-effectiveness analysis, we showed that the average cost per QALY was between approximately €10,000 and €20,000 for the time period until MST (Figure 4), and cost per QALY was approximately €2000 lower when the care center served 2000 patients rather than 1000. The establishment of larger care centers therefore seems to be reasonable from a cost perspective.

### Cost-effectiveness analysis

Figure 4 and Table 4 show that NICC is cost-effective with 97.5% probability at a WTP per QALY of respectively €11,089 and €12,571 when 2000 and 1000 patients are served by the care center and when the observed difference in direct cost between NICC and SoC is considered. With average savings in direct cost between NICC and SoC, cost-effectiveness is obtained at WTP of €11,856 and €10,652 for a care center serving 1000 and 2000 patients, respectively. In case of no difference in direct cost between NICC and SoC, NICC with 1000 and 2000 patients is cost-effective at a WTP of €14,513 and €13,289 per QALY per year, respectively. The National Institute for Health and Care Excellence in the United Kingdom accepts cost of £20,000 to £30,000 per QALY (20), which corresponds to approximately €22,700 and €34,000, respectively. NICC can therefore be considered to be cost-effective. This holds true even for its first year of operation (cost per QALY in year 1: €22,987), when 2000 patients are served by the care center.

### Limitations

One limitation of this work is that the analysis of the EQ5D data from this trial is a secondary analysis. The primary analyses investigated duration and frequency of hospitalizations over the 1-year follow-up period and two composite endpoints, where the first composite endpoint consisted in mortality, stroke, and/or myocardial infarction within the first year after treatment assignment. The second composite endpoint consisted in the components of the first composite endpoint plus cardiovascular decompensation. The two composite endpoints as well as many secondary endpoints, such a mortality showed an advantage for NICC when compared with SoC (7). However, the trial did neither show a significant difference in the number of hospitalization days nor a significant difference in the number of hospitalizations between the two treatment groups. For patients in the NICC group, the treating physicians recommended, however, a visit to an established doctor or even a hospital in case patients had unusual telemonitoring findings or called the care center. This might explain differences between the NICC and the SoC group in mortality rates and the two composite endpoints but the lack of superiority of NICC concerning hospitalization endpoints.

Another limitation of this study is the short observation period for the randomized controlled trial (7). The treatment and follow-up period in the trial was limited to 1 year per patient because the trial had to be completed within three years after its initiation. However, a long-term follow-up of all study patients is intended at 5 years after randomization (3).

### Future research

Future research should investigate which component of NICC have the largest positive effect on health status, QoL and survival, and whether it is possible to further increase the positive treatment effect.

## Conclusion

QoL and health utility were higher in the NICC group compared to patients in the SoC group. The difference between treatment groups was clinically relevant for both the EQ5D index and the VAS. Direct cost per patient and year was €323 (CI: €157–489) lower in the NICC group. Cost was lower when more patients were served by a care center due to synergy effects within the care center. When respectively 1000 or 2000 patients are served by the care center, NICC is cost-effective if one is willing to pay 11,856€ and 10,652€ per QALY per year, when NICC is continued until median survival time.

## Data Availability

Individual participant data may not be made available to third parties because of the data protection contract for the trial. Study protocol, statistical analysis plan, and the primary analysis have been published in Trials (2018) 19:120, Trials (2020) 21:131, and Eur J Med Res (2023) 28:22, respectively.

## Acknowledgements

Members of the CardioCare MV Study Group: Dr. H. Bleschke, Med. T. Buchner, Dr. C. Buckow, Dr. K. Bunge, Dr. S. Duda, Dipl. Med. H. El-Sourani, Dr. K. Frey, Dipl. Med. H. Greiner-Leben, Dr. F. Henschel, Dr. R. Hering, Dr. O. Knispel, Dr. J. Kram, Dr. A. Martschewski, PD Dr. R. Mitusch, Dr. S. Plietzsch, Dr. S. Rausch, Dr. A. Rink, Dr. M. Wejda, Dr. R. Wißmann, Dr. B. Wolff.

