## Supplementary Material 1 for "Cost-effectiveness of a telemonitoring program in patients with cardiovascular diseases compared to standard of care"

Running title: Cost-effectiveness in cardiovascular telemonitoring

Ziegler et al.

#### Supplementary Material 1

#### Table of Content

*Table of Content* ... *1*

1. *Supplementary Methods* ... *2*
   1.1 The CardioCare MV trial ... 2
      1.1.1 CardioCare MV trial population ... 2
      1.1.2 Intervention: the integrated care concept (NICC) ... 2
      1.1.3 Control ... 3
      1.1.4 Health-related quality of life (QoL) ... 3
   1.2 Cost data ... 3
      1.2.1 Permission for use ... 3
      1.2.2 Definition of costs ... 4
      1.2.2 Statistical analysis of the EQ-5D data ... 4
      1.2.3 Cost effectiveness analysis ... 5
   1.3 Projection model for estimating incremental cost per QALY ... 6
      1.3.1 Cost for medical devices ... 6
      1.3.2 Costs for physicians and nurses ... 6
      1.3.3 Costs for infrastructure and software ... 6
      1.3.4 Proportion of patients with specific primary heart disease ... 6
      1.3.5 Sex proportions ... 7
      1.3.6 Survival probabilities for the new states from the German population ... 7
      1.3.7 Life table approach ... 7
      1.3.8 Weibull distribution approach ... 7
      1.3.9 Log-logistic distribution approach ... 8
   1.4 Simulation model for estimating cost per QALY ... 9

2. *Health Economic Analysis Concept: Important Research Questions and Hypotheses* ... *11*
   2.1 Overview of the research questions and hypotheses ... 11
      2.1.1 Inpatient treatment and inpatient treatment costs ... 11
      2.1.2 Outpatient treatment and outpatient treatment costs ... 11
      2.1.3 Prescription costs ... 12
      2.1.4 Total costs ... 12
   2.2 Methodological aspects ... 12
   2.3 Specific hypotheses ... 13
      2.3.1 Inpatient area ... 13
      2.3.2 Outpatient area ... 14
      2.3.3 Prescription costs ... 15
      2.3.4 Total cost ... 15

### 1. Supplementary Methods

#### *1.1 The CardioCare MV trial*

The CardioCare MV trial was a randomized controlled trial, which was designed as open-label, bi-center, parallel-group design with two groups and blinded observer in patients with atrial fibrillation, heart failure and/or treatment-resistant hypertension. Details of the study design, population and clinical outcomes have been published elsewhere [1 2]. The study protocol was approved by the Ethics Committee of Rostock University Medical Faculty on 18.07.2017, file number: A 2017-0117. The trial was registered with drks.de, registration number: DRKS00013124, registration date: 05.10.2017, and secondarily with ClinicalTrials.gov, registration number: NCT03317951, registration date: 17.10.2017, both prior to recruitment start.

##### 1.1.1 CardioCare MV trial population

A total of 957 patients were enrolled from a single center in the trial between December 2017 and August 2019. Patients were randomly assigned to treatment with either NICC or SoC. Randomization was done with stratification by primary diagnosis and inpatient/outpatient treatment. Nine hundred twenty-five (96.66%) patients were recruited as outpatients. The primary diagnosis was atrial fibrillation, heart failure and treatment-resistant hypertension in 265 (27.69%), 406 (42.42%) and 286 (29.89%) patients, respectively. Descriptive statistics at baseline are provided in Table 1 for the trial population. Patients were followed up for 1-year, and detailed assessments were at 6 months and 1 year after randomization. The CONSORT flowchart of patients has been provided elsewhere [3].

##### 1.1.2 Intervention: the integrated care concept (NICC)

The NICC has been described in detail elsewhere [1]. In brief, the heart of the NICC was its care center, which was available 24/7. The care center utilized the NICC platform for patient management from the distance. Using the NICC tablet, patients could provide information from home about their health status via vital-sign measurements and by answering questionnaires using a secure communication channel. Patients received feedback about their therapy and their measurements, education as well as reminders and motivation to follow care plans. With this, the patient situation was evaluated on at least a daily basis. Therapy and necessary adjustments were coordinated with care providers of the patient.

###### 1.1.3 Control

Patients not randomized to NICC were treated according to current practice as described in the guidelines of the European Society of Cardiology (ESC) [4-6].

###### 1.1.4 Health-related quality of life (QoL)

QoL was measured using the EuroQoL EQ-5D-5L in its German paper-based version [7]. The EQ-5D-5L is a validated, generic patient-reported outcome measure covering 5 health domains (mobility, self-care, usual activities, pain/discomfort and anxiety/depression) and a visual analogue self-rating scale (VAS). Patients were asked to rate severity of their current problems (level 1 = no problems, level 2 = slight problems, level 3 = moderate problems, level 4 = severe problems, 5 = extreme problems). Patients can therefore be classified into $5^5$ = 3125 health states plus two further states, i.e., unconscious, and dead. EQ-5D-5L health states were converted into a single index value using the Crosswalk Index Value Calculator based on data from the German population [8], ranging from 1 (best health) to -0.205. Patients who died during the observation period were set to 0 as was the single negative index value; the value at 6 months follow-up of that patient was -0.074. As sensitivity analysis, only surviving patients were considered [9]. For the VAS, patients were asked to rate their own health relative to full health (score=100) and worst imaginable health state (score=0). The QALY was estimated for each individual as the area under the curve (AUC) through linear interpolation from EQ5D index values for the periods between measurements [10]. VAS-AL were calculated analogously after transformation to the 0 to 1 scale to ensure comparability with utility-based QALY [11].
EQ-5D-5L analyses were done separately for each level and each category. In addition, "problems" (levels 2-5) vs. "no problems" (level 1) were considered for each category and the index score.

##### *1.2 Cost data*

###### 1.2.1 Permission for use

The participating health insurance companies provided individual costs data in € for the 1-year follow-up period as well as for the 1-year period prior to randomization. Direct cost was calculated as the sum of hospitalization costs, ambulant costs, and prescription costs. The definition of these costs is provided in the next section.
Permission for data transfer was granted on March 19, 2020, for the AOK Nordost and on February 12, 2020, for the Techniker Krankenkasse. Expansion of run time and expansion of the data set were approved on January 6, 2021, and December 30, 2020, for the AOK Nordost and the Techniker Krankenkasse, respectively. Permission to data was granted only for variables specified in a health economic analysis plan, which is available on request from the

corresponding author. Data were transferred through a data custodian, and the specific health insurance company may not be identified in the analyses according to the permission granted by the supervisory authorities. The data transfer from the health insurance companies included data for 923 of the total 957 patients (463 NICC, 460 SoC). The data from 34 (3.55%) patients fell under data protection regulations and were therefore not transferred to the data custodian.

##### 1.2.2 Definition of costs

We defined direct cost as the sum of hospitalization costs, ambulant costs, and prescription costs. The corresponding terms were defined as follows:

*Hospitalization:* "Datei enthält für die Versicherten der Studiengruppe alle geschlossenen stationären Fälle aus DTA §301, die im Beobachtungszeitraum begonnen haben. Krankenhausfälle mit Beginn vor oder Ende nach Beobachtungszeitraum werden nicht berücksichtigt. Datei enthält keine Fälle aus ambulanter Behandlung oder ambulante OP im Krankenhaus. Es werden die bundeseinheitlichen Schlüssel benutzt."

*Translation:* "For the insured persons of the trial, the file contains all closed inpatient cases from DTA (Data Exchange Procedure) §301 that started during the observation period. Hospitalization cases beginning before or ending after the observation period are not considered. File does not contain cases from outpatient treatment or outpatient surgery in hospital. National keys are used."

*Ambulant costs:* "Kostenwerte der ambulanten, budgetär und extrabudgetär vergüteten Leistungen in Euro, inklusive Dialyse und Sachkosten."

Translation: "Costs of outpatient, budgetary and extrabudgetary services in Euros, including dialysis and material costs."

*Prescription:* "Datei enthält für die Versicherten der Studiengruppe alle Arzneimittel und Hilfsmittel aus Apotheken, die im Beobachtungszeitraum verordnet wurden. Es werden die bundeseinheitlichen Schlüssel aus DTA §300 benutzt."

*Translation:* "For the insured persons in the study group, the file contains all drugs and aids from pharmacies that were prescribed during the observation period. The nationwide keys from DTA (Data Exchange Procedure) §300 are used".

##### 1.2.2 Statistical analysis of the EQ-5D data

Descriptive statistics were calculated as absolute and relative frequencies for categorical variables and as means and standard deviations/medians and quartiles for continuous variables.

Mean QALY differences between treatment groups were estimated using linear regression models controlling for stratification variables of the trial, i.e., primary cardiovascular diagnosis and patient status. The same approach was used to determine the effect on the EQ5D index at 1-year follow-up. Missing values for scores were imputed using MICE [12] with 100 imputations and 10 iterations and predictive mean matching for all variables. The code used for the large imputation model is available in Supplementary Material 1. Sensitivity analyses included the complete case analysis as well as two reduced multiple imputation models. Transformations of the original variables were calculated after imputation.

In one sensitivity analysis, adjustments were made for patient-reported outcomes (PROs), including the EQ5D VAS at baseline, the four domains of the beliefs about medicine questionnaire (BMQ) [13], the generalized anxiety disorder scale (GAD-7) [14], the global score of the health-related quality of life assessment (HeartQoL) [15], the two domains of the illness-specific social support scale short version-8 (ISSS-8) [16], the medication adherence report scale (MARS) [17], the patient activation measure (PAM-13D), measured on the quantitative 0 to 100 scale [18], the patient health questionnaire (PHQ-9) [19] and the world health organization well-being index (WHO-5) [20]. All PROs were used in their validated German version in a paper-based mode. Item non-response from patient-reported outcomes were handled according to the respective manual.

Costs were log-transformed after a shift by +1 to achieve symmetric distributions in the comparison of treatment groups. Transformed costs were analyzed by quantile regression for the median and linear regression with adjustments for stratification variables and the costs in the year prior to randomization. Because costs data were complete for all patients with data transfer permission, we performed a complete case analysis on the costs data.

##### 1.2.3 Cost effectiveness analysis

Life expectancy was obtained from data provided by the German Statistical Office for the new federal states [21]. Mean age of the population was 71 years, and the proportion of males was 61% (Table 1). The discount rate was assumed to be 3%; sensitivity analyses were performed with 0% and 5% discount rates.

For the cost effectiveness analysis incremental cost per QALY were calculated. Details on costs is provided in Supplementary Material 1. In brief, costs for medical devices were taken from the actual costs. Costs for trained cardiologists and for nurses were based on actual costs (€160,000 and €50,000 annually, including employer contribution). We assumed annual infrastructure costs to cost € 100,000 in case of 1000 patients served by the care center and the infrastructure cost to be at € 150,000 in case of 2000 patients served by the care center. These costs were calculated from required office space. Software costs were based on required costs and were € 600 per patient.

We employed two different models for estimating cost effectiveness from the payer's perspective. Both models are described in detail in Supplementary Material 1. In brief, the first model was a projection model with fixed survival probabilities and QALYs, while the second cohort-based model simulated survival and costs for the population. Two costs were of primary interest, cost in the first year and cost until median survival time.

Calculations were done by using R version 4 in conjunction with R Markdown. The fixed cost effectiveness model was estimated using MS Excel and is available as Supplementary Material 3.

#### *1.3 Projection model for estimating incremental cost per QALY*

Supplementary Material 3 provides the MS Excel implementation of all model calculations for the projection model. In this section, we justify the cost used in the projection model and in the cohort simulations.

##### 1.3.1 Cost for medical devices

Cost for medical devices were taken from actual cost, and these were

- EUR 85 for a blood pressure meter, replacement every 5 years, available to all patients,
- EUR 125 for a pulse oximeter, replacement every 3 years, available to patients with atrial fibrillation and patients with heart failure,
- EUR 60 for a scale, replacement every 10 years, available to patients with heart failure, and
- EUR 290 for a tablet, replacement every 3 years, available to all patients.

##### 1.3.2 Costs for physicians and nurses

The common number of working days per year is 220. If 20 days for educational purposes and inability to work are subtracted, 200 days can be assumed as days available in the care center. With this assumption, 1.8 trained cardiologists are required for serving 365 days. For the night service, we assumed a collaboration with a hospital and added 0.5 trained cardiologists for this service. This totals to 2.3 trained cardiologists à EUR 160,000 in case of 1000 patients to be served by the care center. Furthermore, we experienced that 10 nurses à EUR 50,000 can serve 1000 patients. In case of 2000 patients served by the care center we assumed three trained cardiologists and 20 nurses to be working in the care center.

##### 1.3.3 Costs for infrastructure and software

We assumed annual infrastructure costs (office rent, computer system, workplace, telephone system) to cost

- EUR 100,000 in case of 1000 patients served by the care center and the infrastructure cost to be at
- EUR 150,000 in case of 2000 patients served by the care center.
- Software cost were EUR 600 per patient.

##### 1.3.4 Proportion of patients with specific primary heart disease

- Atrial fibrillation: 27.7%,
- Heart failure: 42.4%,
- Treatment-resistant hypertension: 29.9%.

1.3.5 Sex proportions

Male patients: 61.1%

1.3.6 Survival probabilities for the new states from the German population

Age- and sex-specific survival probabilities were obtained for the East German population. Specifically, mortality tables 2018/2020 were downloaded from https://www.destatis.de/DE/Themen/Gesellschaft-Umwelt/Bevoelkerung/Sterbefaelle-Lebenserwartung/Publikationen/Downloads-Sterbefaelle/periodensterbetafeln-bundeslaender-5126204207005.xlsx?__blob=publicationFile on Nov 8, 2021.

1.3.7 Life table approach

In one approach, it was assumed that survival probabilities in the NICC group followed those in the general population. Supplementary Table 2 provides median survival times and hazard ratios for several studies investigating telemonitoring in patients with heart failure. Estimated hazard ratios (HR) were:

- Hindricks et al. [22]: 0.36 (0.17 – 0.74)
- Koehler et al. [23]: 0.70 (0.50 – 0.96)
- Öner et al. [3]: 0.19 (0.07 – 0.56)

For the NICC group, we used mortality rates from the general East German population to avoid any overoptimism. Indeed, 1-year mortality rates in the NICC group were lower (1%) than in the general East German population (2.2%). For the SoC group, we calculated 1-year survival probabilities by using the HRs from the studies which reported HRs.

1.3.8 Weibull distribution approach

In a sensitivity analysis, we estimated the survival probabilities using the Weibull distribution and using the log-logistic distribution.

Four grid searches were performed for the Weibull survival distribution:

1. NICC (intervention group): survival probability after 1 year < 0.99 and after 30 years < 0.02
2. NICC (intervention group): survival probability after 1 year < 0.995 and after 30 years < 0.01
3. Control (standard of care): survival probability after 1 year between 0.93 and 0.947 and after 30 years < 0.01
4. Control (standard of care): survival probability after 1 year between 0.94 and 0.96 and after 30 years < 0.01

In all four cases, the hazard rate parameter λ was searched up to a precision of $10^{-4}$ and k with a precision of $10^{-3}$. The parameter constellation with the largest median survival time (MST) was selected. The R code is provided in Section 5 of this document.

Optimal parameters were:

1. NICC: $\lambda = 0.0726$, $k = 1.753$ with values
   - Survival probability after 1 year: 0.9900

- Survival probability after 20 years: 0.1462
- Survival probability after 30 years: 0.0200
- Median survival time: 11.18

2. NICC: λ = 0.0714, k = 2.006 with values
   - Survival probability after 1 year: 0.9950
   - Survival probability after 20 years: 0.1300
   - Survival probability after 30 years: 0.0100
   - Median survival time: 11.67
3. Control: λ = 0.1076, k = 1.305 with values
   - Survival probability after 1 year: 0.947
   - Survival probability after 20 years: 0.660
   - Survival probability after 30 years: 0.0100
   - Median survival time: 7.02
4. Control: λ = 0.1001, k = 1.389 with values
   - Survival probability after 1 year: 0.9600
   - Survival probability after 20 years: 0.0726
   - Survival probability after 30 years: 0.0100
   - Median survival time: 7.67

The formulae for the Weibull distribution are:

- Survival probability for time past t: $P(T > t) = exp\big(-((\lambda \cdot t)^k)\big)$
- Median survival time: $\mathrm{MST} = \big(\ln(2)\big)^{(1/k)}/\lambda$

##### 1.3.9 Log-logistic distribution approach

The log-logistic distribution is a two-parameter distribution, where event rates increase initially and decrease later. This is, however, not in accordance with the model expected in this application, where especially in the later years of age, i.e., 90+, the mortality rate increases steeply.

The same four grid searches were performed as for the Weibull distribution. In all four cases, the parameter α and β were searched up to a precision of $10^{-3}$. The parameter constellation with the largest median survival time (MST) was selected. The R code is provided in Section 5 of this document.

Optimal parameters were:

1. NICC: α = 6.305, β = 2.495 with values
   - Survival probability after 1 year: 0.9900
   - Survival probability after 20 years: 0.0531
   - Survival probability after 30 years: 0.0200
   - Median survival time: 6.61
2. NICC: α = 6.174, β = 2.907 with values
   - Survival probability after 1 year: 0.9950

- Survival probability after 20 years: 0.0318
- Survival probability after 30 years: 0.0100
- Median survival time: 6.174

3. Control: α = 3.710, β = 2.199 with values
   - Survival probability after 1 year: 0.947
   - Survival probability after 20 years: 0.0240
   - Survival probability after 30 years: 0.0100
   - Median survival time: 3.71
4. Control: α = 4.015, β = 2.285 with values
   - Survival probability after 1 year: 0.9600
   - Survival probability after 20 years: 0.0249
   - Survival probability after 30 years: 0.0100
   - Median survival time: 4.02

The formulae for the log-logistic distribution are:

- Survival probability for time past t: $P(T > t) = 1 - \left(1 + (t/\alpha)^{-\beta}\right)^{-1}$
- Median survival time: $\mathrm{MST} = \alpha$

##### *1.4 Simulation model for estimating cost per QALY*

In a simulation approach, the survival of the cohort, the QALYs and the direct cost were estimated by simulation.

1. Age- and sex-specific survival probabilities were taken from the survival data for the German population from the new states, see above.
2. QALYs were estimated from a linear regression with independent variables sex, age, cardiovascular disease, and treatment group. Regression estimates are provided in Supplementary Table 3.
3. Direct cost were estimated using quantile regression on the log plus 1 transformed cost data with independent variables sex, age, cardiovascular disease, treatment group, and the direct cost from the year before (log plus 1 transformed). Regression estimates are provided in Supplementary Table 3.
   For sensitivity analysis with the assumption of no cost difference between treatment groups, a second quantile regression was estimated on the log plus 1 transformed cost data with independent variables sex, age, cardiovascular disease, and the direct cost from the year before (log plus 1 transformed), without the independent variable treatment group.
4. EQ5D index values, thus QALY values were partly missing in the total cohort of 957 subjects. Similarly, cost data were only transferred for 923 patients from health insurance companies. Missing data were filled for the simulation model using a simple regression approach. Specifically, missing EQ5D values at baseline and direct cost were imputed by predictions from corresponding regression models using the complete cases. Similarly, missing EQ5D values at 6-month follow-up were imputed using the independent variables and the EQ5D

value at baseline. For the EQ5D values at 12 months follow-up both EQ5D values and baseline and at 6-month follow-up were used for imputation.

5. A cohort of respectively 1000 and 2000 patients was drawn with replacement from the original cohort of 957 patients. Per patient, the trajectory of survival was simulated using the age- and sex-specific survival probabilities. Direct cost and QALY were estimated for every simulated patient and every year using the regression models estimated under 2 and 3 with adjustment for sex, age, and cardiovascular disease. In one set of calculations, we assumed that all simulated patients were treated according to NICC. In the other set of calculations, we assumed that patients were treated according to SoC. This way we were able to perform a direct comparison of the treatment group with identical simulated cohorts. We replicated each model 1000 times and estimated the QALY difference between treatment groups per year, the cost differences per year, and the cost per QALY for year 1 as well as the cost per QALY for each year until median survival time in the NICC group.

#### 2. Health Economic Analysis Concept: Important Research Questions and Hypotheses

This concept was finalized on December 11, 2019. Data from health insurance companies were applied for after finalization of the health economic analysis concept.

##### *2.1 Overview of the research questions and hypotheses*

In this section, research questions and derived null hypotheses are verbally formulated. Statistical tests will be considered as alternative tests. Outcomes, their definition and measurement time points and time periods, respectively, are also listed. The statistical analysis approach is provided. Core of the NICC is the care center, a 24/7 call center. Patients of the NICC group additionally receive a computer tablet and, depending on their disease medical devices, such as a pulse oximeter. The design of the evaluation trial has been published [1]. Because the intervention is complex, several effects are expected in comparison with the standard of care (SoC) treatment. SoC is patient treatment according to the guidelines of the European Society for Cardiology (ESC).
Effects are expected in the following areas:

1. Inpatient treatment and inpatient treatment costs,
2. Outpatient treatment and outpatient treatment costs,
3. Prescription costs,
4. Total cost.

###### 2.1.1 Inpatient treatment and inpatient treatment costs

A lower number of hospitalizations is expected in the NICC group compared with SoC. The lower number of hospitalizations may especially result from a lower number of cardiovascular hospitalizations. The same argument holds true for the number of inpatient days. The number of cardiac decompensations should also be reduced by the NICC. This, in turn, should lead to a lower number of emergency hospitalizations and cardiovascular hospitalizations. The corresponding research questions are therefore formulated in Section 3 as questions with an advantage for the NICC group.

###### 2.1.2 Outpatient treatment and outpatient treatment costs

The Care Center as integral part acts as first contact for patients of the NICC group, and the Care Center has partly taken over the role of an "agony aunt". If patients feel well treated and accepted, it could be that the number of visits at an outpatient doctor, and in consequence costs, may be reduced. It could also be that patients feel better, which could result in fewer consultations with a neurologist or psychiatric working doctors or psychologists. The NICC could, however, just have the opposite effect. With the close care of the Care Center and the regularly displayed questionnaires on depression and anxiety, it could well be that relevant developments can be detected earlier. This early detection could lead to a higher number of visits at the corresponding specialized doctors in the short run, thus lead to higher costs in this area for NICC patients compared to SoC patients. Since effects could be in both directions, the research questions are formulated accordingly.

###### 2.1.3 Prescription costs

The closely monitored medical treatment could also lead to a change in costs for medicines. Maybe, the adherence increases per se so that the cost for medicines drops. It could also be that the optimized treatment by the NICC and by additional care givers reduces the cost for medicines. The effect could, however, also be in the opposite direction: with the closely monitored care it could well be that an additional need for treatment is identified, including medicines.

###### 2.1.4 Total costs

To get an overview of the total costs, inpatient and outpatient costs and costs for medicines will be considered.

##### *2.2 Methodological aspects*

Research questions are formulated below so that analyses first aim at a comparison of the NICC group with the SoC group.

The following two patient groups are therefore used in further analyses:

1. Patients of the NICC group,
2. Patients of the SoC group who are trial participants.

In the first step, the time point at the end of the 12 months observation period will be analyzed. To investigate whether structural differences exist between randomized treatment groups at the time of randomization, treatment groups will also be compared for the time period of the 12 months prior to randomization. If there is a treatment group difference at the end of the 12 month observation period, a pre-post comparison should also be performed within treatment group for investigating causality using the time period 12 months prior to randomization until 12 months post randomization.

The analysis set for this project is the full analysis set (FAS) of the evaluation study based on the intention to treat (ITT) principle. This data set consists of all patients who have received at least one treatment. Patients from the NICC group are included in the FAS if they have received at least one medical device in the trial, which is the tablet. Patients from the SoC group are included in the FAS if they have successfully completed the baseline investigation.

The type I error level is 5% two-sided for all hypotheses. Adjustments for multiple testing will not be done because independent research questions are primarily considered. Several sensitivity analyses will be conducted for the formulated research questions and primary statistical analyses. These include adjustments for possible confounders, specifically type of recruitment (inpatient or outpatient), age, sex, type of cardiovascular disease and disease severity. The cardiovascular diseases heart failure, atrial fibrillation and treatment-resistant hypertension will also be considered as subgroups.

Research questions are formulated below so that analyses first aim at a comparison of the NICC group with the SoC group.

Three different statistical approaches can be considered for most of the scientific questions formulated below:

1. Only the 12 months post randomization are considered for an outcome.
2. The difference of the endpoint values for the 12 months pre and post randomization are considered as outcome.
3. Endpoint is based on the 12 months post randomization, but an adjustment is made for the 12 month pre randomization.

As all analyses will be adjusted for the stratification variables cardiovascular disease and randomization inpatient/outpatient recruitment, all analyses described below will be regression analyses.

The use of the difference as described in 2. is meaningful if the outcome is continuous. The following regression approaches will be used:

- Linear regression for continuous outcome,
- Tobit regression for truncated continuous outcome,
- Poisson regression with a parameter to allow for over- or under-dispersion for count data; possibly negative binomial regression; in case of low number of events zero-inflated Poisson regression,
- Logistic regression for binary outcome; in case of low event rates Firth regression.

Wald tests and corresponding 95% confidence intervals will be estimated. Since cardiovascular disease has only three categories, i.e., atrial fibrillation, heart failure and resistant hypertension, and since recruitment had only the two access points inpatient and outpatient, adjustments for these stratification variables will be made in all regression analyses with fixed effects.

#### *2.3 Specific hypotheses*

##### 2.3.1 Inpatient area

Research question: Does NICC lead to a lower number of hospitalizations compared with SoC?

Outcomes:

a) Number of hospitalizations of a patient within 12 months post-trial inclusion.
b) Difference of number of hospitalizations of a patient within 12 months pre- and post-trial inclusion.
c) Number of hospitalizations of a patient within 12 months post-trial inclusion adjusted by the number of inpatient stays within 12 months pre-trial inclusion.

Null hypothesis: There is no difference in the number of hospitalizations between NICC and SoC.

Statistical analysis:

a) Poisson regression
b) Linear regression for difference of ln (x + 1).

c) Poisson regression.

…

Research question: Does NICC lead to a lower number of inpatient treatment days compared with SoC?

Outcomes:

a) Number of inpatient treatment days of a patient within 12 months post-trial inclusion.
b) Difference of number of inpatient treatment days of a patient within 12 months pre and post-trial inclusion.
c) Number of inpatient treatment days of a patient within 12 months post-trial inclusion adjusted by the number of inpatient treatment days within 12 months pre-trial inclusion.

Null hypothesis: There is no difference in the number of inpatient days between NICC and SoC.

Statistical analysis:

a) Poisson regression
b) Linear regression for difference of ln (x + 1).
c) Poisson regression.

…

Research question: Does NICC lead to lower inpatient treatment cost compared with SoC?

Outcomes:

a) Inpatient treatment cost of a patient within 12 months post-trial inclusion.
b) Difference of inpatient treatment cost of a patient within 12 months pre and post-trial inclusion.
c) Inpatient treatment cost of a patient within 12 months post-trial inclusion adjusted by the inpatient treatment cost within 12 months pre-trial inclusion.

Null hypothesis: There is no difference in the inpatient treatment cost between NICC and SoC.

Statistical analysis:

a) Linear regression
b) Linear regression
c) Linear regression

…

2.3.2 Outpatient area

…

Research question: Is there a difference in the outpatient treatment cost between NICC and SoC?

Outcomes:

a) Outpatient treatment cost of a patient within 12 months post-trial inclusion.

b) Difference of outpatient treatment cost of a patient within 12 months pre and post-trial inclusion.
c) Outpatient treatment cost of a patient within 12 months post-trial inclusion adjusted by the outpatient treatment cost within 12 months pre-trial inclusion.

Null hypothesis: There is no difference in the outpatient treatment cost between NICC and SoC.

Statistical analysis:

a) Linear regression
b) Linear regression
c) Linear regression

…

##### 2.3.3 Prescription costs

Research question: Are there differences in the costs for medicines between NICC and SoC?

Outcomes:

a) Cost for medicines of a patient within 12 months post-trial inclusion.
b) Difference of cost for medicines of a patient within 12 months pre and post-trial inclusion.
c) Cost for medicines of a patient within 12 months post-trial inclusion adjusted by the cost for medicines within 12 months pre-trial inclusion.

Null hypothesis: There is no difference in the cost for medicines between NICC and SoC.

Statistical analysis:

a) Linear regression
b) Linear regression
c) Linear regression

…

##### 2.3.4 Total cost

Research question: Does NICC lead to lower total treatment cost compared to SoC?

Outcomes:

a) Total treatment cost of a patient within 12 months post-trial inclusion.
b) Difference of total treatment cost of a patient within 12 months pre and post-trial inclusion.
c) Total treatment cost of a patient within 12 months post-trial inclusion adjusted by the total treatment cost within 12 months pre-trial inclusion.

Null hypothesis: There is no difference in the total treatment cost between NICC and SoC.

Statistical analysis:

a) Linear regression
b) Linear regression
c) Linear regression

#### 3. Supplementary Figures

**Supplementary Figure 1.** Frequency distribution of EQ-5D-5L questionnaire at baseline (grey), 6-month (red) and 1-year follow-up (orange) for complete cases. Left: NICC – integrated care concept; right: SoC – standard of care. Display per item.

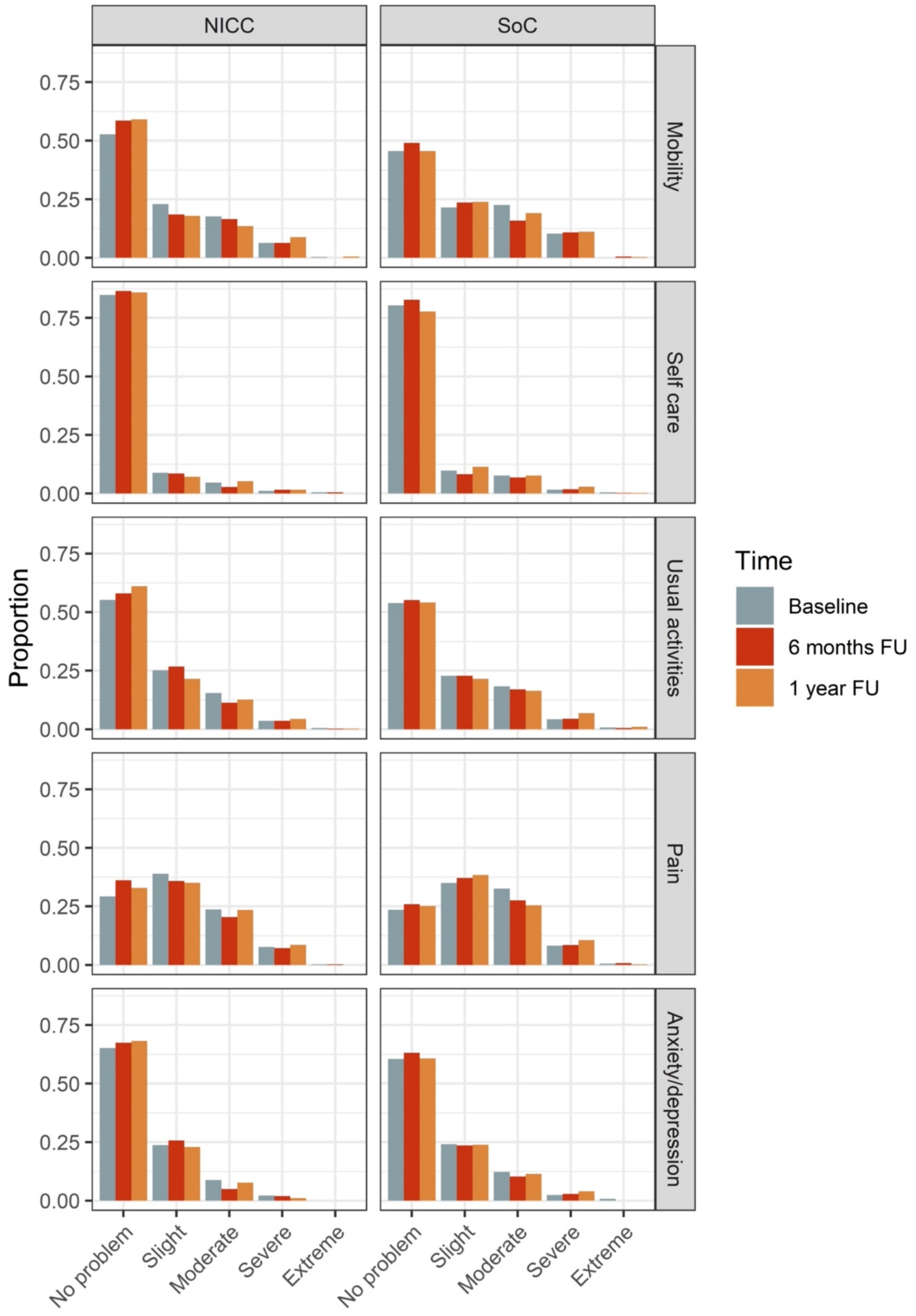

**Supplementary Figure 2.** Needle plot with relative frequencies for EQ5D index values during the course of the CardioCare MV trial by treatment group. Vertical dotted line represents mean values per treatment group and observation time point. NICC: integrated care concept; SoC: standard of care.

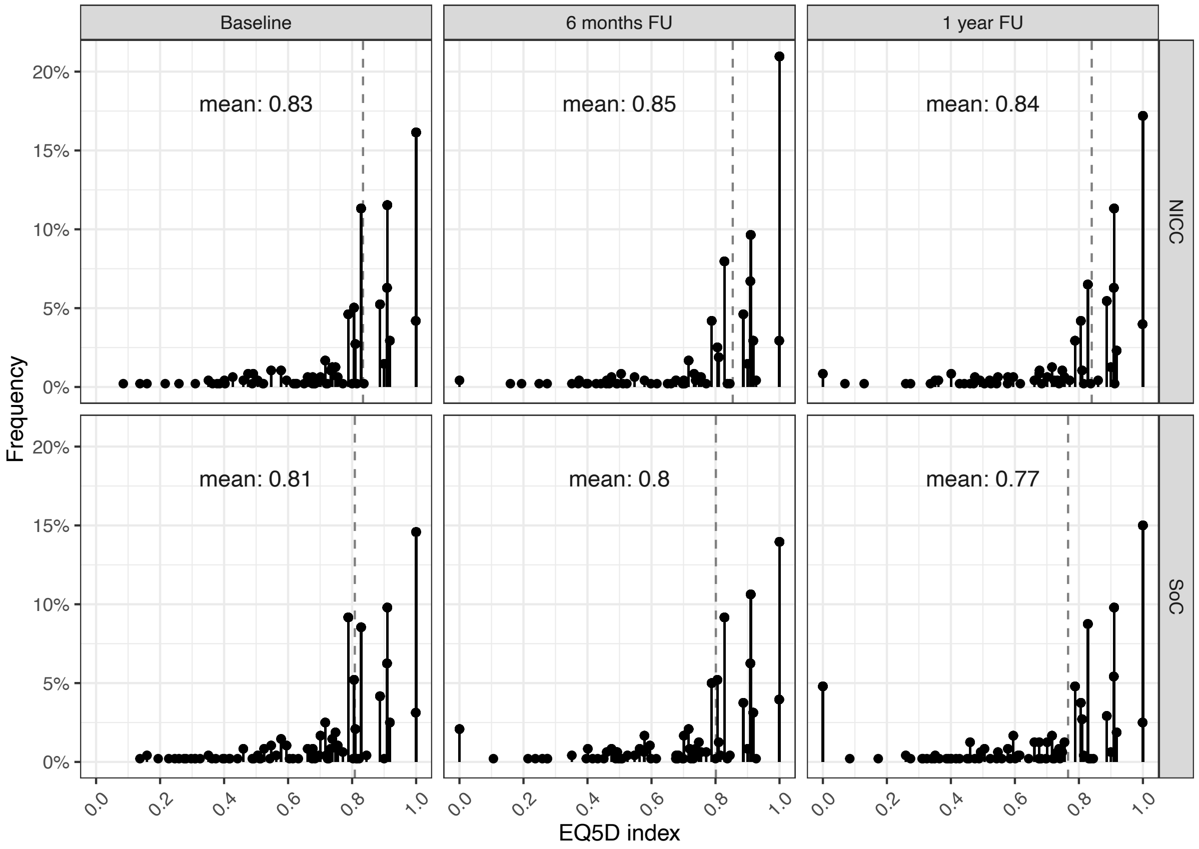

**Supplementary Figure 3.** Needle plot with relative frequencies for EQ5D VAS score during the course of the CardioCare MV trial by treatment group. Vertical dotted line represents mean values per treatment group and observation time point. NICC: integrated care concept; SoC: standard of care.

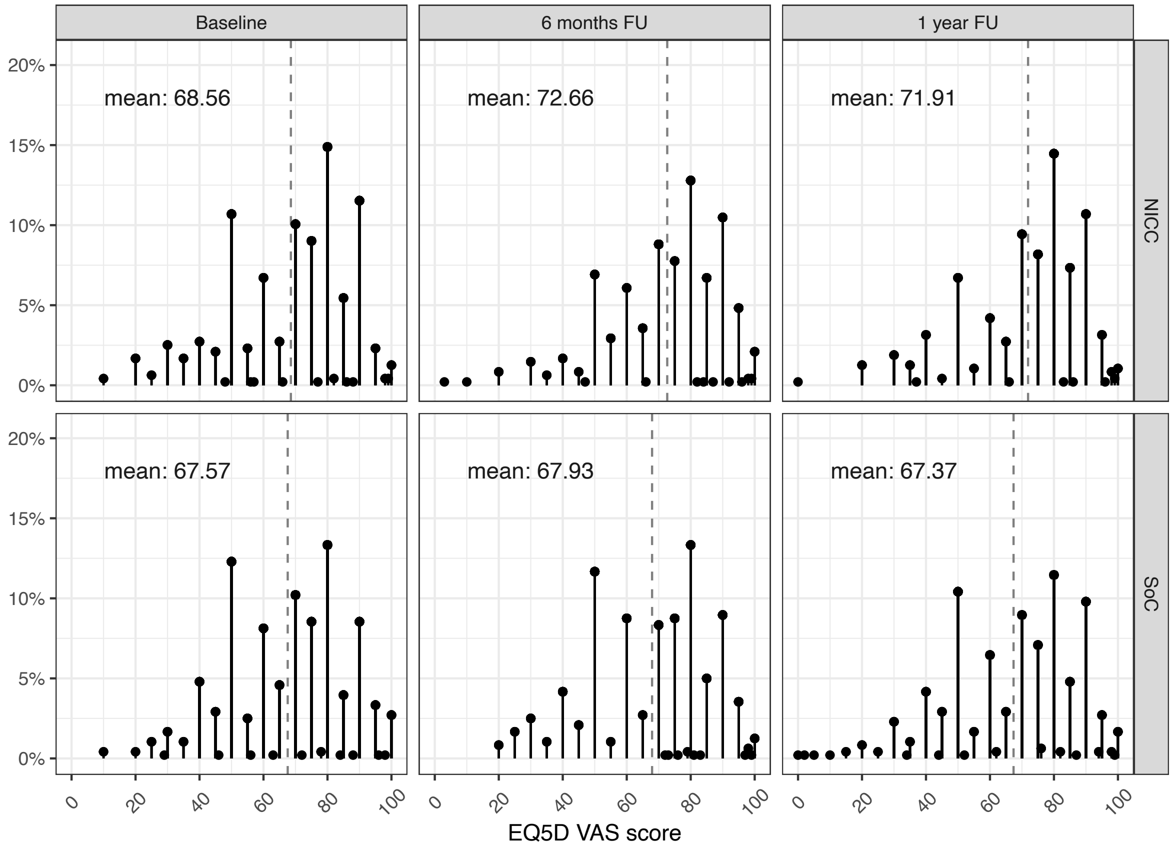

Supplementary Figure 3 shows that many patients frequently scored VAS values which can be divided by 10. Very few subjects scored values > 95 and < 5. There were neither floor nor ceiling effects for the VAS. For the 1-year follow-up, this also depicts that fewer patients in the NICC group scored a value of 50 compared with patients in the SoC group. Concurrently, more patients in the NICC group scored a EQ5D VAS score of 80 at the 1-year follow-up.

**Supplementary Figure 4.** Cost effectiveness analysis from the simulated cohort data. Incremental cost per QALY is displayed per year for direct cost and the case of 1000 patients and 2000 patients, respectively, served by the care center. Incremental cost was averaged up to the year displayed. The discount rate was 0%.

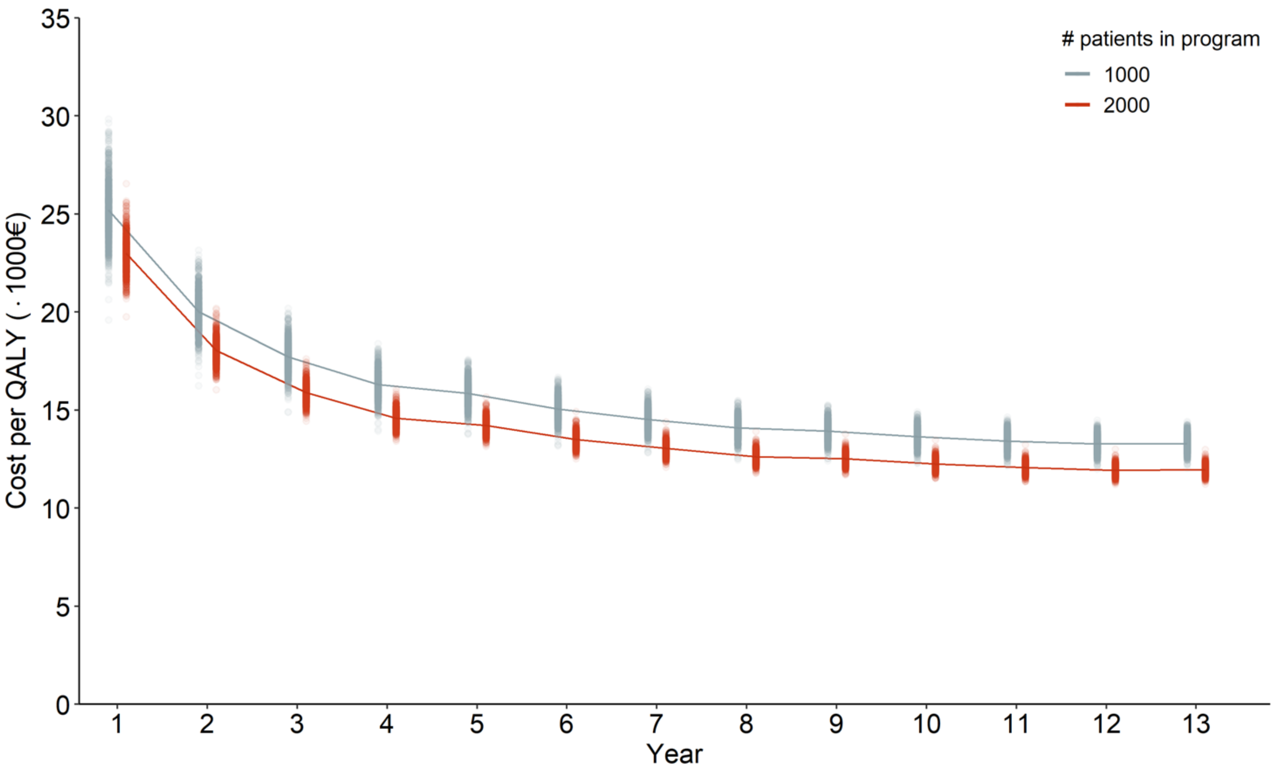

**Supplementary Figure 5.** Cost effectiveness analysis from the simulated cohort data. Incremental cost per QALY is displayed per year for direct cost and the case of 1000 patients and 2000 patients, respectively, served by the care center. Incremental cost was averaged up to the year displayed. The discount rate was 5%.

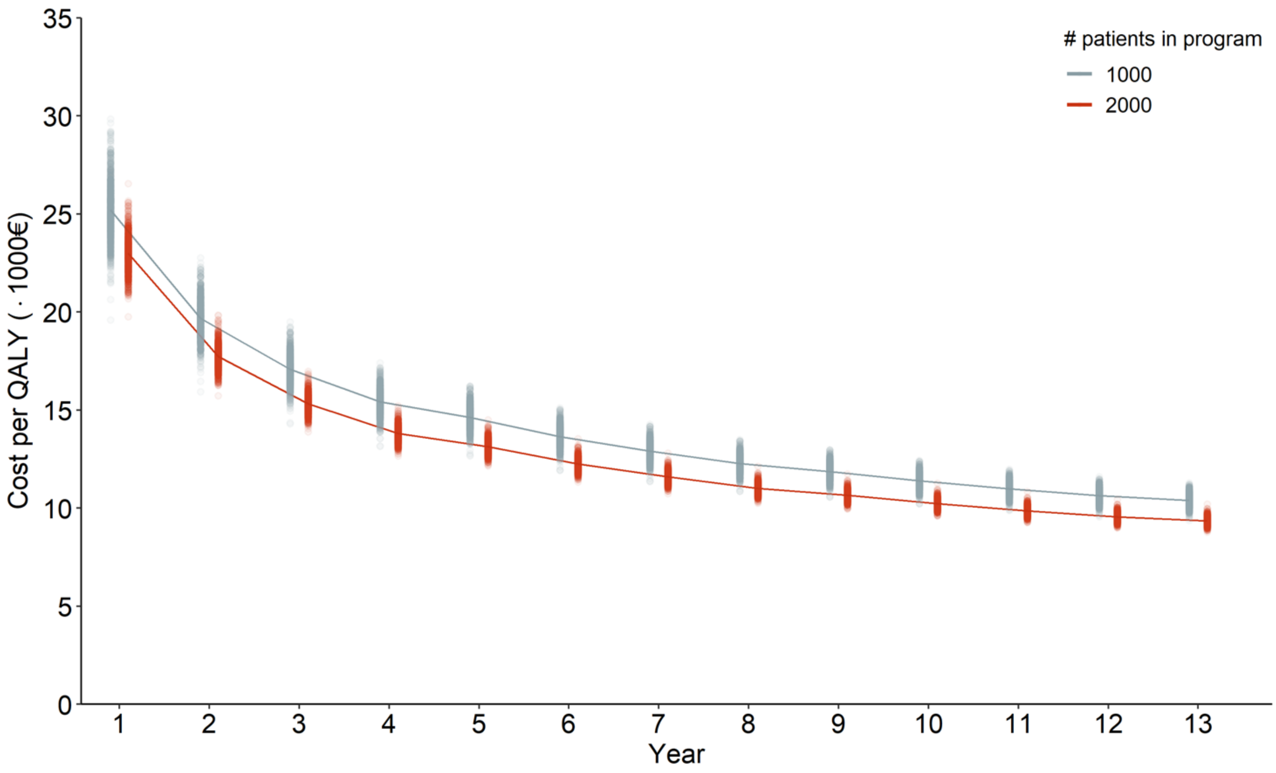

#### 4. Supplementary Tables

**Supplementary Table 1.** Cost utility and quality of life analysis in the CardioCare MV trial. Results were obtained from regression with adjustments for stratification variables. Estimates, 95% confidence intervals (95% CI) and p-values are provided.

| Outcome variable | Independent variable | Primary analysis: multiple imputation | | | Sensitivity analysis: complete case | | |
|---|---|---|---|---|---|---|---|
| | | Estimate | 95% CI | p-value | Estimate | 95% CI | p-value |
| QALY | Intercept | 0.794 | 0.739 – 0.851 | < 0.001 | 0.808 | 0.731 – 0.885 | < 0.001 |
| | SoC | -0.031 | -0.050 – -0.012 | 0.001 | -0.050 | -0.073 – -0.026 | < 0.001 |
| | AF | 0.032 | 0.008 – 0.055 | 0.008 | 0.054 | 0.024 – 0.083 | < 0.001 |
| | TRH | 0.012 | -0.011 – 0.034 | 0.317 | 0.036 | 0.008 – 0.065 | 0.012 |
| | Outpatient | 0.041 | -0.014 – 0.096 | 0.143 | 0.066 | -0.001 – 0.133 | 0.054 |
| VAS-AL | Intercept | 0.626 | 0.563 – 0.688 | < 0.001 | 0.664 | 0.586 – 0.741 | <0.001 |
| | SoC | -0.031 | -0.053 – -0.010 | 0.004 | -0.034 | -0.057 – -0.011 | 0.004 |
| | AF | 0.050 | 0.024 – 0.076 | < 0.001 | 0.060 | 0.031 – 0.089 | < 0.001 |
| | TRH | 0.036 | 0.011 – 0.062 | 0.005 | 0.036 | 0.008 – 0.063 | 0.011 |
| | Outpatient | 0.061 | -0.001– 0.122 | 0.053 | 0.063 | -0.005 – 0.130 | 0.068 |
| EQ5D index at 1 year FU | Intercept | 0.797 | 0.730 – 0.865 | < 0.001 | 0.806 | 0.705 – 0.906 | < 0.001 |
| | SoC | -0.034 | -0.056 – -0.011 | 0.003 | -0.072 | -0.103 – -0.042 | < 0.001 |
| | AF | 0.042 | 0.014 – 0.069 | 0.003 | 0.060 | 0.022 – 0.097 | 0.002 |
| | TRH | 0.006 | -0.021 – 0.033 | 0.655 | 0.042 | 0.006 – 0.079 | 0.025 |
| | Outpatient | 0.032 | -0.034 – 0.099 | 0.338 | 0.079 | -0.008 – 0.165 | 0.077 |
| VAS at 1 year FU | Intercept | 60.702 | 52.738 – 68.665 | < 0.001 | 65.954 | 56.834 – 75.075 | < 0.001 |
| | SoC | -3.686 | -6.331 – -1.042 | 0.007 | -4.402 | -7.090 – -1.713 | 0.001 |
| | AF | 4.460 | 1.232 – 7.688 | 0.007 | 5.412 | 2.102 – 8.721 | 0.001 |
| | TRH | 2.687 | -0.423 – 5.798 | 0.091 | 2.856 | -0.336 – 6.047 | 0.080 |
| | Outpatient | 8.256 | 0.488 – 16.025 | 0.038 | 8.124 | 0.130 – 16.118 | 0.047 |

AF: atrial fibrillation; FU: follow-up; QALY: quality-adjusted life-year; SoC: standard of care; TRH: treatment-resistant hypertension; VAS: visual analogue scale; VAS-AL: visual analogue scale adjusted life-years. Deceased patients were scored as 0.

**Supplementary Table 2.** Cost utility and quality of life outcomes in the CardioCare MV trial. Means and 95% confidence intervals are provided for all patients (All; n = 957) and by their primary diagnosis, i.e., atrial fibrillation (AF; n = 265), heart failure (HF; n = 406) or treatment-resistant hypertension (TRH; n = 286). The p-value is from the two-sample t-test.

| Variable | Group | NICC | SoC | p |
|---|---|---|---|---|
| QALY | All | 0.850 (0.834 – 0.865) | 0.799 (0.780 – 0.817) | < 0.001 |
| | AF | 0.878 (0.853 – 0.903) | 0.822 (0.790 – 0.853) | 0.006 |
| | HF | 0.832 (0.806 – 0.859) | 0.765 (0.733 – 0.796) | 0.001 |
| | TRH | 0.845 (0.817 – 0.874) | 0.824 (0.795 – 0.853) | 0.297 |
| VAS-AL | All | 0.719 (0.703 – 0.735) | 0.684 (0.667 – 0.700) | 0.003 |
| | AF | 0.753 (0.724 – 0.782) | 0.710 (0.681 – 0.739) | 0.039 |
| | HF | 0.693 (0.666 – 0.719) | 0.654 (0.626 – 0.683) | 0.052 |
| | TRH | 0.721 (0.691 – 0.751) | 0.698 (0.669 – 0.727) | 0.278 |
| EQ5D index | | | | |
| at baseline | All | 0.834 (0.819 – 0.849) | 0.808 (0.792 – 0.824) | 0.023 |
| | AF | 0.849 (0.818 – 0.880) | 0.834 (0.807 – 0.861) | 0.466 |
| | HF | 0.818 (0.795 – 0.842) | 0.784 (0.758 – 0.811) | 0.065 |
| | TRH | 0.841 (0.814 – 0.867) | 0.818 (0.790 – 0.846) | 0.246 |
| at 6 months FU | All | 0.854 (0.837 – 0.871) | 0.801 (0.782 – 0.821) | < 0.001 |
| | AF | 0.878 (0.848 – 0.908) | 0.818 (0.783 – 0.854) | 0.013 |
| | HF | 0.839 (0.812 – 0.866) | 0.772 (0.738 – 0.805) | 0.003 |
| | TRH | 0.851 (0.820 – 0.882) | 0.825 (0.794 – 0.856) | 0.253 |
| at 1 year FU | All | 0.840 (0.822 – 0.859) | 0.766 (0.742 – 0.790) | < 0.001 |
| | AF | 0.881 (0.857 – 0.905) | 0.781 (0.733 – 0.830) | < 0.001 |
| | HF | 0.817 (0.784 – 0.851) | 0.731 (0.690 – 0.771) | 0.002 |
| | TRH | 0.831 (0.796 – 0.865) | 0.801 (0.764 – 0.837) | 0.242 |
| VAS | | | | |
| at baseline | All | 68.56 (66.76 – 70.36) | 67.57 (65.84 –69.30) | 0.436 |
| | AF | 71.88 (68.40 – 75.37) | 71.62 (68.83 – 74.41) | 0.907 |
| | HF | 64.564 (61.74 – 67.39) | 63.74 (60.83 – 66.65) | 0.689 |
| | TRH | 70.861 (67.79 – 73.93) | 69.08 (66.03 – 72.13) | 0.417 |
| at 6 months FU | All | 72.67 (70.88 – 74.45) | 67.93 (66.11 – 69.74) | < 0.001 |
| | AF | 75.82 (72.50 – 79.14) | 70.09 (66.96 – 73.210) | 0.013 |
| | HF | 70.15 (67.36 – 72.94) | 65.18 (62.15 – 68.20) | 0.019 |
| | TRH | 72.97 (69.73 – 76.20) | 69.64 (66.42 – 72.86) | 0.151 |
| at 1 year FU | All | 71.98 (70.05 – 73.77) | 67.37 (65.41 – 69.34) | 0.001 |
| | AF | 74.51 (71.57 – 77.46) | 70.37 (66.92 – 73.81) | 0.071 |
| | HF | 70.86 (67.78 – 73.93) | 63.82 (60.64 – 67.00) | 0.002 |
| | TRH | 70.86 (67.25 – 74.46) | 69.50 (65.97 – 73.02) | 0.594 |

FU: follow-up; QALY: Quality-adjusted life-year; VAS-AL: visual analogue scale adjusted life-years.

**Supplementary Table 3.** Survival rates, hazard rates, hazard ratios and median survival times by publication.

| First author | Group | NYHA | Age (years) | Follow-up time | Survival rate | Hazard rate λ | Median survival time (years) |
|---|---|---|---|---|---|---|---|
| **Drozd** [24] | Stable chronic heart failure | | 69.6 ± 12.5 | 5 years | | | 6.6 |
| | Heart failure male | | | 10 years | 0.658 | 0.0418 | 16.576 |
| | Heart failure female | | | 10 years | 0.671 | 0.0399 | 17.376 |
| | Healthy | | | 10 years | 0.714 | 0.0337 | 20.576 |
| **Hindricks** [22] | Heart failure, telemonitoring group | II: 45%, III: 55% | 65.3 ± 9.3 | 1 year | 0.964 | 0.0346 | 20.038 |
| | Heart failure, control group | II: 41%, III: 59% | 65.8 ± 9.6 | 1 year | 0.913 | 0.0910 | 7.6154 |
| | | | | | *Hazard ratio* | *0.36 (0.17 – 0.74)* | |
| **Kotooka** [25] | Hospitalized heart failure | II: 79%, III: 21% | 66.3 ± 13.4 | 1 year | 0.921 | 0.0801 | 8.6507 |
| | | | | 2 years | 0.835 | 0.0902 | 7.6878 |
| **Koehler** [26] | Heart failure, telemonitoring | II: 50%, III: 50% | 66.9 ± 10.7 | 1 year | 0.916 | 0.0801 | 7.9 |
| | Heart failure, usual care | | | 1 year | 0.913 | 0.0902 | 7.6154 |
| **Koehler** [23] | Heart failure, telemonitoring | I: 0%, II: 52%, III: 47%, IV: 0% | 70 ± 11 | 1 year | 0.921 | 0.0823 | 8.4227 |
| | Heart failure, usual care | I: 1%, II: 51%, III: 47%, IV: 0% | 70 ± 10 | 1 year | 0.887 | 0.1199 | 5.7805 |
| | | | | | *Hazard ratio* | *0.70 (0.50 – 0.96)* | |
| **Öner** [3] | Heart failure 423 (44.2%) | I: 3%, II: 67%, III: 29%, IV: 1% | | | | | |
| | Atrial fibrillation 440 (45.5%) | EHRA I: 5%, II: 84%, III: 11%, IV: 1% | | | | | |
| | Usual care | | 71.1 ± 10.8 | 1 year | 0.947 | 0.0545 | 12.729 |
| | Telemonitoring | | 71.3 ± 10.4 | 1 year | 0.990 | 0.0101 | 68.968 |
| | | | | | *Hazard ratio* | *0.1942 (0.07 - 0.5618)* | |
| | Usual care | | | | | 0.0545 | 12.729 |
| | Telemonitoring | *Obtained from upper bound confidence interval of hazard ratio* | | | | 0.0306 | 22.657 |
| **Störk** [27] | Heart failure | NYHA I | | 2 years | 0.844 | 0.0848 | 8.1738 |
| | | NYHA II | | | 0.831 | 0.0926 | 7.4884 |

| | | | | | | | |
|---|---|---|---|---|---|---|---|
| | | NYHA III | | | 0.692 | 0.1841 | 3.7654 |
| | | NYHA IV | | | 0.467 | 0.3807 | 1.8207 |
| **Taylor** [28] | No heart failure | | 76.1 ± 10.4 | 5 years | 0.743 | 0.0594 | 11.667 |
| | | | | 10 years | 0.552 | 0.0594 | 11.665 |
| | | | | 15 years | 0.409 | 0.0596 | 11.629 |
| | Heart failure, not admitted to hospital | | 77.1 ± 10.6 | 5 years | 0.513 | 0.1335 | 5.1923 |
| | | | | 10 years | 0.289 | 0.1241 | 5.5839 |
| | | | | 15 years | 0.157 | 0.1234 | 5.6155 |

**Supplementary Table 4.** Estimates used in cost effectiveness analysis when simulated cohorts were used. Estimates were obtained from linear regression for QALY and from quantile regression for direct cost.

| Variable | QALY | | Direct cost | | | |
|---|---|---|---|---|---|---|
| | | | Allowing for cost difference | | Identical direct cost between NICC and SoC | |
| | Estimate | 95% CI | Estimate | 95% CI | Estimate | 95% CI |
| Intercept | 1.072 | 1.002 – 1.143 | 2.400 | 1.988 – 3.057 | 2.438 | 1.926 – 3.174 |
| SoC | -0.051 | -0.071 – -0.031 | 0.063 | -0.067 – 0.171 | – | – |
| AF | 0.052 | 0.027 – 0.076 | -0.053 | -0.158 – 0.089 | -0.069 | -0.162 – 0.098 |
| TRH | 0.034 | 0.010 – 0.058 | -0.156 | -0.283 – -0.014 | -0.166 | -0.291 – -0.022 |
| Sex | -0.040 | -0.061 – -0.019 | -0.067 | -0.167 – 0.082 | -0.058 | -0.160 – 0.069 |
| Age | -0.003 | -0.004 – -0.002 | 0.007 | 0.001 – 0.011 | 0.007 | 0.000 – 0.011 |
| Cost year before | | | 0.639 | 0.585 – 0.716 | 0.644 | 0.583 – 0.708 |

**Supplementary Table 5.** CHEERS 2022 Checklist.

| **Section** | **Guidance for Reporting** | **Reported in section** |
|---|---|---|
| Title | Identify the study as an economic evaluation and specify the interventions being compared. | Title |
| Abstract | Provide a structured summary that highlights context, key methods, results and alternative analyses. | Abstract |
| Background and objectives | Give the context for the study, the study question and its practical relevance for decision making in policy or practice. | Introduction |
| Health economic analysis plan | Indicate whether a health economic analysis plan was developed and where available. | Methods: subsection on cost data |
| Study population | Describe characteristics of the study population. | Table 1; Methods: subsection on trial population |
| Setting and location | Provide relevant contextual information that may influence findings. | Methods: subsection on trial population |
| Comparators | Describe the interventions or strategies being compared and why chosen. | Methods: subsection on intervention and control |
| Perspective | State the perspective(s) adopted by the study and why chosen. | Methods: subsection on cost effectiveness analysis |
| Time horizon | State the time horizon for the study and why appropriate. | Supplementary material 1 |
| Discount rate | Report the discount rate(s) and reason chosen. | Methods: subsection on cost effectiveness analysis |
| Selection of outcomes | Describe what outcomes were used as the measure(s) of benefit(s) and harm(s). | Methods: subsections on health-related quality of life, cost data, and cost effectiveness analysis |

| | | |
|---|---|---|
| Measurement of outcomes | Describe how outcomes used to capture benefit(s) and harm(s) were measured. | Methods: subsections on health-related quality of life, cost data, and cost effectiveness analysis |
| Valuation of outcomes | Describe the population and methods used to measure and value outcomes. | Methods: subsection on cost effectiveness analysis; supplementary material 1 |
| Measurement and valuation of resources and costs | Describe how costs were valued. | Methods: subsection on cost effectiveness analysis |
| Currency, price date, and conversion | Report the dates of the estimated resource quantities and unit costs, plus the currency and year of conversion. | Methods: subsection on cost effectiveness analysis |
| Rationale and description of model | If modelling is used, describe in detail and why used. Report if the model is publicly available and where it can be accessed. | Supplementary material 1 |
| Analytics and assumptions | Describe any methods for analysing or statistically transforming data, any extrapolation methods, and approaches for validating any model used. | Methods: subsection on statistical analysis; supplementary material 1 |
| Characterizing heterogeneity | Describe any methods used for estimating how the results of the study vary for sub-groups. | Methods: subsections on statistical analysis and cost-effectiveness analysis |
| Characterizing distributional effects | Describe how impacts are distributed across different individuals or adjustments made to reflect priority populations. | Methods: subsection on statistical analysis |
| Characterizing uncertainty | Describe methods to characterize any sources of uncertainty in the analysis. | Methods: subsections on statistical analysis |

| | | |
|---|---|---|
| Approach to engagement with patients and others affected by the study | Describe any approaches to engage patients or service recipients, the general public, communities, or stakeholders (e.g., clinicians or payers) in the design of the study. | Methods: subsection on cost data |
| Study parameters | Report all analytic inputs (e.g., values, ranges, references) including uncertainty or distributional assumptions. | Results; Supplementary Material |
| Summary of main results | Report the mean values for the main categories of costs and outcomes of interest and summarise them in the most appropriate overall measure. | Table 3 |
| Effect of uncertainty | Describe how uncertainty about analytic judgments, inputs, or projections affect findings. Report the effect of choice of discount rate and time horizon, if applicable. | Table 3 |
| Effect of engagement with patients and others affected by the study | Report on any difference patient/service recipient, general public, community, or stakeholder involvement made to the approach or findings of the study | Not applicable |
| Study findings, limitations, generalizability, and current knowledge | Report key findings, limitations, ethical or equity considerations not captured, and how these could impact patients, policy, or practice. | Discussion |

| | | |
|---|---|---|
| Source of funding | Describe how the study was funded and any role of the funder in the identification, design, conduct, and reporting of the analysis | Section on study funding |
| Conflicts of interest | Report authors conflicts of interest according to journal or International Committee of Medical Journal Editors requirements. | Section on conflicts of interest |

#### 5. R code

##### *5.1 R code to identify the best fitting Weibull distribution*

```
library(dplyr)
MST <- function( lambda, k) 1/lambda*(log(2)^(1/k))
fx <- function(x, lambda, k) (exp(-(lambda*x)^k))

grids <- expand.grid(
  lambda = seq(from = 0.01 , to = 1,   by = 0.0001),
  k      = seq(from = 0.01 , to = 5,       by = 0.001)
)

grids$F_1   = fx(1, grids$lambda, grids$k)
grids$F_20  = fx(20, grids$lambda, grids$k)
grids$F_30  = fx(30, grids$lambda, grids$k)
grids$MST = MST( grids$lambda, grids$k)

## NICC1
grids %>% filter(between(MST, 11, 20 ) & F_30 < 0.02 & F_1 < 0.99) %>%
arrange(desc(MST)) %>% head(20)

## control3
grids  %>%  filter(  F_30  <  0.01  &  between(F_1,  0.93,  0.947))
%>% arrange(desc(MST)) %>% head(20)

## NICC2
grids %>% filter(between(MST, 1, 20 ) & F_30 < 0.01 & F_1 < 0.995) %>%
arrange(desc(MST)) %>% head(20)

## control4
grids  %>%  filter(  F_30  <  0.01  &  between(F_1,  0.94,  0.96))  %>%
arrange(desc(MST)) %>% head(20)
```

##### *5.2 R code to identify the best fitting log-logistic distribution*

```
MST2 <- function( alpha) alpha
fx <- function(x, alpha, beta) (1 -(1 / (1 + ((x/alpha)^(-beta)))))

grids <- expand.grid(
  alpha = seq(from = 3 , to = 7,   by = 0.001),
  beta   = seq(from = 1 , to = 5,       by = 0.001)
)

grids$F_1   = fx(1, grids$alpha, grids$beta)
grids$F_20  = fx(20, grids$alpha, grids$beta)
grids$F_30  = fx(30, grids$alpha, grids$beta)
grids$MST2 = MST2( grids$alpha)
```

```
## NICC1
grids %>% filter(between(MST2, 1, 20 ) & F_30 < 0.02 & F_1 < 0.99) %>%
arrange(desc(MST2)) %>% head(20)

## control3
grids %>% filter( F_30 < 0.01 & between(F_1, 0.93, 0.947)) %>%
arrange(desc(MST2)) %>% head(20)

## NICC2
grids %>% filter(between(MST2, 1, 20 ) & F_30 < 0.01 & F_1 < 0.995)
%>% arrange(desc(MST2)) %>% head(20)

## control4
grids %>% filter( F_30 < 0.01 & between(F_1, 0.94, 0.96)) %>%
arrange(desc(MST2)) %>% head(20)
```

#### *5.3 Code used for the large imputation model*

```
# Code explanation: .3: baseline, .2; 6 mo follow-up (FU), .1: 12 mo
FU; _YN: yes/no
# Variables: Sexfac: Sex, rndRESULTfac: randomization result, rndCHD:
primary CVD, rndPATSTATUS: inpatient/outpatient, Urban: degree of
urbanization, BMI: body mass index, AF_YN: atrial fibrillation, AP_YN:
angina pectoris, HBP_YN: hypertension, HF_YN; heart failure, Hosp_YN:
hospitalization, NrPers: number of subjects in household, NrPers18:
number of subjects < 18 in household, Maritalfac: marital status,
Alcfac; any alcohol, Smokefac: smoking, Sport: sports, Racefac: race,
TVfac: TV in household, UMTSfac: internet in household,
HouseholdOther: living at own household YN, famMIfac: familial history
of MI, Countryfac: country of birth (Germany YN), EQ5D.q1.mobil.3:
question 1 (mobility of EG-5D) at baseline, EQ5D.q1.mobil.2,
EQ5D.q1.mobil.1, EQ5D.q2.selfcare.3, EQ5D.q2.selfcare.2,
EQ5D.q2.selfcare.1, EQ5D.q3.usualact.3, EQ5D.q3.usualact.2,
EQ5D.q3.usualact.1, EQ5D.q4.pain.3, EQ5D.q4.pain.2, EQ5D.q4.pain.1,
EQ5D.q5.anxdep.3, EQ5D.q5.anxdep.2, EQ5D.q5.anxdep.1,
EQ5D.score.index.3, EQ5D.score.index.2, EQ5D.score.index.1,
EQ5D.q6.VAS.3, EQ5D.q6.VAS.2, EQ5D.q6.VAS.1, HQOL.emot.score.3:
HeartQoL emotional score at baseline, HQOL.emot.score.2,
HQOL.emot.score.1, HQOL.phys.score.3: HeartQoL physical score at
baseline, HQOL.phys.score.2, HQOL.phys.score.1, HQOL.global.score.3:
HeartQoL global score at baseline, HQOL.global.score.2,
HQOL.global.score.1, GAD.score.3: GAD-7 score at baseline,
GAD.score.2, GAD.score.1, PHQ.score.3: PHQ-9 score at baseline,
PHQ.score.2, PHQ.score.1, WHO.score.3: WHO-5 score at baseline,
WHO.score.2, WHO.score.1, BMQ.genharm.3: BMQ general harm score at
baseline, BMQ.genharm.2, BMQ.genharm.1, BMQ.genoveruse.3: BMQ general
overuse score at baseline, BMQ.genoveruse.2, BMQ.genoveruse.1,
BMQ.specconc.3: BMQ specific concerns score at baseline,
BMQ.specconc.2, BMQ.specconc.1, BMQ.specnec.3: BMQ specific
necessities score at baseline, BMQ.specnec.2, BMQ.specnec.1,
```

MARS.score.3: MARS-5 score at baseline, MARS.score.2, MARS.score.1, PAM.score.3: PAM13-D score at baseline, PAM.score.2, PAM.score.1, WHR.3: waist hip ratio at baseline, WHR.2, WHR.1, CHA2DS2VASc.3: modified CHA2DS2VASc score at baseline, CHA2DS2VASc.2, CHA2DS2VASc.1, SSUK.score.detint.3: ISSS-8 detrimental interaction score at baseline, SSUK.score.detint.2, SSUK.score.detint.1, SSUK.score.possupp.3: ISSS-8 possible support score at baseline, SSUK.score.possupp.2, SSUK.score.possupp.1, numHospFreqpred: number of hospitalizations, primary1: primary endpoint 1, primary3: primary endpoint 3, Death: death within 1 year, CardDec_YN: cardiovascular death within 1 year, MACE: major acute cardiovascular event within 1 year, CVDeath2: cardiovascular death within 1 year, alternative definition

```
set.seed(4242, kind = "Mersenne-Twister")
Nimputations <- 3 # 100
Iterations <- 10 # 10

# dry imputation run for modifying predictor matrix in next step
dry.imp <- mice(data=df.impute.wide %>% select(Sexfac, Age, rndRESULTfac, rndCHD, rndPATSTATUS, Urban, BMI.3, BMI.2, BMI.1, AF_YN, AP_YN, HBP_YN, HF_YN, Hosp_YN, NrPers, NrPers18, Maritalfac, Alcfac, Smokefac, Sport, Racefac, TVfac, UMTSfac, HouseholdOther, famMIfac, Countryfac, EQ5D.q1.mobil.3, EQ5D.q1.mobil.2, EQ5D.q1.mobil.1, EQ5D.q2.selfcare.3, EQ5D.q2.selfcare.2, EQ5D.q2.selfcare.1, EQ5D.q3.usualact.3, EQ5D.q3.usualact.2, EQ5D.q3.usualact.1, EQ5D.q4.pain.3, EQ5D.q4.pain.2, EQ5D.q4.pain.1, EQ5D.q5.anxdep.3, EQ5D.q5.anxdep.2, EQ5D.q5.anxdep.1, EQ5D.score.index.3, EQ5D.score.index.2, EQ5D.score.index.1, EQ5D.q6.VAS.3, EQ5D.q6.VAS.2, EQ5D.q6.VAS.1, HQOL.emot.score.3, HQOL.emot.score.2, HQOL.emot.score.1, HQOL.phys.score.3, HQOL.phys.score.2, HQOL.phys.score.1, HQOL.global.score.3, HQOL.global.score.2, HQOL.global.score.1, GAD.score.3, GAD.score.2, GAD.score.1, PHQ.score.3, PHQ.score.2, PHQ.score.1, WHO.score.3, WHO.score.2, WHO.score.1, BMQ.genharm.3, BMQ.genharm.2, BMQ.genharm.1, BMQ.genoveruse.3, BMQ.genoveruse.2, BMQ.genoveruse.1, BMQ.specconc.3, BMQ.specconc.2, BMQ.specconc.1, BMQ.specnec.3, BMQ.specnec.2, BMQ.specnec.1, MARS.score.3, MARS.score.2, MARS.score.1, PAM.score.3, PAM.score.2, PAM.score.1, WHR.3, WHR.2, WHR.1, CHA2DS2VASc.3, CHA2DS2VASc.2, CHA2DS2VASc.1, SSUK.score.detint.3, SSUK.score.detint.2, SSUK.score.detint.1, SSUK.score.possupp.3, SSUK.score.possupp.2, SSUK.score.possupp.1, numHospFreqpred, primary1, primary3, Death, CardDec_YN, MACE, CVDeath2), maxit = 0, printFlag = F)

# Avoid multicollinearity in predictor matrix during imputaiton
predictormat <- dry.imp$predictorMatrix
predictormat[96,97:102] <- 0
predictormat[97,c(96, 98:102)] <- 0
predictormat[98,c(96, 97, 99, 100, 101, 102)] <- 0
```

```
predictormat[99,c(96:98, 100, 101, 102)] <- 0
predictormat[100,c(96:99, 101, 102)] <- 0
predictormat[101,c(96:100, 102)] <- 0
predictormat[102,96:101] <- 0

# Actual imputation run
imputation.model.1 <- parlmice(data = df.impute.wide %>% select(
Sexfac, Age, rndRESULTfac, rndCHD, rndPATSTATUS, Urban, BMI.3, BMI.2,
BMI.1, AF_YN, AP_YN, HBP_YN, HF_YN, Hosp_YN, NrPers, NrPers18,
Maritalfac, Alcfac, Smokefac, Sport, Racefac, TVfac, UMTSfac,
HouseholdOther, famMIfac, Countryfac, EQ5D.q1.mobil.3,
EQ5D.q1.mobil.2, EQ5D.q1.mobil.1, EQ5D.q2.selfcare.3,
EQ5D.q2.selfcare.2, EQ5D.q2.selfcare.1, EQ5D.q3.usualact.3,
EQ5D.q3.usualact.2, EQ5D.q3.usualact.1, EQ5D.q4.pain.3,
EQ5D.q4.pain.2, EQ5D.q4.pain.1, EQ5D.q5.anxdep.3, EQ5D.q5.anxdep.2,
EQ5D.q5.anxdep.1, EQ5D.score.index.3, EQ5D.score.index.2,
EQ5D.score.index.1, EQ5D.q6.VAS.3, EQ5D.q6.VAS.2, EQ5D.q6.VAS.1,
HQOL.emot.score.3, HQOL.emot.score.2, HQOL.emot.score.1,
HQOL.phys.score.3, HQOL.phys.score.2, HQOL.phys.score.1,
HQOL.global.score.3, HQOL.global.score.2, HQOL.global.score.1,
GAD.score.3, GAD.score.2, GAD.score.1, PHQ.score.3, PHQ.score.2,
PHQ.score.1, WHO.score.3, WHO.score.2, WHO.score.1, BMQ.genharm.3,
BMQ.genharm.2, BMQ.genharm.1, BMQ.genoveruse.3, BMQ.genoveruse.2,
BMQ.genoveruse.1, BMQ.specconc.3, BMQ.specconc.2, BMQ.specconc.1,
BMQ.specnec.3, BMQ.specnec.2, BMQ.specnec.1, MARS.score.3,
MARS.score.2, MARS.score.1, PAM.score.3, PAM.score.2, PAM.score.1,
WHR.3, WHR.2, WHR.1, CHA2DS2VASc.3, CHA2DS2VASc.2, CHA2DS2VASc.1,
SSUK.score.detint.3, SSUK.score.detint.2, SSUK.score.detint.1,
SSUK.score.possupp.3, SSUK.score.possupp.2, SSUK.score.possupp.1,
numHospFreqpred, primary1, primary3, Death, CardDec_YN, MACE,
CVDeath2) , maxit = Iterations, method = "pmm", predictorMatrix =
predictormat, printFlag = T, n.core = 12, n.imp.core = 1, cluster.seed
= 4242)
```

#### 5.4 Code for the dynamic model (simulation model)

```
library(data.table)
library(dplyr)
library(ggplot2)
library(parallel)
library(quantreg)
library(readxl)
library(tidyr)

######### Function declaration #########

mutate_cond <-
  function(.data, condition, ..., envir = parent.frame()) {
    condition <- eval(substitute(condition), .data, envir)
    .data[condition,] <- .data[condition,] %>% mutate(...)
    .data
```

```
  }

simulate_data <- function(data, directcosts.model, QALY.model, devices.cost,
mortality, mortality.mat, group_name){

  n.samples <- nrow(data)
  max.years <- ncol(devices.cost)
  data$rndRESULTfac <- group_name

  survival <- matrix(FALSE, nrow = n.samples, ncol = max.years)
  directcosts.mat <- matrix(0, nrow = n.samples, ncol = max.years)
  QALY.mat <- matrix(0, nrow = n.samples, ncol = max.years)
  if(tolower(group_name)=="nicc"){
    devices.mat <- matrix(0, nrow = n.samples, ncol = max.years)
  }

  data$alive <- TRUE
  data$PID <- 1:n.samples
  data <- data %>% mutate(directcost_Y_before = directcost_Y_after)

  group_fac <- 1 + 0.78*(tolower(group_name) == "soc")
  year <- 1
  condition_check <- TRUE

  while(condition_check){

    if(tolower(group_name)=="nicc"){
      # 1.a Calculate device costs
      devices.mat[data$PID,                        year]                     <-
devices.cost[cbind(as.integer(data$rndCHD), year)]
    }

    data <- data %>% mutate(EQ5D.QALY = predict(QALY.model,
            newdata = .), directcost_Y_after =
            exp(predict(directcosts.model, newdata = .)) - 1)

    directcosts.mat[data$PID, year] <- data$directcost_Y_after
    QALY.mat[data$PID, year] <- data$EQ5D.QALY

    mortality.year                 <-                  sapply(1:nrow(data),
function(n)mortality[[as.integer(data$Sexfac[n])+1]][min(100,
data$Age.round[n])])
    status <- mortality.year*group_fac < mortality.mat[data$PID, year]

    survival[data$PID, year] <- status

    data <- data %>% filter(status) %>% mutate(Age = Age + 1,
  Age.round = Age.round + 1, directcost_Y_before = directcost_Y_after)
    condition_check <- nrow(data) > 0
    year <- year + 1
  }

  QALY.mat[!survival] <- 0
```

```
  results <- list(survival = survival,  directcosts = directcosts.mat, QALY
= QALY.mat)

  if(tolower(group_name)=="nicc"){
    license.cost <- 600
    software.costs <- survival*license.cost
    cc.costs <- colSums(software.costs)
    if(n.samples == 1000){
      cc.costs[cc.costs > 0] <- cc.costs[cc.costs > 0] + 1e5 + 3.68e5 + 5e5
    }else if(n.samples == 2000){
      cc.costs[cc.costs > 0] <- cc.costs[cc.costs > 0] + 1.5e5 + 4.8e5 + 1e6
    }

    results$carecenter <- cc.costs
    results$devices <- devices.mat
  }
  return(results)
}

get_simulation_results <-
  function(data,
           directcosts.model,
           QALY.model,
           mortality,
           n.samples = 1000,
           plot.figures = FALSE) {
    sampled_inds <-
      sample.int(nrow(data), size = n.samples, replace = TRUE)
    data <- data[sampled_inds, ]
    data$Age.round <- round(data$Age)

    max_age <- 101
    age_diff <- max_age - min(data$Age.round)

    bpm_cost <- c(85, numeric(5))
    t_cost <- c(290, numeric(3))
    po_cost <- c(125, numeric(3))
    s_cost <- c(60, numeric(10))

    trh.devices  <-  rep(bpm_cost,  length.out  =  age_diff)  +  rep(t_cost,
length.out = age_diff)
    af.devices <- trh.devices + rep(po_cost, length.out = age_diff)
    hf.devices <- af.devices + rep(s_cost, length.out = age_diff)

    devices.cost <- rbind(hf.devices, af.devices, trh.devices)
    rownames(devices.cost) <- c("HF", "AF", "TRH")

    mortality.mat <-
      matrix(runif(age_diff * n.samples),
             nrow = n.samples,
             ncol = age_diff)

    SoC.results <-
      simulate_data(data, directcosts.model, QALY.model,
```

```
                  devices.cost, mortality, mortality.mat, "SoC")
    NICC.results <-
      simulate_data(data, directcosts.model, QALY.model,
        devices.cost, mortality, mortality.mat, "NICC")

    SoC.survival.median <-
      which.min(colSums(SoC.results$survival) > round(.5 * n.samples))
    SoC.costs.yearly <-  colSums(SoC.results$directcosts)
    SoC.costs.year1 <- SoC.costs.yearly[1]

    NICC.survival.median <-
      which.min(colSums(NICC.results$survival) > round(.5 * n.samples))

    NICC.costs.yearly <-
      colSums(NICC.results$directcosts)    +    NICC.results$carecenter    +
colSums(NICC.results$devices)
    NICC.year1.cost <-
      NICC.results$carecenter[1] + sum(NICC.results$devices[, 1])

    survival.max.min <-
      min(which.min(colSums(NICC.results$survival)),
which.min(colSums(SoC.results$survival)))
    survival.median.min <-
      min(SoC.survival.median, NICC.survival.median)

    df.results <-
      data.frame(
        year = 1:age_diff,
        SoC.survival = colSums(SoC.results$survival) / n.samples,
        NICC.survival = colSums(NICC.results$survival) / n.samples,
        NICC.QALY = colSums(NICC.results$QALY, na.rm = TRUE),
        SoC.QALY = colSums(SoC.results$QALY, na.rm = TRUE),
        NICC.costs = NICC.costs.yearly,
        SoC.costs = SoC.costs.yearly
      ) %>%
      mutate(QALY.diff = NICC.QALY - SoC.QALY,
             costs.diff = NICC.costs - SoC.costs) %>%
      mutate(
        QALY.cost = costs.diff / QALY.diff,
        QALY.diff.cum = cumsum(QALY.diff),
        NICC.costs.cum = cumsum(NICC.costs),
        SoC.costs.cum = cumsum(SoC.costs)
      ) %>%
      mutate(QALY.diff.cum.cost   =   (NICC.costs.cum   -   SoC.costs.cum)   /
QALY.diff.cum) %>%
      filter(pmax(SoC.survival, pmax(NICC.survival)) > 0)

    if (plot.figures) {
      extract.df <- function(variable.string) {
        SoC.string <- paste0("SoC.", variable.string)
        NICC.string <- paste0("NICC.", variable.string)
        df <-
          df.results %>%
```

```
          select(year, SoC.string, NICC.string) %>%
          rename(SoC = SoC.string, NICC = NICC.string) %>%
          gather(key = "group", value = variable.string,-year) %>%
          rename()
        return(df)
      }
      year.lim <- survival.median.min

      p.survival <- ggplot(data = extract.df("survival"),
                    aes(x = year, y = variable.string))
      p.survival + geom_line(aes(color = group, linetype = group)) +
                    xlim(1, year.lim) + ylab("Survival rate")

      p.QALY <- ggplot(data = extract.df("QALY"),
                aes(x = year, y = variable.string))
      p.QALY + geom_line(aes(color = group, linetype = group)) +
                xlim(1, year.lim) + ylab("QALY")

      p.costs <- ggplot(data = extract.df("costs"),
                 aes(x = year, y = variable.string))
      p.costs + geom_line(aes(color = group, linetype = group)) +
                 xlim(1, year.lim) + ylab("costs")

      p.costs.cum <- ggplot(data = extract.df("costs.cum"),
                 aes(x = year, y = variable.string))
      p.costs.cum + geom_line(aes(color = group, linetype = group)) +
                 xlim(1, year.lim) + ylab("costs.cum")

      p.QALY.cost <- ggplot(data = df.results, aes(x = year))
      p.QALY.cost + geom_line(aes(y = QALY.cost)) + xlim(1, year.lim) +
                 ylim(0, max(df.results$QALY.cost[1:year.lim]))

      p.QALY.diff.cum.cost <- ggplot(data = df.results, aes(x = year))
      p.QALY.diff.cum.cost + geom_line(aes(y = QALY.diff.cum.cost)) +
                 xlim(1, year.lim) +
                 ylim(0, max(df.results$QALY.diff.cum.cost[1:year.lim]))
    }

    return(df.results)
  }

######### Load necessary data #########

HE_all <- as.data.table(readRDS("HE_all.Rda"))
costanalysis <- as.data.table(readRDS("costanalysis.Rds"))
mortality <- readxl::read_xlsx(
  'Mortality_EastGermany_2021_11_26.xlsx', sheet = "Original"
)

######### Script #########

df.base <- HE_all %>%
  select(Patient_Number, PID, Visit_Definition_Id, Sexfac, Age, rndCHD,
         rndPATSTATUS, Death) %>% dplyr::filter(Visit_Definition_Id == 3)
```

```
df.base <- costanalysis %>%
  select(PID, directcost_Y_after, directcost_Y_before, rndRESULTfac,
    EQ5D.QALY, EQ5D.score.index.3, EQ5D.score.index.2, EQ5D.score.index.1)
    %>%
  right_join(., df.base) %>%
  relocate(Patient_Number, PID, Sexfac, Age, Death, rndCHD, rndRESULTfac,
    rndPATSTATUS, EQ5D.QALY, EQ5D.score.index.3, EQ5D.score.index.2,
    EQ5D.score.index.1, directcost_Y_after, directcost_Y_before)

df.base <- df.base %>%
  mutate(
    directcost_Y_after.isna = is.na(directcost_Y_after),
    directcost_Y_before.isna  = is.na(directcost_Y_before)
  )

set.seed(4242, kind = "Mersenne-Twister")
Nimputations <- 100
Iterations <- 10

score.model.3 <- df.base %>%
  filter(!is.na(EQ5D.score.index.3)) %>%
  lm(EQ5D.score.index.3 ~ Sexfac + Age + rndCHD + rndRESULTfac, data = .)

df.base <- df.base %>%
  mutate_cond(is.na(EQ5D.score.index.3),
          EQ5D.score.index.3 = predict(score.model.3, newdata = (df.base %>%
filter(
is.na(EQ5D.score.index.3)
                                                                  ))))

score.model.2 <- df.base %>%
  filter(!is.na(EQ5D.score.index.2)) %>%
  lm(EQ5D.score.index.2 ~ EQ5D.score.index.3 + Sexfac + Age + rndCHD +
     rndRESULTfac, data = .)

df.base <- df.base %>%
  mutate_cond(is.na(EQ5D.score.index.2),
          EQ5D.score.index.2 = predict(score.model.2, newdata = (df.base %>%
filter(
is.na(EQ5D.score.index.2)
                                                                  ))))

score.model.1 <- df.base %>%
  filter(!is.na(EQ5D.score.index.1)) %>%
  lm(EQ5D.score.index.1 ~ EQ5D.score.index.3 + EQ5D.score.index.2 + Sexfac +
      Age + rndCHD + rndRESULTfac, data = .)

df.base <- df.base %>%
  mutate_cond(is.na(EQ5D.score.index.1),
          EQ5D.score.index.1 = predict(score.model.1, newdata = (df.base %>%
filter(is.na(EQ5D.score.index.1)
              ))))
```

```
df.base <- df.base %>%
  mutate(EQ5D.QALY.calc = .25 * (EQ5D.score.index.1 + 2 * EQ5D.score.index.2
+ EQ5D.score.index.3))

EQ5D.QALY.imp.model <-
  lm(EQ5D.QALY.calc ~ rndRESULTfac + rndCHD + rndPATSTATUS, data = df.base)

EQ5D.QALY.model <-
  lm(EQ5D.QALY.calc ~ Sexfac + Age + rndCHD + rndRESULTfac, data = df.base)

directcosts.model.effect <-
  quantreg::rq(
    data = df.base,
    log(directcost_Y_after + 1) ~ rndRESULTfac + rndCHD + Age + Sexfac +
log(directcost_Y_before + 1)
  )
directcosts.model.noeffect <-
  quantreg::rq(data = df.base,
     log(directcost_Y_after   +   1)   ~   rndCHD   +   Age   +   Sexfac   +
log(directcost_Y_before + 1))

directcosts.base.model <-
  df.base %>% filter(!is.na(directcost_Y_after)) %>%
  quantreg::rq(log(directcost_Y_after + 1) ~ rndRESULTfac + rndCHD + Age +
               Sexfac, data = .)
df.base <- df.base %>% mutate_cond(is.na(directcost_Y_after),
           directcost_Y_after = exp(predict(directcosts.base.model, newdata
= (df.base %>% filter(is.na(directcost_Y_after))) )) - 1)

df.base <- df.base %>% mutate(EQ5D.QALY = EQ5D.QALY.calc)

df.simulation <-
  df.base %>% select(Sexfac, Age, rndCHD, rndRESULTfac, rndPATSTATUS,
    EQ5D.QALY, directcost_Y_after, directcost_Y_before)

discount <- 0
n.samples <- 2000
n.repeats <- 1000

results <- list()

directcosts.model <- directcosts.model.effect

num.cores <- detectCores() - 1
cluster <- makeCluster(num.cores)
clusterEvalQ(cluster, {
  library(quantreg)
  library(tidyverse)
})

clusterExport(
  cluster,
  c(
    "df.simulation",
```

```
    "directcosts.model",
    "EQ5D.QALY.model",
    "n.samples",
    "mortality"
  )
)
clusterExport(cluster, c("get_simulation_results", "simulate_data"))

results <-
  parLapply(cluster, 1:n.repeats, function(k)
    get_simulation_results(
      df.simulation,
      directcosts.model,
      EQ5D.QALY.model,
      mortality,
      n.samples
    ))
time_out = Sys.time()

stopCluster(cluster)

survival.median <-
  sapply(1:n.repeats, function(k)
    c(
      which.min(results[[k]]$SoC.survival > 0.5),
      which.min(results[[k]]$NICC.survival > 0.5)
    ))
year.lim <- median(survival.median[2, ])

df.results <- bind_rows(results)
```
