## Supplementary Material 3 for "Cost-effectiveness of a telemonitoring program in patients with cardiovascular diseases compared to standard of care"

Running title: Cost-effectiveness in cardiovascular telemonitoring

Ziegler et al.

### Supplementary Material 3

### 1. EQ5D profile by treatment for all subjects by treatment

Table 1.1: Descriptive statistics for EQ5D profiles. Display is by treatment group.

| | NICC (N=477) | SoC (N=480) | Total (N=957) | p value |
|---|---|---|---|---|
| **EQ5D.QALY** | | | | < 0.001 |
| Mean (SD) | 0.850 (0.150) | 0.799 (0.184) | 0.823 (0.171) | |
| 95% CI | 0.850 (0.834, 0.865) | 0.799 (0.780, 0.817) | 0.823 (0.811, 0.835) | |
| Median (Q1, Q3) | 0.890 (0.805, 0.955) | 0.848 (0.718, 0.914) | 0.869 (0.773, 0.934) | |
| Min - Max | 0.171 - 1.000 | 0.061 - 1.000 | 0.061 - 1.000 | |
| Missing | 111 | 82 | 193 | |
| **EQ5D VAS adjusted life-years** | | | | 0.003 |
| Mean (SD) | 0.719 (0.159) | 0.684 (0.168) | 0.701 (0.164) | |
| 95% CI | 0.719 (0.703, 0.735) | 0.684 (0.667, 0.700) | 0.701 (0.689, 0.713) | |
| Median (Q1, Q3) | 0.750 (0.625, 0.838) | 0.723 (0.550, 0.812) | 0.738 (0.588, 0.825) | |
| Min - Max | 0.152 - 1.000 | 0.188 - 1.000 | 0.152 - 1.000 | |
| Missing | 115 | 97 | 212 | |
| **EQ5D VAS at baseline** | | | | 0.436 |
| Mean (SD) | 68.557 (19.121) | 67.569 (18.577) | 68.058 (18.844) | |
| 95% CI | 68.557 (66.757, 70.357) | 67.569 (65.838, 69.299) | 68.058 (66.812, 69.304) | |
| Median (Q1, Q3) | 75.000 (55.000, 80.000) | 70.000 (50.000, 80.000) | 70.000 (50.000, 80.000) | |
| Min - Max | 10.000 - 100.000 | 10.000 - 100.000 | 10.000 - 100.000 | |
| Missing | 41 | 35 | 76 | |
| **EQ5D VAS at 6 mo FU** | | | | < 0.001 |
| Mean (SD) | 72.664 (17.835) | 67.927 (19.010) | 70.190 (18.598) | |
| 95% CI | 72.664 (70.882, 74.447) | 67.927 (66.110, 69.744) | 70.190 (68.907, 71.473) | |
| Median (Q1, Q3) | 75.000 (60.000, 85.000) | 70.000 (50.000, 80.000) | 75.000 (60.000, 85.000) | |
| Min - Max | 3.000 - 100.000 | 20.000 - 100.000 | 3.000 - 100.000 | |
| Missing | 90 | 57 | 147 | |

Table 1.1: Descriptive statistics for EQ5D profiles. Display is by treatment group.

| | NICC (N=477) | SoC (N=480) | Total (N=957) | p value |
|---|---|---|---|---|
| **EQ5D VAS at 12 mo FU** | | | | 0.001 |
| Mean (SD) | 71.908 (18.425) | 67.371 (20.093) | 69.567 (19.424) | |
| 95% CI | 71.908 (70.047, 73.769) | 67.371 (65.406, 69.336) | 69.567 (68.204, 70.930) | |
| Median (Q1, Q3) | 75.000 (60.000, 85.000) | 70.000 (50.000, 80.000) | 75.000 (55.000, 85.000) | |
| Min - Max | 0.000 - 100.000 | 0.000 - 100.000 | 0.000 - 100.000 | |
| Missing | 98 | 76 | 174 | |
| **EQ index at baseline** | | | | 0.023 |
| Mean (SD) | 0.834 (0.163) | 0.808 (0.173) | 0.821 (0.168) | |
| 95% CI | 0.834 (0.819, 0.849) | 0.808 (0.792, 0.824) | 0.821 (0.810, 0.832) | |
| Median (Q1, Q3) | 0.887 (0.788, 0.918) | 0.828 (0.748, 0.910) | 0.828 (0.788, 0.910) | |
| Min - Max | 0.085 - 1.000 | 0.137 - 1.000 | 0.085 - 1.000 | |
| Missing | 40 | 33 | 73 | |
| **EQ index at 6 mo FU** | | | | < 0.001 |
| Mean (SD) | 0.854 (0.169) | 0.801 (0.206) | 0.826 (0.191) | |
| 95% CI | 0.854 (0.837, 0.871) | 0.801 (0.782, 0.821) | 0.826 (0.813, 0.839) | |
| Median (Q1, Q3) | 0.909 (0.806, 1.000) | 0.828 (0.740, 0.910) | 0.887 (0.788, 0.918) | |
| Min - Max | 0.000 - 1.000 | 0.000 - 1.000 | 0.000 - 1.000 | |
| Missing | 91 | 50 | 141 | |
| **EQ index at 12 mo FU** | | | | < 0.001 |
| Mean (SD) | 0.840 (0.184) | 0.766 (0.253) | 0.801 (0.225) | |
| 95% CI | 0.840 (0.822, 0.859) | 0.766 (0.742, 0.790) | 0.801 (0.786, 0.817) | |
| Median (Q1, Q3) | 0.909 (0.806, 0.999) | 0.828 (0.701, 0.910) | 0.887 (0.748, 0.918) | |
| Min - Max | 0.000 - 1.000 | 0.000 - 1.000 | 0.000 - 1.000 | |
| Missing | 96 | 62 | 158 | |
| **EQ5D problem score at baseline** | | | | 0.015 |
| Mean (SD) | 8.529 (3.371) | 9.105 (3.622) | 8.820 (3.510) | |
| 95% CI | 8.529 (8.212, 8.846) | 9.105 (8.768, 9.442) | 8.820 (8.588, 9.052) | |
| Median (Q1, Q3) | 8.000 (6.000, 10.000) | 8.000 (6.000, 11.000) | 8.000 (6.000, 11.000) | |
| Min - Max | 5.000 - 20.000 | 5.000 - 21.000 | 5.000 - 21.000 | |
| Missing | 40 | 33 | 73 | |

Table 1.1: Descriptive statistics for EQ5D profiles. Display is by treatment group.

| | NICC (N=477) | SoC (N=480) | Total (N=957) | p value |
|---|---|---|---|---|
| **EQ5D problem score at 6 mo FU** | | | | < 0.001 |
| Mean (SD) | 7.995 (3.222) | 8.848 (3.575) | 8.441 (3.436) | |
| 95% CI | 7.995 (7.672, 8.318) | 8.848 (8.505, 9.190) | 8.441 (8.203, 8.679) | |
| Median (Q1, Q3) | 7.000 (5.000, 9.000) | 8.000 (6.000, 11.000) | 7.000 (6.000, 10.000) | |
| Min - Max | 5.000 - 20.000 | 5.000 - 21.000 | 5.000 - 21.000 | |
| Missing | 93 | 59 | 152 | |
| **EQ5D problem score at 12 mo FU** | | | | < 0.001 |
| Mean (SD) | 8.119 (3.313) | 9.010 (3.752) | 8.575 (3.570) | |
| 95% CI | 8.119 (7.784, 8.455) | 9.010 (8.639, 9.381) | 8.575 (8.323, 8.827) | |
| Median (Q1, Q3) | 7.000 (6.000, 10.000) | 8.000 (6.000, 11.000) | 7.000 (6.000, 10.000) | |
| Min - Max | 5.000 - 20.000 | 5.000 - 20.000 | 5.000 - 20.000 | |
| Missing | 100 | 85 | 185 | |

### 2. EQ5D profile by treatment for all subjects by treatment and heart disease

Table 2.1: Descriptive statistics for EQ5D profiles. Display is by treatment group and heart disease.

| | NICC.HF (N=202) | SoC.HF (N=204) | NICC.AF (N=132) | SoC.AF (N=133) | NICC.TRH (N=143) | SoC.TRH (N=143) | Total (N=957) | p value |
|---|---|---|---|---|---|---|---|---|
| **EQ5D.QALY** | | | | | | | | < 0.001 |
| Mean (SD) | 0.832 (0.158) | 0.765 (0.205) | 0.878 (0.130) | 0.822 (0.168) | 0.845 (0.154) | 0.824 (0.162) | 0.823 (0.171) | |
| 95% CI | 0.832 (0.806, 0.859) | 0.765 (0.733, 0.796) | 0.878 (0.853, 0.903) | 0.822 (0.790, 0.853) | 0.845 (0.817, 0.874) | 0.824 (0.795, 0.853) | 0.823 (0.811, 0.835) | |
| Median (Q1, Q3) | 0.874 (0.784, 0.933) | 0.818 (0.669, 0.909) | 0.907 (0.823, 0.978) | 0.861 (0.781, 0.911) | 0.889 (0.812, 0.949) | 0.864 (0.749, 0.944) | 0.869 (0.773, 0.934) | |
| Min - Max | 0.171 - 1.000 | 0.081 - 1.000 | 0.351 - 1.000 | 0.197 - 1.000 | 0.324 - 1.000 | 0.061 - 1.000 | 0.061 - 1.000 | |
| Missing | 60 | 39 | 24 | 23 | 27 | 20 | 193 | |
| **EQ5D VAS adjusted life-years** | | | | | | | | < 0.001 |
| Mean (SD) | 0.693 (0.159) | 0.654 (0.178) | 0.753 (0.149) | 0.710 (0.147) | 0.721 (0.164) | 0.698 (0.165) | 0.701 (0.164) | |
| 95% CI | 0.693 (0.666, 0.719) | 0.654 (0.626, 0.683) | 0.753 (0.724, 0.782) | 0.710 (0.681, 0.739) | 0.721 (0.691, 0.751) | 0.698 (0.669, 0.727) | 0.701 (0.689, 0.713) | |
| Median (Q1, Q3) | 0.725 (0.594, 0.821) | 0.662 (0.525, 0.787) | 0.800 (0.681, 0.850) | 0.738 (0.600, 0.831) | 0.750 (0.634, 0.850) | 0.725 (0.588, 0.812) | 0.738 (0.588, 0.825) | |
| Min - Max | 0.225 - 0.985 | 0.200 - 1.000 | 0.152 - 0.990 | 0.375 - 0.975 | 0.175 - 1.000 | 0.188 - 0.950 | 0.152 - 1.000 | |
| Missing | 59 | 48 | 29 | 31 | 27 | 18 | 212 | |
| **EQ5D VAS at baseline** | | | | | | | | < 0.001 |
| Mean (SD) | 64.564 (19.129) | 63.741 (20.079) | 71.883 (19.260) | 71.621 (15.677) | 70.861 (18.163) | 69.081 (18.004) | 68.058 (18.844) | |
| 95% CI | 64.564 (61.743, 67.386) | 63.741 (60.828, 66.653) | 71.883 (68.402, 75.365) | 71.621 (68.834, 74.408) | 70.861 (67.793, 73.930) | 69.081 (66.028, 72.134) | 68.058 (66.812, 69.304) | |
| Median (Q1, Q3) | 70.000 (50.000, 80.000) | 65.000 (50.000, 80.000) | 80.000 (60.000, 85.000) | 75.000 (60.000, 80.000) | 75.000 (60.000, 85.000) | 70.000 (58.750, 80.000) | 70.000 (50.000, 80.000) | |
| Min - Max | 10.000 - 100.000 | 10.000 - 100.000 | 20.000 - 100.000 | 35.000 - 100.000 | 10.000 - 100.000 | 10.000 - 100.000 | 10.000 - 100.000 | |
| Missing | 23 | 19 | 12 | 9 | 6 | 7 | 76 | |

Table 2.1: Descriptive statistics for EQ5D profiles. Display is by treatment group and heart disease.

| | NICC.HF (N=202) | SoC.HF (N=204) | NICC.AF (N=132) | SoC.AF (N=133) | NICC.TRH (N=143) | SoC.TRH (N=143) | Total (N=957) | p value |
|---|---|---|---|---|---|---|---|---|
| **EQ5D VAS at 6 mo FU** | | | | | | | | < 0.001 |
| Mean (SD) | 70.148 (17.587) | 65.178 (20.218) | 75.821 (17.721) | 70.085 (17.063) | 72.967 (17.912) | 69.636 (18.693) | 70.190 (18.598) | |
| 95% CI | 70.148 (67.358, 72.939) | 65.178 (62.153, 68.203) | 75.821 (72.503, 79.140) | 70.085 (66.961, 73.210) | 72.967 (69.729, 76.204) | 69.636 (66.418, 72.855) | 70.190 (68.907, 71.473) | |
| Median (Q1, Q3) | 75.000 (57.500, 85.000) | 65.000 (50.000, 80.000) | 80.000 (70.000, 90.000) | 75.000 (60.000, 80.000) | 75.000 (60.000, 90.000) | 74.000 (60.000, 80.000) | 75.000 (60.000, 85.000) | |
| Min - Max | 20.000 - 100.000 | 20.000 - 100.000 | 3.000 - 100.000 | 25.000 - 100.000 | 20.000 - 100.000 | 20.000 - 100.000 | 3.000 - 100.000 | |
| Missing | 47 | 30 | 20 | 16 | 23 | 11 | 147 | |
| **EQ5D VAS at 12 mo FU** | | | | | | | | < 0.001 |
| Mean (SD) | 70.855 (19.195) | 63.821 (20.862) | 74.514 (15.498) | 70.367 (18.134) | 70.856 (19.765) | 69.496 (20.086) | 69.567 (19.424) | |
| 95% CI | 70.855 (67.779, 73.931) | 63.821 (60.644, 66.999) | 74.514 (71.571, 77.456) | 70.367 (66.924, 73.810) | 70.856 (67.253, 74.459) | 69.496 (65.969, 73.023) | 69.567 (68.204, 70.930) | |
| Median (Q1, Q3) | 75.000 (60.000, 85.000) | 65.000 (50.000, 80.000) | 80.000 (70.000, 85.000) | 75.000 (60.000, 80.000) | 75.000 (60.000, 85.000) | 75.000 (50.000, 85.000) | 75.000 (55.000, 85.000) | |
| Min - Max | 20.000 - 100.000 | 2.000 - 100.000 | 30.000 - 100.000 | 0.000 - 100.000 | 0.000 - 100.000 | 10.000 - 100.000 | 0.000 - 100.000 | |
| Missing | 50 | 36 | 23 | 24 | 25 | 16 | 174 | |
| **EQ index at baseline** | | | | | | | | 0.010 |
| Mean (SD) | 0.818 (0.160) | 0.784 (0.187) | 0.849 (0.173) | 0.834 (0.152) | 0.841 (0.155) | 0.818 (0.166) | 0.821 (0.168) | |
| 95% CI | 0.818 (0.795, 0.842) | 0.784 (0.758, 0.811) | 0.849 (0.818, 0.880) | 0.834 (0.807, 0.861) | 0.841 (0.814, 0.867) | 0.818 (0.790, 0.846) | 0.821 (0.810, 0.832) | |
| Median (Q1, Q3) | 0.828 (0.788, 0.910) | 0.806 (0.716, 0.910) | 0.909 (0.806, 0.999) | 0.887 (0.788, 0.910) | 0.887 (0.804, 0.918) | 0.828 (0.755, 0.910) | 0.828 (0.788, 0.910) | |
| Min - Max | 0.159 - 1.000 | 0.137 - 1.000 | 0.085 - 1.000 | 0.275 - 1.000 | 0.386 - 1.000 | 0.227 - 1.000 | 0.085 - 1.000 | |
| Missing | 25 | 17 | 8 | 10 | 7 | 6 | 73 | |

Table 2.1: Descriptive statistics for EQ5D profiles. Display is by treatment group and heart disease.

| | NICC.HF (N=202) | SoC.HF (N=204) | NICC.AF (N=132) | SoC.AF (N=133) | NICC.TRH (N=143) | SoC.TRH (N=143) | Total (N=957) | p value |
|---|---|---|---|---|---|---|---|---|
| **EQ index at 6 mo FU** | | | | | | | | < 0.001 |
| Mean (SD) | 0.839 (0.171) | 0.772 (0.225) | 0.878 (0.160) | 0.818 (0.200) | 0.851 (0.172) | 0.825 (0.179) | 0.826 (0.191) | |
| 95% CI | 0.839 (0.812, 0.866) | 0.772 (0.738, 0.805) | 0.878 (0.848, 0.908) | 0.818 (0.783, 0.854) | 0.851 (0.820, 0.882) | 0.825 (0.794, 0.856) | 0.826 (0.813, 0.839) | |
| Median (Q1, Q3) | 0.887 (0.788, 0.918) | 0.828 (0.701, 0.910) | 0.910 (0.828, 1.000) | 0.828 (0.788, 0.910) | 0.909 (0.806, 0.999) | 0.887 (0.788, 0.910) | 0.887 (0.788, 0.918) | |
| Min - Max | 0.000 - 1.000 | 0.000 - 1.000 | 0.194 - 1.000 | 0.000 - 1.000 | 0.159 - 1.000 | 0.000 - 1.000 | 0.000 - 1.000 | |
| Missing | 46 | 27 | 21 | 10 | 24 | 13 | 141 | |
| **EQ index at 12 mo FU** | | | | | | | | < 0.001 |
| Mean (SD) | 0.817 (0.209) | 0.731 (0.271) | 0.881 (0.128) | 0.781 (0.262) | 0.831 (0.190) | 0.801 (0.209) | 0.801 (0.225) | |
| 95% CI | 0.817 (0.784, 0.851) | 0.731 (0.690, 0.771) | 0.881 (0.857, 0.905) | 0.781 (0.733, 0.830) | 0.831 (0.796, 0.865) | 0.801 (0.764, 0.837) | 0.801 (0.786, 0.817) | |
| Median (Q1, Q3) | 0.900 (0.788, 0.918) | 0.810 (0.661, 0.910) | 0.909 (0.810, 0.999) | 0.887 (0.737, 0.910) | 0.909 (0.788, 0.999) | 0.828 (0.712, 0.938) | 0.887 (0.748, 0.918) | |
| Min - Max | 0.000 - 1.000 | 0.000 - 1.000 | 0.351 - 1.000 | 0.000 - 1.000 | 0.129 - 1.000 | 0.000 - 1.000 | 0.000 - 1.000 | |
| Missing | 51 | 30 | 21 | 17 | 24 | 15 | 158 | |
| **EQ5D problem score at baseline** | | | | | | | | < 0.001 |
| Mean (SD) | 8.944 (3.250) | 9.807 (3.927) | 8.202 (3.536) | 8.480 (3.220) | 8.287 (3.340) | 8.708 (3.383) | 8.820 (3.510) | |
| 95% CI | 8.944 (8.461, 9.426) | 9.807 (9.241, 10.374) | 8.202 (7.573, 8.830) | 8.480 (7.905, 9.054) | 8.287 (7.720, 8.853) | 8.708 (8.136, 9.280) | 8.820 (8.588, 9.052) | |
| Median (Q1, Q3) | 8.000 (6.000, 11.000) | 9.000 (6.000, 12.000) | 7.000 (5.750, 10.000) | 7.000 (6.000, 10.000) | 7.000 (6.000, 10.000) | 8.000 (6.000, 11.000) | 8.000 (6.000, 11.000) | |
| Min - Max | 5.000 - 20.000 | 5.000 - 21.000 | 5.000 - 19.000 | 5.000 - 18.000 | 5.000 - 19.000 | 5.000 - 21.000 | 5.000 - 21.000 | |
| Missing | 25 | 17 | 8 | 10 | 7 | 6 | 73 | |

Table 2.1: Descriptive statistics for EQ5D profiles. Display is by treatment group and heart disease.

| | NICC.HF (N=202) | SoC.HF (N=204) | NICC.AF (N=132) | SoC.AF (N=133) | NICC.TRH (N=143) | SoC.TRH (N=143) | Total (N=957) | p value |
|---|---|---|---|---|---|---|---|---|
| **EQ5D problem score at 6 mo FU** | | | | | | | | < 0.001 |
| Mean (SD) | 8.344 (3.103) | 9.442 (3.865) | 7.532 (3.233) | 8.350 (3.064) | 7.975 (3.333) | 8.519 (3.527) | 8.441 (3.436) | |
| 95% CI | 8.344 (7.850, 8.838) | 9.442 (8.860, 10.024) | 7.532 (6.923, 8.140) | 8.350 (7.796, 8.904) | 7.975 (7.370, 8.580) | 8.519 (7.905, 9.134) | 8.441 (8.203, 8.679) | |
| Median (Q1, Q3) | 8.000 (6.000, 10.000) | 8.000 (6.000, 12.000) | 6.000 (5.000, 9.000) | 7.500 (6.000, 10.000) | 7.000 (6.000, 9.000) | 7.000 (6.000, 10.000) | 7.000 (6.000, 10.000) | |
| Min - Max | 5.000 - 20.000 | 5.000 - 21.000 | 5.000 - 19.000 | 5.000 - 20.000 | 5.000 - 20.000 | 5.000 - 19.000 | 5.000 - 21.000 | |
| Missing | 48 | 32 | 21 | 13 | 24 | 14 | 152 | |
| **EQ5D problem score at 12 mo FU** | | | | | | | | < 0.001 |
| Mean (SD) | 8.374 (3.336) | 9.602 (3.971) | 7.721 (2.939) | 8.444 (3.449) | 8.176 (3.595) | 8.738 (3.636) | 8.575 (3.570) | |
| 95% CI | 8.374 (7.830, 8.918) | 9.602 (8.984, 10.220) | 7.721 (7.168, 8.274) | 8.444 (7.787, 9.102) | 8.176 (7.524, 8.829) | 8.738 (8.097, 9.379) | 8.575 (8.323, 8.827) | |
| Median (Q1, Q3) | 7.000 (6.000, 10.000) | 9.000 (6.000, 12.000) | 7.000 (6.000, 9.000) | 7.000 (6.000, 10.000) | 7.000 (6.000, 10.000) | 7.000 (6.000, 11.000) | 7.000 (6.000, 10.000) | |
| Min - Max | 5.000 - 19.000 | 5.000 - 20.000 | 5.000 - 19.000 | 5.000 - 19.000 | 5.000 - 20.000 | 5.000 - 18.000 | 5.000 - 20.000 | |
| Missing | 55 | 43 | 21 | 25 | 24 | 17 | 185 | |

### 3. EQ5D profile by treatment for AF subjects by treatment

Table 3.1: Descriptive statistics for EQ5D profiles in patients with AF. Display is by treatment group.

| | NICC (N=132) | SoC (N=133) | Total (N=265) | p value |
|---|---|---|---|---|
| **EQ5D.QALY** | | | | 0.006 |
| Mean (SD) | 0.878 (0.130) | 0.822 (0.168) | 0.849 (0.153) | |
| 95% CI | 0.878 (0.853, 0.903) | 0.822 (0.790, 0.853) | 0.849 (0.829, 0.870) | |
| Median (Q1, Q3) | 0.907 (0.823, 0.978) | 0.861 (0.781, 0.911) | 0.889 (0.802, 0.955) | |
| Min - Max | 0.351 - 1.000 | 0.197 - 1.000 | 0.197 - 1.000 | |
| Missing | 24 | 23 | 47 | |
| **EQ5D VAS adjusted life-years** | | | | 0.039 |
| Mean (SD) | 0.753 (0.149) | 0.710 (0.147) | 0.732 (0.149) | |
| 95% CI | 0.753 (0.724, 0.782) | 0.710 (0.681, 0.739) | 0.732 (0.711, 0.752) | |
| Median (Q1, Q3) | 0.800 (0.681, 0.850) | 0.738 (0.600, 0.831) | 0.762 (0.650, 0.838) | |
| Min - Max | 0.152 - 0.990 | 0.375 - 0.975 | 0.152 - 0.990 | |
| Missing | 29 | 31 | 60 | |
| **EQ5D VAS at baseline** | | | | 0.907 |
| Mean (SD) | 71.883 (19.260) | 71.621 (15.677) | 71.750 (17.495) | |
| 95% CI | 71.883 (68.402, 75.365) | 71.621 (68.834, 74.408) | 71.750 (69.544, 73.956) | |
| Median (Q1, Q3) | 80.000 (60.000, 85.000) | 75.000 (60.000, 80.000) | 75.000 (60.000, 85.000) | |
| Min - Max | 20.000 - 100.000 | 35.000 - 100.000 | 20.000 - 100.000 | |
| Missing | 12 | 9 | 21 | |
| **EQ5D VAS at 6 mo FU** | | | | 0.013 |
| Mean (SD) | 75.821 (17.721) | 70.085 (17.063) | 72.891 (17.586) | |
| 95% CI | 75.821 (72.503, 79.140) | 70.085 (66.961, 73.210) | 72.891 (70.601, 75.181) | |
| Median (Q1, Q3) | 80.000 (70.000, 90.000) | 75.000 (60.000, 80.000) | 75.000 (60.000, 85.000) | |
| Min - Max | 3.000 - 100.000 | 25.000 - 100.000 | 3.000 - 100.000 | |
| Missing | 20 | 16 | 36 | |
| **EQ5D VAS at 12 mo FU** | | | | 0.071 |
| Mean (SD) | 74.514 (15.498) | 70.367 (18.134) | 72.440 (16.956) | |
| 95% CI | 74.514 (71.571, 77.456) | 70.367 (66.924, 73.810) | 72.440 (70.177, 74.704) | |
| Median (Q1, Q3) | 80.000 (70.000, 85.000) | 75.000 (60.000, 80.000) | 75.000 (65.000, 85.000) | |
| Min - Max | 30.000 - 100.000 | 0.000 - 100.000 | 0.000 - 100.000 | |
| Missing | 23 | 24 | 47 | |

Table 3.1: Descriptive statistics for EQ5D profiles in patients with AF. Display is by treatment group.

| | NICC (N=132) | SoC (N=133) | Total (N=265) | p value |
|---|---|---|---|---|
| **EQ index at baseline** | | | | 0.466 |
| Mean (SD) | 0.849 (0.173) | 0.834 (0.152) | 0.842 (0.163) | |
| 95% CI | 0.849 (0.818, 0.880) | 0.834 (0.807, 0.861) | 0.842 (0.821, 0.862) | |
| Median (Q1, Q3) | 0.909 (0.806, 0.999) | 0.887 (0.788, 0.910) | 0.887 (0.788, 0.999) | |
| Min - Max | 0.085 - 1.000 | 0.275 - 1.000 | 0.085 - 1.000 | |
| Missing | 8 | 10 | 18 | |
| **EQ index at 6 mo FU** | | | | 0.013 |
| Mean (SD) | 0.878 (0.160) | 0.818 (0.200) | 0.847 (0.184) | |
| 95% CI | 0.878 (0.848, 0.908) | 0.818 (0.783, 0.854) | 0.847 (0.823, 0.871) | |
| Median (Q1, Q3) | 0.910 (0.828, 1.000) | 0.828 (0.788, 0.910) | 0.909 (0.806, 0.999) | |
| Min - Max | 0.194 - 1.000 | 0.000 - 1.000 | 0.000 - 1.000 | |
| Missing | 21 | 10 | 31 | |
| **EQ index at 12 mo FU** | | | | < 0.001 |
| Mean (SD) | 0.881 (0.128) | 0.781 (0.262) | 0.830 (0.213) | |
| 95% CI | 0.881 (0.857, 0.905) | 0.781 (0.733, 0.830) | 0.830 (0.802, 0.858) | |
| Median (Q1, Q3) | 0.909 (0.810, 0.999) | 0.887 (0.737, 0.910) | 0.909 (0.806, 0.999) | |
| Min - Max | 0.351 - 1.000 | 0.000 - 1.000 | 0.000 - 1.000 | |
| Missing | 21 | 17 | 38 | |
| **EQ5D problem score at baseline** | | | | 0.519 |
| Mean (SD) | 8.202 (3.536) | 8.480 (3.220) | 8.340 (3.378) | |
| 95% CI | 8.202 (7.573, 8.830) | 8.480 (7.905, 9.054) | 8.340 (7.917, 8.763) | |
| Median (Q1, Q3) | 7.000 (5.750, 10.000) | 7.000 (6.000, 10.000) | 7.000 (6.000, 10.000) | |
| Min - Max | 5.000 - 19.000 | 5.000 - 18.000 | 5.000 - 19.000 | |
| Missing | 8 | 10 | 18 | |
| **EQ5D problem score at 6 mo FU** | | | | 0.049 |
| Mean (SD) | 7.532 (3.233) | 8.350 (3.064) | 7.957 (3.166) | |
| 95% CI | 7.532 (6.923, 8.140) | 8.350 (7.796, 8.904) | 7.957 (7.546, 8.367) | |
| Median (Q1, Q3) | 6.000 (5.000, 9.000) | 7.500 (6.000, 10.000) | 7.000 (5.500, 10.000) | |
| Min - Max | 5.000 - 19.000 | 5.000 - 20.000 | 5.000 - 20.000 | |
| Missing | 21 | 13 | 34 | |

Table 3.1: Descriptive statistics for EQ5D profiles in patients with AF. Display is by treatment group.

| | NICC (N=132) | SoC (N=133) | Total (N=265) | p value |
|---|---|---|---|---|
| **EQ5D problem score at 12 mo FU** | | | | 0.096 |
| Mean (SD) | 7.721 (2.939) | 8.444 (3.449) | 8.078 (3.214) | |
| 95% CI | 7.721 (7.168, 8.274) | 8.444 (7.787, 9.102) | 8.078 (7.650, 8.506) | |
| Median (Q1, Q3) | 7.000 (6.000, 9.000) | 7.000 (6.000, 10.000) | 7.000 (6.000, 10.000) | |
| Min - Max | 5.000 - 19.000 | 5.000 - 19.000 | 5.000 - 19.000 | |
| Missing | 21 | 25 | 46 | |

### 4. EQ5D profile by treatment for HF subjects by treatment

Table 4.1: Descriptive statistics for EQ5D profiles in patients with HF Display is by treatment group.

| | NICC (N=202) | SoC (N=204) | Total (N=406) | p value |
|---|---|---|---|---|
| **EQ5D.QALY** | | | | 0.001 |
| Mean (SD) | 0.832 (0.158) | 0.765 (0.205) | 0.796 (0.188) | |
| 95% CI | 0.832 (0.806, 0.859) | 0.765 (0.733, 0.796) | 0.796 (0.775, 0.817) | |
| Median (Q1, Q3) | 0.874 (0.784, 0.933) | 0.818 (0.669, 0.909) | 0.848 (0.724, 0.919) | |
| Min - Max | 0.171 - 1.000 | 0.081 - 1.000 | 0.081 - 1.000 | |
| Missing | 60 | 39 | 99 | |
| **EQ5D VAS adjusted life-years** | | | | 0.052 |
| Mean (SD) | 0.693 (0.159) | 0.654 (0.178) | 0.673 (0.170) | |
| 95% CI | 0.693 (0.666, 0.719) | 0.654 (0.626, 0.683) | 0.673 (0.653, 0.692) | |
| Median (Q1, Q3) | 0.725 (0.594, 0.821) | 0.662 (0.525, 0.787) | 0.688 (0.550, 0.810) | |
| Min - Max | 0.225 - 0.985 | 0.200 - 1.000 | 0.200 - 1.000 | |
| Missing | 59 | 48 | 107 | |
| **EQ5D VAS at baseline** | | | | 0.689 |
| Mean (SD) | 64.564 (19.129) | 63.741 (20.079) | 64.146 (19.595) | |
| 95% CI | 64.564 (61.743, 67.386) | 63.741 (60.828, 66.653) | 64.146 (62.126, 66.165) | |
| Median (Q1, Q3) | 70.000 (50.000, 80.000) | 65.000 (50.000, 80.000) | 65.000 (50.000, 80.000) | |
| Min - Max | 10.000 - 100.000 | 10.000 - 100.000 | 10.000 - 100.000 | |
| Missing | 23 | 19 | 42 | |
| **EQ5D VAS at 6 mo FU** | | | | 0.019 |
| Mean (SD) | 70.148 (17.587) | 65.178 (20.218) | 67.520 (19.157) | |
| 95% CI | 70.148 (67.358, 72.939) | 65.178 (62.153, 68.203) | 67.520 (65.442, 69.597) | |
| Median (Q1, Q3) | 75.000 (57.500, 85.000) | 65.000 (50.000, 80.000) | 70.000 (50.000, 80.000) | |
| Min - Max | 20.000 - 100.000 | 20.000 - 100.000 | 20.000 - 100.000 | |
| Missing | 47 | 30 | 77 | |
| **EQ5D VAS at 12 mo FU** | | | | 0.002 |
| Mean (SD) | 70.855 (19.195) | 63.821 (20.862) | 67.162 (20.363) | |
| 95% CI | 70.855 (67.779, 73.931) | 63.821 (60.644, 66.999) | 67.162 (64.923, 69.402) | |
| Median (Q1, Q3) | 75.000 (60.000, 85.000) | 65.000 (50.000, 80.000) | 70.000 (50.000, 82.000) | |
| Min - Max | 20.000 - 100.000 | 2.000 - 100.000 | 2.000 - 100.000 | |
| Missing | 50 | 36 | 86 | |

Table 4.1: Descriptive statistics for EQ5D profiles in patients with HF Display is by treatment group.

| | NICC (N=202) | SoC (N=204) | Total (N=406) | p value |
|---|---|---|---|---|
| **EQ index at baseline** | | | | 0.065 |
| Mean (SD) | 0.818 (0.160) | 0.784 (0.187) | 0.801 (0.175) | |
| 95% CI | 0.818 (0.795, 0.842) | 0.784 (0.758, 0.811) | 0.801 (0.783, 0.819) | |
| Median (Q1, Q3) | 0.828 (0.788, 0.910) | 0.806 (0.716, 0.910) | 0.828 (0.738, 0.910) | |
| Min - Max | 0.159 - 1.000 | 0.137 - 1.000 | 0.137 - 1.000 | |
| Missing | 25 | 17 | 42 | |
| **EQ index at 6 mo FU** | | | | 0.003 |
| Mean (SD) | 0.839 (0.171) | 0.772 (0.225) | 0.803 (0.204) | |
| 95% CI | 0.839 (0.812, 0.866) | 0.772 (0.738, 0.805) | 0.803 (0.781, 0.825) | |
| Median (Q1, Q3) | 0.887 (0.788, 0.918) | 0.828 (0.701, 0.910) | 0.828 (0.733, 0.918) | |
| Min - Max | 0.000 - 1.000 | 0.000 - 1.000 | 0.000 - 1.000 | |
| Missing | 46 | 27 | 73 | |
| **EQ index at 12 mo FU** | | | | 0.002 |
| Mean (SD) | 0.817 (0.209) | 0.731 (0.271) | 0.771 (0.248) | |
| 95% CI | 0.817 (0.784, 0.851) | 0.731 (0.690, 0.771) | 0.771 (0.744, 0.798) | |
| Median (Q1, Q3) | 0.900 (0.788, 0.918) | 0.810 (0.661, 0.910) | 0.828 (0.716, 0.910) | |
| Min - Max | 0.000 - 1.000 | 0.000 - 1.000 | 0.000 - 1.000 | |
| Missing | 51 | 30 | 81 | |
| **EQ5D problem score at baseline** | | | | 0.023 |
| Mean (SD) | 8.944 (3.250) | 9.807 (3.927) | 9.387 (3.635) | |
| 95% CI | 8.944 (8.461, 9.426) | 9.807 (9.241, 10.374) | 9.387 (9.013, 9.762) | |
| Median (Q1, Q3) | 8.000 (6.000, 11.000) | 9.000 (6.000, 12.000) | 9.000 (6.000, 12.000) | |
| Min - Max | 5.000 - 20.000 | 5.000 - 21.000 | 5.000 - 21.000 | |
| Missing | 25 | 17 | 42 | |
| **EQ5D problem score at 6 mo FU** | | | | 0.005 |
| Mean (SD) | 8.344 (3.103) | 9.442 (3.865) | 8.923 (3.563) | |
| 95% CI | 8.344 (7.850, 8.838) | 9.442 (8.860, 10.024) | 8.923 (8.535, 9.311) | |
| Median (Q1, Q3) | 8.000 (6.000, 10.000) | 8.000 (6.000, 12.000) | 8.000 (6.000, 11.000) | |
| Min - Max | 5.000 - 20.000 | 5.000 - 21.000 | 5.000 - 21.000 | |
| Missing | 48 | 32 | 80 | |

Table 4.1: Descriptive statistics for EQ5D profiles in patients with HF Display is by treatment group.

| | NICC (N=202) | SoC (N=204) | Total (N=406) | p value |
|---|---|---|---|---|
| **EQ5D problem score at 12 mo FU** | | | | 0.004 |
| Mean (SD) | 8.374 (3.336) | 9.602 (3.971) | 9.016 (3.726) | |
| 95% CI | 8.374 (7.830, 8.918) | 9.602 (8.984, 10.220) | 9.016 (8.598, 9.434) | |
| Median (Q1, Q3) | 7.000 (6.000, 10.000) | 9.000 (6.000, 12.000) | 8.000 (6.000, 11.000) | |
| Min - Max | 5.000 - 19.000 | 5.000 - 20.000 | 5.000 - 20.000 | |
| Missing | 55 | 43 | 98 | |

### 5. EQ5D profile by treatment for TRH subjects by treatment

Table 5.1: Descriptive statistics for EQ5D profiles in patients with TRH. Display is by treatment group.

| | NICC (N=143) | SoC (N=143) | Total (N=286) | p value |
|---|---|---|---|---|
| **EQ5D.QALY** | | | | 0.297 |
| Mean (SD) | 0.845 (0.154) | 0.824 (0.162) | 0.834 (0.158) | |
| 95% CI | 0.845 (0.817, 0.874) | 0.824 (0.795, 0.853) | 0.834 (0.814, 0.854) | |
| Median (Q1, Q3) | 0.889 (0.812, 0.949) | 0.864 (0.749, 0.944) | 0.879 (0.782, 0.949) | |
| Min - Max | 0.324 - 1.000 | 0.061 - 1.000 | 0.061 - 1.000 | |
| Missing | 27 | 20 | 47 | |
| **EQ5D VAS adjusted life-years** | | | | 0.278 |
| Mean (SD) | 0.721 (0.164) | 0.698 (0.165) | 0.709 (0.164) | |
| 95% CI | 0.721 (0.691, 0.751) | 0.698 (0.669, 0.727) | 0.709 (0.688, 0.730) | |
| Median (Q1, Q3) | 0.750 (0.634, 0.850) | 0.725 (0.588, 0.812) | 0.738 (0.613, 0.838) | |
| Min - Max | 0.175 - 1.000 | 0.188 - 0.950 | 0.175 - 1.000 | |
| Missing | 27 | 18 | 45 | |
| **EQ5D VAS at baseline** | | | | 0.417 |
| Mean (SD) | 70.861 (18.163) | 69.081 (18.004) | 69.974 (18.073) | |
| 95% CI | 70.861 (67.793, 73.930) | 69.081 (66.028, 72.134) | 69.974 (67.821, 72.128) | |
| Median (Q1, Q3) | 75.000 (60.000, 85.000) | 70.000 (58.750, 80.000) | 75.000 (60.000, 85.000) | |
| Min - Max | 10.000 - 100.000 | 10.000 - 100.000 | 10.000 - 100.000 | |
| Missing | 6 | 7 | 13 | |
| **EQ5D VAS at 6 mo FU** | | | | 0.151 |
| Mean (SD) | 72.967 (17.912) | 69.636 (18.693) | 71.222 (18.364) | |
| 95% CI | 72.967 (69.729, 76.204) | 69.636 (66.418, 72.855) | 71.222 (68.944, 73.501) | |
| Median (Q1, Q3) | 75.000 (60.000, 90.000) | 74.000 (60.000, 80.000) | 75.000 (60.000, 85.000) | |
| Min - Max | 20.000 - 100.000 | 20.000 - 100.000 | 20.000 - 100.000 | |
| Missing | 23 | 11 | 34 | |
| **EQ5D VAS at 12 mo FU** | | | | 0.594 |
| Mean (SD) | 70.856 (19.765) | 69.496 (20.086) | 70.151 (19.903) | |
| 95% CI | 70.856 (67.253, 74.459) | 69.496 (65.969, 73.023) | 70.151 (67.646, 72.656) | |
| Median (Q1, Q3) | 75.000 (60.000, 85.000) | 75.000 (50.000, 85.000) | 75.000 (60.000, 85.000) | |
| Min - Max | 0.000 - 100.000 | 10.000 - 100.000 | 0.000 - 100.000 | |
| Missing | 25 | 16 | 41 | |

Table 5.1: Descriptive statistics for EQ5D profiles in patients with TRH. Display is by treatment group.

| | NICC (N=143) | SoC (N=143) | Total (N=286) | p value |
|---|---|---|---|---|
| **EQ index at baseline** | | | | 0.246 |
| Mean (SD) | 0.841 (0.155) | 0.818 (0.166) | 0.829 (0.161) | |
| 95% CI | 0.841 (0.814, 0.867) | 0.818 (0.790, 0.846) | 0.829 (0.810, 0.848) | |
| Median (Q1, Q3) | 0.887 (0.804, 0.918) | 0.828 (0.755, 0.910) | 0.887 (0.788, 0.918) | |
| Min - Max | 0.386 - 1.000 | 0.227 - 1.000 | 0.227 - 1.000 | |
| Missing | 7 | 6 | 13 | |
| **EQ index at 6 mo FU** | | | | 0.253 |
| Mean (SD) | 0.851 (0.172) | 0.825 (0.179) | 0.837 (0.176) | |
| 95% CI | 0.851 (0.820, 0.882) | 0.825 (0.794, 0.856) | 0.837 (0.815, 0.859) | |
| Median (Q1, Q3) | 0.909 (0.806, 0.999) | 0.887 (0.788, 0.910) | 0.909 (0.788, 0.999) | |
| Min - Max | 0.159 - 1.000 | 0.000 - 1.000 | 0.000 - 1.000 | |
| Missing | 24 | 13 | 37 | |
| **EQ index at 12 mo FU** | | | | 0.242 |
| Mean (SD) | 0.831 (0.190) | 0.801 (0.209) | 0.815 (0.200) | |
| 95% CI | 0.831 (0.796, 0.865) | 0.801 (0.764, 0.837) | 0.815 (0.790, 0.840) | |
| Median (Q1, Q3) | 0.909 (0.788, 0.999) | 0.828 (0.712, 0.938) | 0.887 (0.736, 0.999) | |
| Min - Max | 0.129 - 1.000 | 0.000 - 1.000 | 0.000 - 1.000 | |
| Missing | 24 | 15 | 39 | |
| **EQ5D problem score at baseline** | | | | 0.301 |
| Mean (SD) | 8.287 (3.340) | 8.708 (3.383) | 8.498 (3.362) | |
| 95% CI | 8.287 (7.720, 8.853) | 8.708 (8.136, 9.280) | 8.498 (8.098, 8.899) | |
| Median (Q1, Q3) | 7.000 (6.000, 10.000) | 8.000 (6.000, 11.000) | 8.000 (6.000, 10.000) | |
| Min - Max | 5.000 - 19.000 | 5.000 - 21.000 | 5.000 - 21.000 | |
| Missing | 7 | 6 | 13 | |
| **EQ5D problem score at 6 mo FU** | | | | 0.213 |
| Mean (SD) | 7.975 (3.333) | 8.519 (3.527) | 8.258 (3.439) | |
| 95% CI | 7.975 (7.370, 8.580) | 8.519 (7.905, 9.134) | 8.258 (7.828, 8.688) | |
| Median (Q1, Q3) | 7.000 (6.000, 9.000) | 7.000 (6.000, 10.000) | 7.000 (6.000, 10.000) | |
| Min - Max | 5.000 - 20.000 | 5.000 - 19.000 | 5.000 - 20.000 | |
| Missing | 24 | 14 | 38 | |

Table 5.1: Descriptive statistics for EQ5D profiles in patients with TRH. Display is by treatment group.

| | NICC (N=143) | SoC (N=143) | Total (N=286) | p value |
|---|---|---|---|---|
| **EQ5D problem score at 12 mo FU** | | | | 0.226 |
| Mean (SD) | 8.176 (3.595) | 8.738 (3.636) | 8.465 (3.620) | |
| 95% CI | 8.176 (7.524, 8.829) | 8.738 (8.097, 9.379) | 8.465 (8.010, 8.921) | |
| Median (Q1, Q3) | 7.000 (6.000, 10.000) | 7.000 (6.000, 11.000) | 7.000 (6.000, 11.000) | |
| Min - Max | 5.000 - 20.000 | 5.000 - 18.000 | 5.000 - 20.000 | |
| Missing | 24 | 17 | 41 | |

### 6. Regression models for important EQ5D variables

#### *6.1 Models with adjustment for stratification variables. Outcome variable: QALY*

Table 6.1: Linear model for EQ5D QALY at 1 y FU with adjustments for stratification variables. Complete case data analysis. Deceased patients scored as 0.

| Variable | Estimate | CI low | CI up | p.value |
|---|---|---|---|---|
| Intercept | 0.8081 | 0.7308 | 0.8854 | 0.0000 |
| SoC | -0.0496 | -0.0733 | -0.0258 | 0.0000 |
| AF | 0.0536 | 0.0245 | 0.0827 | 0.0003 |
| TRH | 0.0362 | 0.0079 | 0.0645 | 0.0123 |
| Outpatient | 0.0660 | -0.0010 | 0.1329 | 0.0537 |

Table 6.2: Linear model for EQ5D QALY at 1 y FU with adjustments for stratification variables. Complete case data analysis. Deceased patients excluded.

| Variable | Estimate | CI low | CI up | p.value |
|---|---|---|---|---|
| Intercept | 0.8470 | 0.7767 | 0.9174 | 0.0000 |
| SoC | -0.0330 | -0.0541 | -0.0118 | 0.0023 |
| AF | 0.0398 | 0.0138 | 0.0659 | 0.0028 |
| TRH | 0.0168 | -0.0083 | 0.0420 | 0.1902 |
| Outpatient | 0.0245 | -0.0367 | 0.0857 | 0.4335 |

Table 6.3: Linear model for EQ5D QALY at 1 y FU with adjustments for stratification variables. Multiple imputation data analysis. Deceased patients scored as 0.

| Variable | Estimate | CI low | CI up | p.value |
|---|---|---|---|---|
| Intercept | 0.7943 | 0.7378 | 0.8508 | 0.0000 |
| SoC | -0.0309 | -0.0499 | -0.0119 | 0.0015 |
| AF | 0.0319 | 0.0086 | 0.0552 | 0.0075 |
| TRH | 0.0118 | -0.0109 | 0.0345 | 0.3084 |
| Outpatient | 0.0413 | -0.0140 | 0.0966 | 0.1432 |

Table 6.4: Linear model for EQ5D QALY at 1 y FU with adjustments for stratification variables. Multiple imputation data analysis. Deceased patients excluded.

| Variable | Estimate | CI low | CI up | p.value |
|---|---|---|---|---|
| Intercept | 0.7951 | 0.7387 | 0.8514 | 0.0000 |
| SoC | -0.0314 | -0.0505 | -0.0122 | 0.0014 |
| AF | 0.0312 | 0.0079 | 0.0545 | 0.0087 |
| TRH | 0.0116 | -0.0111 | 0.0343 | 0.3171 |
| Outpatient | 0.0409 | -0.0140 | 0.0959 | 0.1449 |

*6.2 Models with adjustment for all variables. Outcome variable: QALY*

Table 6.5: Linear model for EQ5D QALY at 1 y FU with adjustments for stratification and baseline variables. Complete case data analysis. Deceased patients scored as 0.

| **Variable** | **Estimate** | **CI low** | **CI up** | **p.value** |
|---|---|---|---|---|
| Intercept | 0.2315 | 0.0580 | 0.4050 | 0.0091 |
| SoC | -0.0262 | -0.0415 | -0.0109 | 0.0008 |
| AF | 0.0092 | -0.0101 | 0.0286 | 0.3504 |
| TRH | 0.0062 | -0.0125 | 0.0248 | 0.5159 |
| Outpatient | 0.0343 | -0.0105 | 0.0790 | 0.1337 |
| Female sex | 0.0007 | -0.0158 | 0.0173 | 0.9305 |
| Age | -0.0015 | -0.0023 | -0.0008 | 0.0001 |
| HeartQoL | 0.0642 | 0.0438 | 0.0846 | 0.0000 |
| EQ5D VAS | 0.0002 | -0.0004 | 0.0008 | 0.4748 |
| EQ5D index | 0.5781 | 0.5102 | 0.6460 | 0.0000 |
| GAD-7 | 0.0022 | -0.0012 | 0.0056 | 0.2011 |
| PHQ-9 | -0.0002 | -0.0036 | 0.0032 | 0.9119 |
| WHO-5 | 0.0000 | -0.0005 | 0.0005 | 0.9918 |
| BMQ general harm | 0.0008 | -0.0021 | 0.0038 | 0.5728 |
| BMQ general overuse | -0.0011 | -0.0035 | 0.0013 | 0.3528 |
| BMQ specific concerns | 0.0002 | -0.0016 | 0.0020 | 0.7924 |
| BMQ specific necessity | 0.0009 | -0.0015 | 0.0033 | 0.4564 |
| MARS-5 | 0.0006 | -0.0045 | 0.0056 | 0.8263 |
| PAM13-D score | 0.0004 | -0.0001 | 0.0010 | 0.1142 |
| ISSS-8 detrimental interaction | 0.0003 | -0.0020 | 0.0027 | 0.7812 |
| ISSS-8 positive support | 0.0011 | -0.0012 | 0.0033 | 0.3626 |

Table 6.6: Linear model for EQ5D QALY at 1 y FU with adjustments for stratification and baseline variables. Complete case data analysis. Deceased patients excluded.

| Variable | Estimate | CI low | CI up | p.value |
|---|---|---|---|---|
| Intercept | 0.2578 | 0.1312 | 0.3844 | 0.0001 |
| SoC | -0.0138 | -0.0249 | -0.0026 | 0.0156 |
| AF | 0.0031 | -0.0110 | 0.0173 | 0.6627 |
| TRH | -0.0044 | -0.0180 | 0.0091 | 0.5213 |
| Outpatient | 0.0100 | -0.0236 | 0.0436 | 0.5606 |
| Female sex | -0.0043 | -0.0164 | 0.0078 | 0.4847 |
| Age | -0.0010 | -0.0015 | -0.0004 | 0.0006 |
| HeartQoL | 0.0470 | 0.0320 | 0.0619 | 0.0000 |
| EQ5D VAS | 0.0003 | -0.0001 | 0.0007 | 0.1381 |
| EQ5D index | 0.5773 | 0.5271 | 0.6275 | 0.0000 |
| GAD-7 | 0.0031 | 0.0007 | 0.0055 | 0.0129 |
| PHQ-9 | -0.0018 | -0.0043 | 0.0007 | 0.1590 |
| WHO-5 | 0.0003 | -0.0001 | 0.0007 | 0.1025 |
| BMQ general harm | 0.0020 | -0.0001 | 0.0042 | 0.0656 |
| BMQ general overuse | -0.0008 | -0.0025 | 0.0010 | 0.3885 |
| BMQ specific concerns | -0.0013 | -0.0027 | 0.0000 | 0.0441 |
| BMQ specific necessity | 0.0004 | -0.0014 | 0.0021 | 0.6947 |
| MARS-5 | 0.0008 | -0.0029 | 0.0044 | 0.6789 |
| PAM13-D score | 0.0002 | -0.0002 | 0.0006 | 0.3469 |
| ISSS-8 detrimental interaction | -0.0001 | -0.0018 | 0.0015 | 0.8649 |
| ISSS-8 positive support | 0.0012 | -0.0005 | 0.0028 | 0.1598 |

Table 6.7: Linear model for EQ5D QALY at 1 y FU with adjustments for stratification and baseline variables. Multiple imputation data analysis. Deceased patients scored as 0.

| Variable | Estimate | CI low | CI up | p.value |
|---|---|---|---|---|
| Intercept | 0.2533 | 0.1288 | 0.3779 | 0.0001 |
| SoC | -0.0154 | -0.0263 | -0.0045 | 0.0059 |
| AF | 0.0008 | -0.0128 | 0.0144 | 0.9069 |
| TRH | -0.0133 | -0.0265 | 0.0000 | 0.0502 |
| Outpatient | 0.0136 | -0.0184 | 0.0456 | 0.4066 |
| Female sex | -0.0012 | -0.0130 | 0.0107 | 0.8473 |
| Age | -0.0008 | -0.0014 | -0.0003 | 0.0041 |
| HeartQoL | 0.0356 | 0.0212 | 0.0500 | 0.0000 |
| EQ5D VAS | 0.0003 | -0.0001 | 0.0007 | 0.1735 |
| EQ5D index | 0.5881 | 0.5395 | 0.6367 | 0.0000 |
| GAD-7 | 0.0027 | 0.0003 | 0.0051 | 0.0300 |
| PHQ-9 | -0.0013 | -0.0038 | 0.0012 | 0.3138 |
| WHO-5 | 0.0003 | 0.0000 | 0.0007 | 0.0862 |
| BMQ general harm | 0.0015 | -0.0006 | 0.0035 | 0.1757 |
| BMQ general overuse | -0.0010 | -0.0028 | 0.0007 | 0.2347 |
| BMQ specific concerns | -0.0009 | -0.0022 | 0.0004 | 0.1884 |
| BMQ specific necessity | 0.0002 | -0.0015 | 0.0019 | 0.8193 |
| MARS-5 | 0.0007 | -0.0030 | 0.0044 | 0.6992 |
| PAM13-D score | 0.0002 | -0.0002 | 0.0006 | 0.2807 |
| ISSS-8 detrimental interaction | 0.0001 | -0.0016 | 0.0018 | 0.9425 |
| ISSS-8 positive support | 0.0009 | -0.0007 | 0.0026 | 0.2712 |

Table 6.8: Linear model for EQ5D QALY at 1 y FU with adjustments for stratification and baseline variables. Multiple imputation data analysis. Deceased patients excluded.

| Variable | Estimate | CI low | CI up | p.value |
|---|---|---|---|---|
| Intercept | 0.2571 | 0.1317 | 0.3826 | 0.0001 |
| SoC | -0.0157 | -0.0268 | -0.0046 | 0.0058 |
| AF | 0.0007 | -0.0130 | 0.0145 | 0.9180 |
| TRH | -0.0132 | -0.0266 | 0.0002 | 0.0544 |
| Outpatient | 0.0129 | -0.0191 | 0.0450 | 0.4285 |
| Female sex | -0.0009 | -0.0129 | 0.0112 | 0.8885 |
| Age | -0.0008 | -0.0014 | -0.0003 | 0.0032 |
| HeartQoL | 0.0359 | 0.0210 | 0.0508 | 0.0000 |
| EQ5D VAS | 0.0003 | -0.0001 | 0.0007 | 0.1762 |
| EQ5D index | 0.5871 | 0.5349 | 0.6393 | 0.0000 |
| GAD-7 | 0.0027 | 0.0002 | 0.0052 | 0.0363 |
| PHQ-9 | -0.0014 | -0.0038 | 0.0011 | 0.2655 |
| WHO-5 | 0.0003 | -0.0001 | 0.0007 | 0.1018 |
| BMQ general harm | 0.0013 | -0.0009 | 0.0035 | 0.2363 |
| BMQ general overuse | -0.0011 | -0.0028 | 0.0007 | 0.2324 |
| BMQ specific concerns | -0.0008 | -0.0022 | 0.0005 | 0.2105 |
| BMQ specific necessity | 0.0002 | -0.0016 | 0.0019 | 0.8517 |
| MARS-5 | 0.0007 | -0.0031 | 0.0044 | 0.7224 |
| PAM13-D score | 0.0002 | -0.0002 | 0.0006 | 0.2425 |
| ISSS-8 detrimental interaction | 0.0001 | -0.0016 | 0.0018 | 0.8894 |
| ISSS-8 positive support | 0.0009 | -0.0007 | 0.0025 | 0.2491 |

#### *6.3 Models with adjustment for stratification variables. Outcome variable: VAS-AL*

Table 6.9: Linear model for EQ5D VAS-AL at 1 y FU with adjustments for stratification variables. Complete case data analysis. Deceased patients scored as 0.

| Variable | Estimate | CI low | CI up | p.value |
|---|---|---|---|---|
| Intercept | 0.6635 | 0.5860 | 0.7410 | 0.0000 |
| SoC | -0.0342 | -0.0574 | -0.0109 | 0.0041 |
| AF | 0.0600 | 0.0312 | 0.0889 | 0.0000 |
| TRH | 0.0356 | 0.0081 | 0.0630 | 0.0113 |
| Outpatient | 0.0629 | -0.0047 | 0.1304 | 0.0684 |

Table 6.10: Linear model for EQ5D VAS-AL at 1 y FU with adjustments for stratification variables. Complete case data analysis. Deceased patients excluded.

| Variable | Estimate | CI low | CI up | p.value |
|---|---|---|---|---|
| Intercept | 0.6635 | 0.5860 | 0.7410 | 0.0000 |
| SoC | -0.0342 | -0.0574 | -0.0109 | 0.0041 |
| AF | 0.0600 | 0.0312 | 0.0889 | 0.0000 |
| TRH | 0.0356 | 0.0081 | 0.0630 | 0.0113 |
| Outpatient | 0.0629 | -0.0047 | 0.1304 | 0.0684 |

Table 6.11: Linear model for EQ5D VAS-AL at 1 y FU with adjustments for stratification variables. Multiple imputation data analysis. Deceased patients scored as 0.

| Variable | Estimate | CI low | CI up | p.value |
|---|---|---|---|---|
| Intercept | 0.6255 | 0.5631 | 0.6880 | 0.0000 |
| SoC | -0.0313 | -0.0526 | -0.0100 | 0.0040 |
| AF | 0.0499 | 0.0239 | 0.0760 | 0.0002 |
| TRH | 0.0364 | 0.0113 | 0.0616 | 0.0047 |
| Outpatient | 0.0605 | -0.0007 | 0.1217 | 0.0530 |

Table 6.12: Linear model for EQ5D VAS-AL at 1 y FU with adjustments for stratification variables. Multiple imputation data analysis. Deceased patients excluded.

| Variable | Estimate | CI low | CI up | p.value |
|---|---|---|---|---|
| Intercept | 0.6242 | 0.5614 | 0.6869 | 0.0000 |
| SoC | -0.0310 | -0.0522 | -0.0098 | 0.0042 |
| AF | 0.0497 | 0.0235 | 0.0759 | 0.0002 |
| TRH | 0.0363 | 0.0108 | 0.0619 | 0.0054 |
| Outpatient | 0.0620 | 0.0008 | 0.1232 | 0.0474 |

#### *6.4 Models with adjustment for all variables. Outcome variable: VAS-AL*

Table 6.13: Linear model for EQ5D VAS-AL at 1 y FU with adjustments for stratification and baseline variables. Complete case data analysis. Deceased patients scored as 0.

| Variable | Estimate | CI low | CI up | p.value |
|---|---|---|---|---|
| Intercept | 0.0998 | -0.0546 | 0.2541 | 0.2056 |
| SoC | -0.0203 | -0.0338 | -0.0067 | 0.0035 |
| AF | 0.0000 | -0.0172 | 0.0172 | 0.9974 |
| TRH | 0.0020 | -0.0143 | 0.0184 | 0.8061 |
| Outpatient | 0.0133 | -0.0278 | 0.0545 | 0.5263 |
| Female sex | 0.0009 | -0.0138 | 0.0156 | 0.9060 |
| Age | -0.0010 | -0.0016 | -0.0003 | 0.0061 |
| HeartQoL | 0.0513 | 0.0332 | 0.0694 | 0.0000 |
| EQ5D VAS | 0.0051 | 0.0046 | 0.0056 | 0.0000 |
| EQ5D index | 0.1162 | 0.0551 | 0.1773 | 0.0002 |
| GAD-7 | 0.0026 | -0.0003 | 0.0056 | 0.0800 |
| PHQ-9 | -0.0004 | -0.0034 | 0.0026 | 0.7888 |
| WHO-5 | 0.0004 | -0.0001 | 0.0008 | 0.1383 |
| BMQ general harm | 0.0040 | 0.0014 | 0.0066 | 0.0027 |
| BMQ general overuse | -0.0006 | -0.0028 | 0.0015 | 0.5528 |
| BMQ specific concerns | -0.0017 | -0.0033 | -0.0001 | 0.0380 |
| BMQ specific necessity | 0.0012 | -0.0009 | 0.0034 | 0.2644 |
| MARS-5 | 0.0022 | -0.0022 | 0.0067 | 0.3260 |
| PAM13-D score | 0.0003 | -0.0001 | 0.0008 | 0.1713 |
| ISSS-8 detrimental interaction | -0.0008 | -0.0028 | 0.0013 | 0.4516 |
| ISSS-8 positive support | -0.0001 | -0.0021 | 0.0019 | 0.9133 |

Table 6.14: Linear model for EQ5D VAS-AL at 1 y FU with adjustments for stratification and baseline variables. Complete case data analysis. Deceased patients excluded.

| Variable | Estimate | CI low | CI up | p.value |
|---|---|---|---|---|
| Intercept | 0.0998 | -0.0546 | 0.2541 | 0.2056 |
| SoC | -0.0203 | -0.0338 | -0.0067 | 0.0035 |
| AF | 0.0000 | -0.0172 | 0.0172 | 0.9974 |
| TRH | 0.0020 | -0.0143 | 0.0184 | 0.8061 |
| Outpatient | 0.0133 | -0.0278 | 0.0545 | 0.5263 |
| Female sex | 0.0009 | -0.0138 | 0.0156 | 0.9060 |
| Age | -0.0010 | -0.0016 | -0.0003 | 0.0061 |
| HeartQoL | 0.0513 | 0.0332 | 0.0694 | 0.0000 |
| EQ5D VAS | 0.0051 | 0.0046 | 0.0056 | 0.0000 |
| EQ5D index | 0.1162 | 0.0551 | 0.1773 | 0.0002 |
| GAD-7 | 0.0026 | -0.0003 | 0.0056 | 0.0800 |
| PHQ-9 | -0.0004 | -0.0034 | 0.0026 | 0.7888 |
| WHO-5 | 0.0004 | -0.0001 | 0.0008 | 0.1383 |
| BMQ general harm | 0.0040 | 0.0014 | 0.0066 | 0.0027 |
| BMQ general overuse | -0.0006 | -0.0028 | 0.0015 | 0.5528 |
| BMQ specific concerns | -0.0017 | -0.0033 | -0.0001 | 0.0380 |
| BMQ specific necessity | 0.0012 | -0.0009 | 0.0034 | 0.2644 |
| MARS-5 | 0.0022 | -0.0022 | 0.0067 | 0.3260 |
| PAM13-D score | 0.0003 | -0.0001 | 0.0008 | 0.1713 |
| ISSS-8 detrimental interaction | -0.0008 | -0.0028 | 0.0013 | 0.4516 |
| ISSS-8 positive support | -0.0001 | -0.0021 | 0.0019 | 0.9133 |

Table 6.15: Linear model for EQ5D VAS-AL at 1 y FU with adjustments for stratification and baseline variables. Multiple imputation data analysis. Deceased patients scored as 0.

| Variable | Estimate | CI low | CI up | p.value |
|---|---|---|---|---|
| Intercept | 0.1158 | -0.0332 | 0.2648 | 0.1284 |
| SoC | -0.0217 | -0.0350 | -0.0083 | 0.0015 |
| AF | 0.0001 | -0.0165 | 0.0166 | 0.9937 |
| TRH | -0.0050 | -0.0207 | 0.0107 | 0.5338 |
| Outpatient | 0.0228 | -0.0157 | 0.0612 | 0.2468 |
| Female sex | 0.0016 | -0.0125 | 0.0156 | 0.8285 |
| Age | -0.0008 | -0.0015 | -0.0002 | 0.0129 |
| HeartQoL | 0.0373 | 0.0191 | 0.0555 | 0.0001 |
| EQ5D VAS | 0.0051 | 0.0046 | 0.0056 | 0.0000 |
| EQ5D index | 0.1256 | 0.0659 | 0.1852 | 0.0000 |
| GAD-7 | 0.0021 | -0.0007 | 0.0050 | 0.1461 |
| PHQ-9 | -0.0001 | -0.0030 | 0.0028 | 0.9263 |
| WHO-5 | 0.0004 | 0.0000 | 0.0009 | 0.0790 |
| BMQ general harm | 0.0032 | 0.0006 | 0.0057 | 0.0146 |
| BMQ general overuse | -0.0006 | -0.0027 | 0.0015 | 0.5673 |
| BMQ specific concerns | -0.0012 | -0.0028 | 0.0004 | 0.1393 |
| BMQ specific necessity | 0.0007 | -0.0014 | 0.0027 | 0.5354 |
| MARS-5 | 0.0016 | -0.0028 | 0.0060 | 0.4764 |
| PAM13-D score | 0.0003 | -0.0002 | 0.0007 | 0.2775 |
| ISSS-8 detrimental interaction | -0.0004 | -0.0024 | 0.0016 | 0.6733 |
| ISSS-8 positive support | -0.0003 | -0.0022 | 0.0017 | 0.7686 |

Table 6.16: Linear model for EQ5D VAS-AL at 1 y FU with adjustments for stratification and baseline variables. Multiple imputation data analysis. Deceased patients excluded.

| Variable | Estimate | CI low | CI up | p.value |
|---|---|---|---|---|
| Intercept | 0.2571 | 0.1317 | 0.3826 | 0.0001 |
| SoC | -0.0157 | -0.0268 | -0.0046 | 0.0058 |
| AF | 0.0007 | -0.0130 | 0.0145 | 0.9180 |
| TRH | -0.0132 | -0.0266 | 0.0002 | 0.0544 |
| Outpatient | 0.0129 | -0.0191 | 0.0450 | 0.4285 |
| Female sex | -0.0009 | -0.0129 | 0.0112 | 0.8885 |
| Age | -0.0008 | -0.0014 | -0.0003 | 0.0032 |
| HeartQoL | 0.0359 | 0.0210 | 0.0508 | 0.0000 |
| EQ5D VAS | 0.0003 | -0.0001 | 0.0007 | 0.1762 |
| EQ5D index | 0.5871 | 0.5349 | 0.6393 | 0.0000 |
| GAD-7 | 0.0027 | 0.0002 | 0.0052 | 0.0363 |
| PHQ-9 | -0.0014 | -0.0038 | 0.0011 | 0.2655 |
| WHO-5 | 0.0003 | -0.0001 | 0.0007 | 0.1018 |
| BMQ general harm | 0.0013 | -0.0009 | 0.0035 | 0.2363 |
| BMQ general overuse | -0.0011 | -0.0028 | 0.0007 | 0.2324 |
| BMQ specific concerns | -0.0008 | -0.0022 | 0.0005 | 0.2105 |
| BMQ specific necessity | 0.0002 | -0.0016 | 0.0019 | 0.8517 |
| MARS-5 | 0.0007 | -0.0031 | 0.0044 | 0.7224 |
| PAM13-D score | 0.0002 | -0.0002 | 0.0006 | 0.2425 |
| ISSS-8 detrimental interaction | 0.0001 | -0.0016 | 0.0018 | 0.8894 |
| ISSS-8 positive support | 0.0009 | -0.0007 | 0.0025 | 0.2491 |

#### *6.5 Models with adjustment for stratification variables. Outcome variable: EQ5D index at 1 y FU*

Table 6.17: Linear model for EQ5D index at 1 y FU with adjustments for stratification variables. Complete case data analysis. Deceased patients scored as 0.

| Variable | Estimate | CI low | CI up | p.value |
|---|---|---|---|---|
| Intercept | 0.8056 | 0.7051 | 0.9060 | 0.0000 |
| SoC | -0.0722 | -0.1030 | -0.0415 | 0.0000 |
| AF | 0.0597 | 0.0221 | 0.0973 | 0.0019 |
| TRH | 0.0421 | 0.0055 | 0.0787 | 0.0245 |
| Outpatient | 0.0785 | -0.0083 | 0.1653 | 0.0768 |

Table 6.18: Linear model for EQ5D index at 1 y FU with adjustments for stratification variables. Complete case data analysis. Deceased patients excluded.

| Variable | Estimate | CI low | CI up | p.value |
|---|---|---|---|---|
| Intercept | 0.8559 | 0.7751 | 0.9367 | 0.0000 |
| SoC | -0.0371 | -0.0611 | -0.0132 | 0.0024 |
| AF | 0.0463 | 0.0168 | 0.0757 | 0.0022 |
| TRH | 0.0078 | -0.0206 | 0.0363 | 0.5900 |
| Outpatient | 0.0145 | -0.0562 | 0.0852 | 0.6873 |

Table 6.19: Linear model for EQ5D index at 1 y FU with adjustments for stratification variables. Multiple imputation data analysis. Deceased patients scored as 0.

| Variable | Estimate | CI low | CI up | p.value |
|---|---|---|---|---|
| Intercept | 0.7972 | 0.7295 | 0.8649 | 0.0000 |
| SoC | -0.0337 | -0.0561 | -0.0112 | 0.0034 |
| AF | 0.0417 | 0.0143 | 0.0690 | 0.0029 |
| TRH | 0.0062 | -0.0209 | 0.0332 | 0.6554 |
| Outpatient | 0.0324 | -0.0338 | 0.0985 | 0.3376 |

Table 6.20: Linear model for EQ5D index at 1 y FU with adjustments for stratification variables. Multiple imputation data analysis. Deceased patients excluded.

| Variable | Estimate | CI low | CI up | p.value |
|---|---|---|---|---|
| Intercept | 0.7971 | 0.7284 | 0.8658 | 0.0000 |
| SoC | -0.0343 | -0.0570 | -0.0116 | 0.0031 |
| AF | 0.0407 | 0.0131 | 0.0683 | 0.0039 |
| TRH | 0.0054 | -0.0213 | 0.0322 | 0.6899 |
| Outpatient | 0.0333 | -0.0335 | 0.1002 | 0.3289 |

*6.6 Models with adjustment for all variables. Outcome variable: EQ5D index at 1 y FU*

Table 6.21: Linear model for EQ5D index at 1 y FU with adjustments for stratification and baseline variables. Complete case data analysis. Deceased patients scored as 0.

| Variable | Estimate | CI low | CI up | p.value |
|---|---|---|---|---|
| Intercept | 0.4937 | 0.1935 | 0.7940 | 0.0013 |
| SoC | -0.0437 | -0.0702 | -0.0172 | 0.0013 |
| AF | 0.0276 | -0.0059 | 0.0610 | 0.1065 |
| TRH | 0.0100 | -0.0221 | 0.0421 | 0.5434 |
| Outpatient | 0.0121 | -0.0659 | 0.0901 | 0.7605 |
| Female sex | 0.0120 | -0.0165 | 0.0404 | 0.4105 |
| Age | -0.0027 | -0.0040 | -0.0014 | 0.0001 |
| HeartQoL | 0.0789 | 0.0438 | 0.1139 | 0.0000 |
| EQ5D VAS | 0.0004 | -0.0005 | 0.0014 | 0.3740 |
| EQ5D index | 0.4683 | 0.3521 | 0.5845 | 0.0000 |
| GAD-7 | -0.0007 | -0.0066 | 0.0051 | 0.8093 |
| PHQ-9 | 0.0007 | -0.0051 | 0.0066 | 0.8085 |
| WHO-5 | -0.0002 | -0.0011 | 0.0007 | 0.6292 |
| BMQ general harm | -0.0007 | -0.0058 | 0.0043 | 0.7787 |
| BMQ general overuse | -0.0020 | -0.0061 | 0.0021 | 0.3376 |
| BMQ specific concerns | 0.0006 | -0.0025 | 0.0037 | 0.7226 |
| BMQ specific necessity | 0.0010 | -0.0031 | 0.0050 | 0.6474 |
| MARS-5 | -0.0025 | -0.0113 | 0.0063 | 0.5764 |
| PAM13-D score | 0.0004 | -0.0005 | 0.0014 | 0.3705 |
| ISSS-8 detrimental interaction | 0.0006 | -0.0035 | 0.0046 | 0.7774 |
| ISSS-8 positive support | 0.0008 | -0.0032 | 0.0047 | 0.7066 |

Table 6.22: Linear model for EQ5D index at 1 y FU with adjustments for stratification and baseline variables. Complete case data analysis. Deceased patients excluded.

| Variable | Estimate | CI low | CI up | p.value |
|---|---|---|---|---|
| Intercept | 0.4490 | 0.2483 | 0.6496 | 0.0000 |
| SoC | -0.0169 | -0.0345 | 0.0007 | 0.0608 |
| AF | 0.0109 | -0.0114 | 0.0333 | 0.3369 |
| TRH | -0.0152 | -0.0366 | 0.0061 | 0.1631 |
| Outpatient | -0.0131 | -0.0669 | 0.0406 | 0.6328 |
| Female sex | -0.0003 | -0.0193 | 0.0188 | 0.9793 |
| Age | -0.0019 | -0.0028 | -0.0010 | 0.0000 |
| HeartQoL | 0.0629 | 0.0393 | 0.0864 | 0.0000 |
| EQ5D VAS | 0.0005 | -0.0001 | 0.0012 | 0.1188 |
| EQ5D index | 0.4388 | 0.3601 | 0.5175 | 0.0000 |
| GAD-7 | 0.0015 | -0.0023 | 0.0054 | 0.4328 |
| PHQ-9 | -0.0022 | -0.0061 | 0.0018 | 0.2789 |
| WHO-5 | 0.0003 | -0.0003 | 0.0009 | 0.3020 |
| BMQ general harm | 0.0030 | -0.0004 | 0.0064 | 0.0815 |
| BMQ general overuse | -0.0013 | -0.0041 | 0.0014 | 0.3470 |
| BMQ specific concerns | -0.0024 | -0.0045 | -0.0004 | 0.0212 |
| BMQ specific necessity | 0.0008 | -0.0019 | 0.0035 | 0.5748 |
| MARS-5 | -0.0002 | -0.0060 | 0.0055 | 0.9326 |
| PAM13-D score | 0.0002 | -0.0004 | 0.0008 | 0.5247 |
| ISSS-8 detrimental interaction | 0.0002 | -0.0025 | 0.0029 | 0.8825 |
| ISSS-8 positive support | 0.0008 | -0.0018 | 0.0034 | 0.5496 |

Table 6.23: Linear model for EQ5D index at 1 y FU with adjustments for stratification and baseline variables. Multiple imputation data analysis. Deceased patients scored as 0.

| Variable | Estimate | CI low | CI up | p.value |
|---|---|---|---|---|
| Intercept | 0.4551 | 0.2588 | 0.6513 | 0.0000 |
| SoC | -0.0195 | -0.0368 | -0.0022 | 0.0272 |
| AF | 0.0108 | -0.0101 | 0.0318 | 0.3120 |
| TRH | -0.0215 | -0.0422 | -0.0008 | 0.0423 |
| Outpatient | 0.0023 | -0.0484 | 0.0529 | 0.9302 |
| Female sex | 0.0017 | -0.0166 | 0.0199 | 0.8576 |
| Age | -0.0017 | -0.0026 | -0.0009 | 0.0001 |
| HeartQoL | 0.0458 | 0.0224 | 0.0692 | 0.0001 |
| EQ5D VAS | 0.0005 | -0.0001 | 0.0011 | 0.1074 |
| EQ5D index | 0.4761 | 0.3999 | 0.5523 | 0.0000 |
| GAD-7 | 0.0008 | -0.0029 | 0.0045 | 0.6702 |
| PHQ-9 | -0.0015 | -0.0053 | 0.0023 | 0.4405 |
| WHO-5 | 0.0002 | -0.0004 | 0.0008 | 0.5940 |
| BMQ general harm | 0.0020 | -0.0013 | 0.0052 | 0.2316 |
| BMQ general overuse | -0.0012 | -0.0039 | 0.0014 | 0.3720 |
| BMQ specific concerns | -0.0020 | -0.0041 | 0.0001 | 0.0611 |
| BMQ specific necessity | 0.0007 | -0.0019 | 0.0034 | 0.5848 |
| MARS-5 | -0.0016 | -0.0074 | 0.0042 | 0.5817 |
| PAM13-D score | 0.0003 | -0.0003 | 0.0009 | 0.2735 |
| ISSS-8 detrimental interaction | 0.0000 | -0.0026 | 0.0026 | 0.9930 |
| ISSS-8 positive support | 0.0005 | -0.0021 | 0.0030 | 0.7079 |

Table 6.24: Linear model for EQ5D index at 1 y FU with adjustments for stratification and baseline variables. Multiple imputation data analysis. Deceased patients excluded.

| Variable | Estimate | CI low | CI up | p.value |
|---|---|---|---|---|
| Intercept | 0.4556 | 0.2522 | 0.6589 | 0.0000 |
| SoC | -0.0199 | -0.0372 | -0.0027 | 0.0240 |
| AF | 0.0106 | -0.0110 | 0.0322 | 0.3373 |
| TRH | -0.0217 | -0.0424 | -0.0009 | 0.0415 |
| Outpatient | 0.0029 | -0.0488 | 0.0546 | 0.9122 |
| Female sex | 0.0020 | -0.0165 | 0.0205 | 0.8345 |
| Age | -0.0017 | -0.0026 | -0.0009 | 0.0001 |
| HeartQoL | 0.0458 | 0.0221 | 0.0695 | 0.0002 |
| EQ5D VAS | 0.0005 | -0.0002 | 0.0011 | 0.1503 |
| EQ5D index | 0.4783 | 0.3996 | 0.5570 | 0.0000 |
| GAD-7 | 0.0009 | -0.0029 | 0.0046 | 0.6575 |
| PHQ-9 | -0.0017 | -0.0055 | 0.0020 | 0.3654 |
| WHO-5 | 0.0001 | -0.0005 | 0.0007 | 0.6390 |
| BMQ general harm | 0.0018 | -0.0015 | 0.0052 | 0.2887 |
| BMQ general overuse | -0.0012 | -0.0039 | 0.0014 | 0.3681 |
| BMQ specific concerns | -0.0019 | -0.0039 | 0.0002 | 0.0713 |
| BMQ specific necessity | 0.0007 | -0.0021 | 0.0035 | 0.6092 |
| MARS-5 | -0.0016 | -0.0074 | 0.0042 | 0.5884 |
| PAM13-D score | 0.0004 | -0.0002 | 0.0010 | 0.2244 |
| ISSS-8 detrimental interaction | 0.0000 | -0.0025 | 0.0025 | 0.9998 |
| ISSS-8 positive support | 0.0004 | -0.0021 | 0.0030 | 0.7396 |

#### *6.7 Models with adjustment for stratification variables. Outcome variable: EQ5D VAS at 1 y FU*

Table 6.25: Linear model for EQ5D VAS at 1 y FU with adjustments for stratification variables. Complete case data analysis. Deceased patients scored as 0.

| Variable | Estimate | CI low | CI up | p.value |
|---|---|---|---|---|
| Intercept | 65.9542 | 56.8338 | 75.0747 | 0.0000 |
| SoC | -4.4016 | -7.0898 | -1.7134 | 0.0014 |
| AF | 5.4119 | 2.1024 | 8.7214 | 0.0014 |
| TRH | 2.8557 | -0.3360 | 6.0474 | 0.0799 |
| Outpatient | 8.1238 | 0.1295 | 16.1181 | 0.0468 |

Table 6.26: Linear model for EQ5D VAS at 1 y FU with adjustments for stratification variables. Complete case data analysis. Deceased patients excluded.

| Variable | Estimate | CI low | CI up | p.value |
|---|---|---|---|---|
| Intercept | 65.9542 | 56.8338 | 75.0747 | 0.0000 |
| SoC | -4.4016 | -7.0898 | -1.7134 | 0.0014 |
| AF | 5.4119 | 2.1024 | 8.7214 | 0.0014 |
| TRH | 2.8557 | -0.3360 | 6.0474 | 0.0799 |
| Outpatient | 8.1238 | 0.1295 | 16.1181 | 0.0468 |

Table 6.27: Linear model for EQ5D VAS at 1 y FU with adjustments for stratification variables. Multiple imputation data analysis. Deceased patients scored as 0.

| Variable | Estimate | CI low | CI up | p.value |
|---|---|---|---|---|
| Intercept | 60.7017 | 52.7383 | 68.6652 | 0.0000 |
| SoC | -3.6864 | -6.3312 | -1.0417 | 0.0065 |
| AF | 4.4596 | 1.2317 | 7.6875 | 0.0069 |
| TRH | 2.6874 | -0.4231 | 5.7979 | 0.0908 |
| Outpatient | 8.2564 | 0.4875 | 16.0253 | 0.0377 |

Table 6.28: Linear model for EQ5D VAS at 1 y FU with adjustments for stratification variables. Multiple imputation data analysis. Deceased patients excluded.

| Variable | Estimate | CI low | CI up | p.value |
|---|---|---|---|---|
| Intercept | 60.5404 | 52.5333 | 68.5476 | 0.0000 |
| SoC | -3.7145 | -6.3041 | -1.1249 | 0.0051 |
| AF | 4.4573 | 1.2224 | 7.6923 | 0.0071 |
| TRH | 2.6141 | -0.5258 | 5.7541 | 0.1032 |
| Outpatient | 8.4531 | 0.7273 | 16.1789 | 0.0324 |

#### *6.8 Models with adjustment for all variables. Outcome variable: EQ5D VAS at 1 y FU*

Table 6.29: Linear model for EQ5D VAS at 1 y FU with adjustments for stratification and baseline variables. Complete case data analysis. Deceased patients scored as 0.

| Variable | Estimate | CI low | CI up | p.value |
|---|---|---|---|---|
| Intercept | 19.2388 | -6.0424 | 44.5200 | 0.1363 |
| SoC | -3.0504 | -5.2701 | -0.8307 | 0.0072 |
| AF | 0.7391 | -2.0725 | 3.5508 | 0.6066 |
| TRH | -0.2040 | -2.8887 | 2.4806 | 0.8816 |
| Outpatient | 4.1549 | -2.6529 | 10.9627 | 0.2320 |
| Female sex | 0.5758 | -1.8336 | 2.9851 | 0.6397 |
| Age | -0.1902 | -0.3009 | -0.0795 | 0.0008 |
| HeartQoL | 6.8829 | 3.9273 | 9.8386 | 0.0000 |
| EQ5D VAS | 0.3303 | 0.2499 | 0.4108 | 0.0000 |
| EQ5D index | 14.6363 | 4.6921 | 24.5804 | 0.0040 |
| GAD-7 | 0.4125 | -0.0737 | 0.8987 | 0.0968 |
| PHQ-9 | -0.3156 | -0.8090 | 0.1779 | 0.2105 |
| WHO-5 | 0.0092 | -0.0683 | 0.0868 | 0.8154 |
| BMQ general harm | 0.4599 | 0.0316 | 0.8881 | 0.0357 |
| BMQ general overuse | -0.0500 | -0.3987 | 0.2988 | 0.7790 |
| BMQ specific concerns | -0.2100 | -0.4715 | 0.0515 | 0.1159 |
| BMQ specific necessity | 0.0413 | -0.3062 | 0.3888 | 0.8159 |
| MARS-5 | 0.3982 | -0.3376 | 1.1341 | 0.2892 |
| PAM13-D score | 0.0354 | -0.0426 | 0.1133 | 0.3745 |
| ISSS-8 detrimental interaction | -0.0591 | -0.3958 | 0.2775 | 0.7307 |
| ISSS-8 positive support | 0.1513 | -0.1770 | 0.4795 | 0.3667 |

Table 6.30: Linear model for EQ5D VAS at 1 y FU with adjustments for stratification and baseline variables. Complete case data analysis. Deceased patients excluded.

| Variable | Estimate | CI low | CI up | p.value |
|---|---|---|---|---|
| Intercept | 19.2388 | -6.0424 | 44.5200 | 0.1363 |
| SoC | -3.0504 | -5.2701 | -0.8307 | 0.0072 |
| AF | 0.7391 | -2.0725 | 3.5508 | 0.6066 |
| TRH | -0.2040 | -2.8887 | 2.4806 | 0.8816 |
| Outpatient | 4.1549 | -2.6529 | 10.9627 | 0.2320 |
| Female sex | 0.5758 | -1.8336 | 2.9851 | 0.6397 |
| Age | -0.1902 | -0.3009 | -0.0795 | 0.0008 |
| HeartQoL | 6.8829 | 3.9273 | 9.8386 | 0.0000 |
| EQ5D VAS | 0.3303 | 0.2499 | 0.4108 | 0.0000 |
| EQ5D index | 14.6363 | 4.6921 | 24.5804 | 0.0040 |
| GAD-7 | 0.4125 | -0.0737 | 0.8987 | 0.0968 |
| PHQ-9 | -0.3156 | -0.8090 | 0.1779 | 0.2105 |
| WHO-5 | 0.0092 | -0.0683 | 0.0868 | 0.8154 |
| BMQ general harm | 0.4599 | 0.0316 | 0.8881 | 0.0357 |
| BMQ general overuse | -0.0500 | -0.3987 | 0.2988 | 0.7790 |
| BMQ specific concerns | -0.2100 | -0.4715 | 0.0515 | 0.1159 |
| BMQ specific necessity | 0.0413 | -0.3062 | 0.3888 | 0.8159 |
| MARS-5 | 0.3982 | -0.3376 | 1.1341 | 0.2892 |
| PAM13-D score | 0.0354 | -0.0426 | 0.1133 | 0.3745 |
| ISSS-8 detrimental interaction | -0.0591 | -0.3958 | 0.2775 | 0.7307 |
| ISSS-8 positive support | 0.1513 | -0.1770 | 0.4795 | 0.3667 |

Table 6.31: Linear model for EQ5D VAS at 1 y FU with adjustments for stratification and baseline variables. Multiple imputation data analysis. Deceased patients scored as 0.

| Variable | Estimate | CI low | CI up | p.value |
|---|---|---|---|---|
| Intercept | 19.7843 | -5.4494 | 45.0179 | 0.1250 |
| SoC | -2.7034 | -4.9117 | -0.4951 | 0.0167 |
| AF | 0.3714 | -2.3693 | 3.1121 | 0.7906 |
| TRH | -1.0442 | -3.6630 | 1.5747 | 0.4348 |
| Outpatient | 4.4603 | -2.0643 | 10.9849 | 0.1809 |
| Female sex | 0.3709 | -1.9643 | 2.7061 | 0.7557 |
| Age | -0.1869 | -0.2938 | -0.0799 | 0.0007 |
| HeartQoL | 4.2311 | 1.2884 | 7.1737 | 0.0051 |
| EQ5D VAS | 0.3469 | 0.2648 | 0.4290 | 0.0000 |
| EQ5D index | 19.6744 | 9.9536 | 29.3952 | 0.0001 |
| GAD-7 | 0.2867 | -0.2011 | 0.7746 | 0.2500 |
| PHQ-9 | -0.1865 | -0.6684 | 0.2954 | 0.4485 |
| WHO-5 | 0.0193 | -0.0571 | 0.0957 | 0.6205 |
| BMQ general harm | 0.3521 | -0.0746 | 0.7788 | 0.1064 |
| BMQ general overuse | -0.0688 | -0.4089 | 0.2713 | 0.6918 |
| BMQ specific concerns | -0.1965 | -0.4570 | 0.0641 | 0.1401 |
| BMQ specific necessity | -0.0020 | -0.3492 | 0.3453 | 0.9911 |
| MARS-5 | 0.2969 | -0.4562 | 1.0501 | 0.4401 |
| PAM13-D score | 0.0251 | -0.0523 | 0.1025 | 0.5250 |
| ISSS-8 detrimental interaction | -0.1204 | -0.4520 | 0.2113 | 0.4773 |
| ISSS-8 positive support | 0.1752 | -0.1516 | 0.5020 | 0.2938 |

Table 6.32: Linear model for EQ5D VAS at 1 y FU with adjustments for stratification and baseline variables. Multiple imputation data analysis. Deceased patients excluded.

| Variable | Estimate | CI low | CI up | p.value |
|---|---|---|---|---|
| Intercept | 18.9805 | -7.5188 | 45.4798 | 0.1611 |
| SoC | -2.7343 | -4.8923 | -0.5763 | 0.0133 |
| AF | 0.4480 | -2.3227 | 3.2187 | 0.7514 |
| TRH | -1.0740 | -3.7339 | 1.5859 | 0.4291 |
| Outpatient | 4.6564 | -1.7617 | 11.0746 | 0.1556 |
| Female sex | 0.4898 | -1.8690 | 2.8486 | 0.6842 |
| Age | -0.1855 | -0.2963 | -0.0747 | 0.0011 |
| HeartQoL | 4.2590 | 1.3024 | 7.2156 | 0.0050 |
| EQ5D VAS | 0.3425 | 0.2625 | 0.4226 | 0.0000 |
| EQ5D index | 19.8543 | 10.1064 | 29.6022 | 0.0001 |
| GAD-7 | 0.3109 | -0.1639 | 0.7858 | 0.2000 |
| PHQ-9 | -0.2035 | -0.6916 | 0.2846 | 0.4143 |
| WHO-5 | 0.0205 | -0.0567 | 0.0977 | 0.6032 |
| BMQ general harm | 0.3670 | -0.0750 | 0.8089 | 0.1044 |
| BMQ general overuse | -0.0694 | -0.4148 | 0.2761 | 0.6940 |
| BMQ specific concerns | -0.2022 | -0.4649 | 0.0605 | 0.1320 |
| BMQ specific necessity | 0.0134 | -0.3317 | 0.3586 | 0.9393 |
| MARS-5 | 0.3020 | -0.4789 | 1.0828 | 0.4489 |
| PAM13-D score | 0.0255 | -0.0513 | 0.1024 | 0.5148 |
| ISSS-8 detrimental interaction | -0.1243 | -0.4568 | 0.2081 | 0.4638 |
| ISSS-8 positive support | 0.1740 | -0.1584 | 0.5063 | 0.3054 |

### 7. Frequency of levels per dimension by treatment group over time

Table 7.1: Frequency of levels of mobility at baseline, 6 and 12 months follow-up.

| | Baseline | | | | Follow up at 6 months | | | | Follow up at 12 months | | | |
|---|---|---|---|---|---|---|---|---|---|---|---|---|
| Mobility | NICC | | SoC | | NICC | | SoC | | NICC | | SoC | |
| EQ5D level | n | Perc | n | Perc | n | Perc | n | Perc | n | Perc | n | Perc |
| 1 | 216 | 49.09 | 193 | 42.98 | 221 | 57.11 | 200 | 46.73 | 223 | 58.53 | 180 | 44.89 |
| 2 | 101 | 22.95 | 98 | 21.83 | 76 | 19.64 | 102 | 23.83 | 69 | 18.11 | 94 | 23.44 |
| 3 | 85 | 19.32 | 107 | 23.83 | 65 | 16.80 | 75 | 17.52 | 52 | 13.65 | 82 | 20.45 |
| 4 | 36 | 8.18 | 51 | 11.36 | 25 | 6.46 | 49 | 11.45 | 34 | 8.92 | 44 | 10.97 |
| 5 | 2 | 0.45 | 0 | 0.00 | 0 | 0.00 | 2 | 0.47 | 3 | 0.79 | 1 | 0.25 |
| Problems | 224 | 50.91 | 256 | 57.02 | 166 | 42.89 | 228 | 53.27 | 158 | 41.47 | 221 | 55.11 |
| Change in problems | | | | | | -8.02 | | -3.74 | | -9.44 | | -1.90 |

Table 7.2: Frequency of levels of selfcare at baseline, 6 and 12 months follow-up.

| | Baseline | | | | Follow up at 6 months | | | | Follow up at 12 months | | | |
|---|---|---|---|---|---|---|---|---|---|---|---|---|
| **Selfcare** | **NICC** | | **SoC** | | **NICC** | | **SoC** | | **NICC** | | **SoC** | |
| **EQ5D level** | **n** | **Perc** | **n** | **Perc** | **n** | **Perc** | **n** | **Perc** | **n** | **Perc** | **n** | **Perc** |
| 1 | 368 | 83.26 | 359 | 79.42 | 333 | 86.05 | 345 | 80.99 | 327 | 85.83 | 306 | 76.12 |
| 2 | 40 | 9.05 | 46 | 10.18 | 34 | 8.79 | 38 | 8.92 | 27 | 7.09 | 47 | 11.69 |
| 3 | 27 | 6.11 | 36 | 7.96 | 12 | 3.10 | 29 | 6.81 | 20 | 5.25 | 35 | 8.71 |
| 4 | 5 | 1.13 | 9 | 1.99 | 6 | 1.55 | 13 | 3.05 | 7 | 1.84 | 13 | 3.23 |
| 5 | 2 | 0.45 | 2 | 0.44 | 2 | 0.52 | 1 | 0.23 | 0 | 0.00 | 1 | 0.25 |
| Problems | 74 | 16.74 | 93 | 20.58 | 54 | 13.95 | 81 | 19.01 | 54 | 14.17 | 96 | 23.88 |
| Change in problems | | | | | | -2.79 | | -1.56 | | -2.57 | | 3.31 |

Table 7.3: Frequency of levels of usual activities at baseline, 6 and 12 months follow-up.

| | Baseline | | | | Follow up at 6 months | | | | Follow up at 12 months | | | |
|---|---|---|---|---|---|---|---|---|---|---|---|---|
| **Usual activities** | **NICC** | | **SoC** | | **NICC** | | **SoC** | | **NICC** | | **SoC** | |
| **EQ5D level** | **n** | **Perc** | **n** | **Perc** | **n** | **Perc** | **n** | **Perc** | **n** | **Perc** | **n** | **Perc** |
| 1 | 230 | 52.15 | 232 | 51.33 | 222 | 57.07 | 223 | 52.22 | 231 | 60.47 | 212 | 53.00 |
| 2 | 120 | 27.21 | 108 | 23.89 | 105 | 26.99 | 103 | 24.12 | 83 | 21.73 | 84 | 21.00 |
| 3 | 70 | 15.87 | 83 | 18.36 | 45 | 11.57 | 75 | 17.56 | 48 | 12.57 | 72 | 18.00 |
| 4 | 19 | 4.31 | 25 | 5.53 | 15 | 3.86 | 24 | 5.62 | 17 | 4.45 | 28 | 7.00 |
| 5 | 2 | 0.45 | 4 | 0.88 | 2 | 0.51 | 2 | 0.47 | 3 | 0.79 | 4 | 1.00 |
| Problems | 211 | 47.85 | 220 | 48.67 | 167 | 42.93 | 204 | 47.78 | 151 | 39.53 | 188 | 47.00 |
| Change in problems | | | | | | -4.92 | | -0.90 | | -8.32 | | -1.67 |

Table 7.4: Frequency of levels of pain at baseline, 6 and 12 months follow-up.

| | Baseline | | | | Follow up at 6 months | | | | Follow up at 12 months | | | |
|---|---|---|---|---|---|---|---|---|---|---|---|---|
| **Pain** | **NICC** | | **SoC** | | **NICC** | | **SoC** | | **NICC** | | **SoC** | |
| **EQ5D level** | **n** | **Perc** | **n** | **Perc** | **n** | **Perc** | **n** | **Perc** | **n** | **Perc** | **n** | **Perc** |
| 1 | 124 | 28.05 | 104 | 23.06 | 140 | 36.08 | 111 | 26.12 | 122 | 32.28 | 101 | 25.51 |
| 2 | 172 | 38.91 | 151 | 33.48 | 137 | 35.31 | 148 | 34.82 | 133 | 35.19 | 147 | 37.12 |
| 3 | 107 | 24.21 | 151 | 33.48 | 79 | 20.36 | 121 | 28.47 | 89 | 23.54 | 105 | 26.52 |
| 4 | 37 | 8.37 | 41 | 9.09 | 30 | 7.73 | 42 | 9.88 | 32 | 8.47 | 42 | 10.61 |
| 5 | 2 | 0.45 | 4 | 0.89 | 2 | 0.52 | 3 | 0.71 | 2 | 0.53 | 1 | 0.25 |
| Problems | 318 | 71.95 | 347 | 76.94 | 248 | 63.92 | 314 | 73.88 | 256 | 67.72 | 295 | 74.49 |
| Change in problems | | | | | | -8.03 | | -3.06 | | -4.22 | | -2.45 |

Table 7.5: Frequency of levels of anxiety and depression at baseline, 6 and 12 months follow-up.

| Anxiety/depression | Baseline | | | | Follow up at 6 months | | | | Follow up at 12 months | | | |
|---|---|---|---|---|---|---|---|---|---|---|---|---|
| | NICC | | SoC | | NICC | | SoC | | NICC | | SoC | |
| **EQ5D level** | **n** | **Perc** | **n** | **Perc** | **n** | **Perc** | **n** | **Perc** | **n** | **Perc** | **n** | **Perc** |
| 1 | 281 | 63.86 | 269 | 59.78 | 261 | 67.27 | 263 | 61.74 | 261 | 68.50 | 240 | 60.15 |
| 2 | 111 | 25.23 | 107 | 23.78 | 101 | 26.03 | 104 | 24.41 | 86 | 22.57 | 97 | 24.31 |
| 3 | 39 | 8.86 | 57 | 12.67 | 19 | 4.90 | 44 | 10.33 | 29 | 7.61 | 46 | 11.53 |
| 4 | 8 | 1.82 | 14 | 3.11 | 7 | 1.80 | 15 | 3.52 | 5 | 1.31 | 16 | 4.01 |
| 5 | 1 | 0.23 | 3 | 0.67 | 0 | 0.00 | 0 | 0.00 | 0 | 0.00 | 0 | 0.00 |
| Problems | 159 | 36.14 | 181 | 40.22 | 127 | 32.73 | 163 | 38.26 | 120 | 31.50 | 159 | 39.85 |
| Change in problems | | | | | | -3.40 | | -1.96 | | -4.64 | | -0.37 |

### 8. Visualize EQ5D score and EQ5D index

#### *8.1 Boxplot & volinplot*

#### 8.1.1 EQ5D index

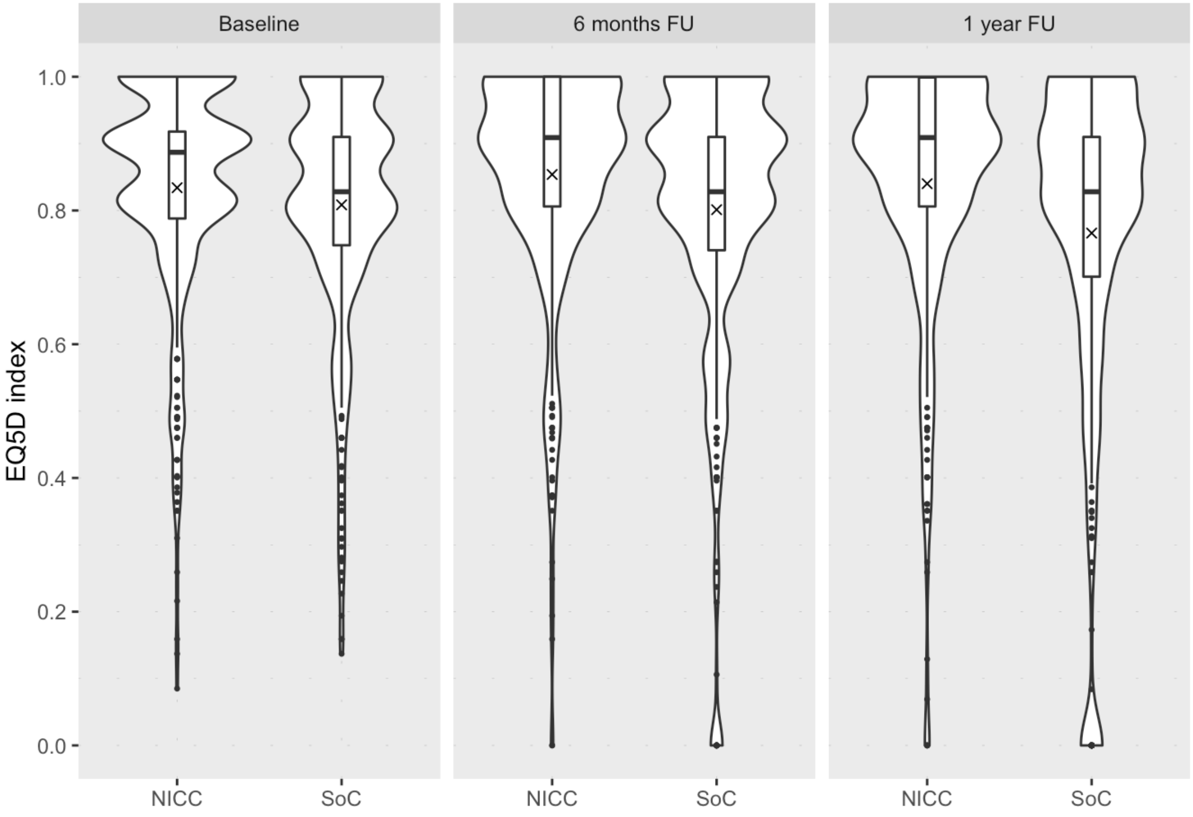

8.1.2 EQ5D VAS score

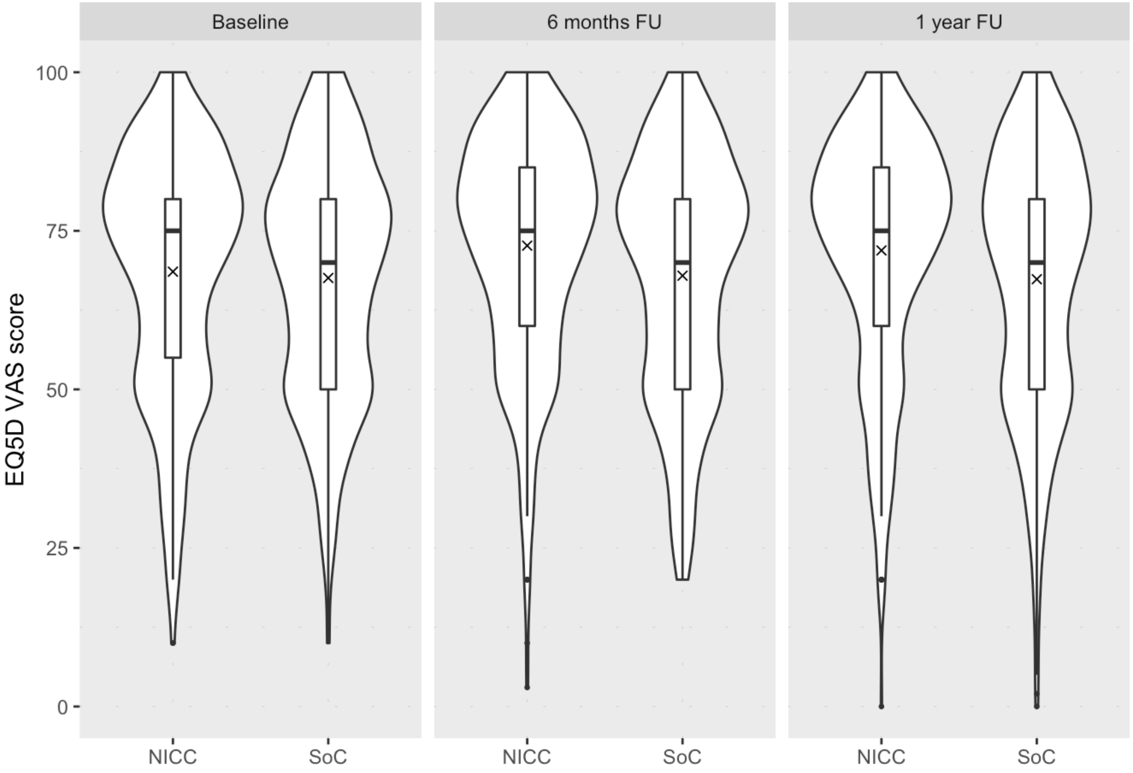

*8.2 Histograms*

### 8.2.1 EQ5D index

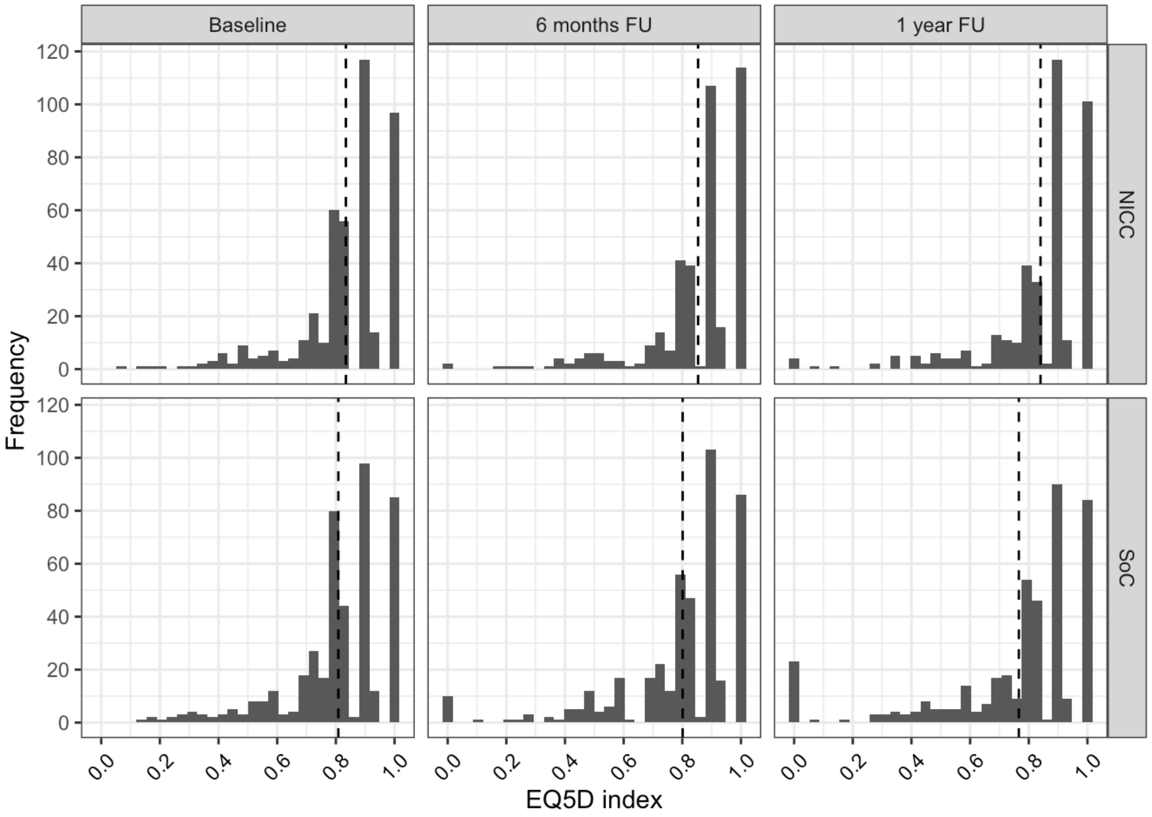


### 8.2.2 EQ5D VAS score

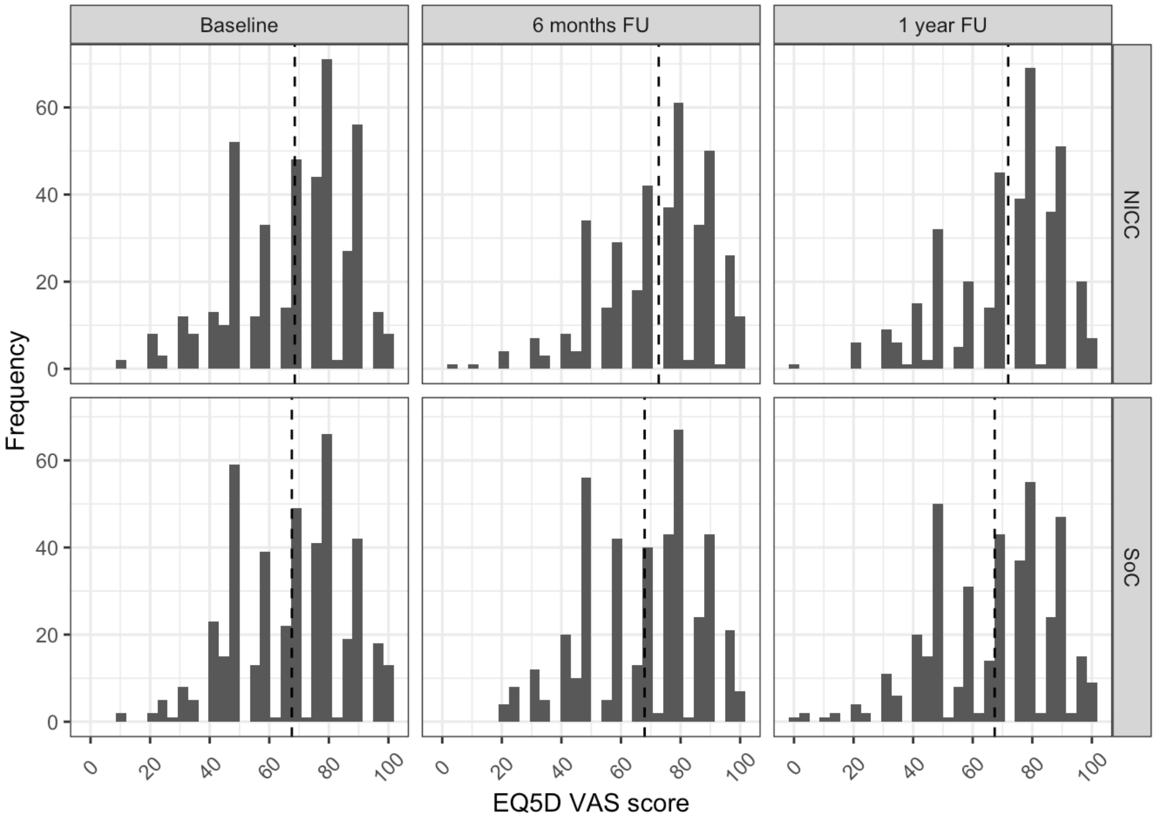

### 9. Information about R session

The analysis started on 2021-05-06 12:56:50 and ended on 2021-05-06 12:57:49. Calculations require 58.427973985672. Analyses were done with R version x86_64-apple-darwin17.0, x86_64, darwin17.0, x86_64, darwin17.0, , 4, 0.2, 2020, 06, 22, 78730, R, R version 4.0.2 (2020-06-22), Taking Off Again.

The following R packages were used in the analyses:

```
## R version 4.0.2 (2020-06-22)
## Platform: x86_64-apple-darwin17.0 (64-bit)
## Running under: macOS Catalina 10.15.7
##
## Matrix products: default
## BLAS:   /Library/Frameworks/R.framework/Versions/4.0/Resources/lib/libRblas.dylib
##                                                                          LAPACK:
/Library/Frameworks/R.framework/Versions/4.0/Resources/lib/libRlapack.dylib
##
## locale:
## [1] de_DE.UTF-8/de_DE.UTF-8/de_DE.UTF-8/C/de_DE.UTF-8/de_DE.UTF-8
##
## attached base packages:
## [1] grid      stats     graphics  grDevices utils     datasets  methods
## [8] base
##
## other attached packages:
##  [1] here_1.0.1        VIM_6.1.0         colorspace_2.0-1 xtable_1.8-4
##  [5] mitools_2.4       mice_3.13.0       kableExtra_1.3.4 knitr_1.33
##  [9] arsenal_3.6.2     sjlabelled_1.1.7 sjPlot_2.8.7     sjmisc_2.8.6
## [13] forcats_0.5.1     stringr_1.4.0     purrr_0.3.4      readr_1.4.0
## [17] tidyr_1.1.3       tibble_3.1.1      ggplot2_3.3.3    tidyverse_1.3.1
## [21] dplyr_1.0.6
##
## loaded via a namespace (and not attached):
##   [1] TH.data_1.0-10      minqa_1.2.4         ellipsis_0.3.2
##   [4] class_7.3-19        rio_0.5.26          rprojroot_2.0.2
##   [7] estimability_1.3    parameters_0.13.0   fs_1.5.0
##  [10] rstudioapi_0.13     proxy_0.4-25        farver_2.1.0
##  [13] fansi_0.4.2         mvtnorm_1.1-1       lubridate_1.7.10
##  [16] ranger_0.12.1       xml2_1.3.2          codetools_0.2-18
##  [19] splines_4.0.2       robustbase_0.93-7   jsonlite_1.7.2
##  [22] nloptr_1.2.2.2      ggeffects_1.1.0     broom_0.7.6
##  [25] dbplyr_2.1.1        effectsize_0.4.4-1 compiler_4.0.2
##  [28] httr_1.4.2          sjstats_0.18.1      emmeans_1.6.0
##  [31] backports_1.2.1     assertthat_0.2.1    Matrix_1.2-18
##  [34] cli_2.5.0           htmltools_0.5.1.1   tools_4.0.2
##  [37] coda_0.19-4         gtable_0.3.0        glue_1.4.2
##  [40] Rcpp_1.0.6          carData_3.0-4       cellranger_1.1.0
##  [43] vctrs_0.3.8         svglite_2.0.0       nlme_3.1-152
##  [46] lmtest_0.9-38       insight_0.13.2      laeken_0.5.1
##  [49] xfun_0.22           openxlsx_4.2.3      lme4_1.1-26
##  [52] rvest_1.0.0         lifecycle_1.0.0     statmod_1.4.35
##  [55] DEoptimR_1.0-8      MASS_7.3-54         zoo_1.8-9
##  [58] scales_1.1.1        hms_1.0.0           sandwich_3.0-0
##  [61] yaml_2.2.1          curl_4.3.1          stringi_1.5.3
##  [64] highr_0.9           bayestestR_0.9.0    e1071_1.7-6
##  [67] zip_2.1.1           boot_1.3-28         rlang_0.4.11
##  [70] pkgconfig_2.0.3     systemfonts_1.0.1   evaluate_0.14
##  [73] lattice_0.20-44     labeling_0.4.2      tidyselect_1.1.1
##  [76] magrittr_2.0.1      bookdown_0.22       R6_2.5.0
##  [79] generics_0.1.0      multcomp_1.4-17     DBI_1.1.1
##  [82] pillar_1.6.0        haven_2.4.1         foreign_0.8-81
##  [85] withr_2.4.2         abind_1.4-5         survival_3.2-11
##  [88] sp_1.4-5            nnet_7.3-16         performance_0.7.1
##  [91] modelr_0.1.8        crayon_1.4.1        car_3.0-10
##  [94] utf8_1.2.1          rmarkdown_2.7       readxl_1.3.1
```

```
##  [97] data.table_1.14.0  vcd_1.4-8          reprex_2.0.0
## [100] digest_0.6.27      webshot_0.5.2      munsell_0.5.0
## [103] viridisLite_0.4.0
```
